# Joint genotypic and phenotypic outcome modeling improves base editing variant effect quantification

**DOI:** 10.1101/2023.09.08.23295253

**Authors:** Jayoung Ryu, Sam Barkal, Tian Yu, Martin Jankowiak, Yunzhuo Zhou, Matthew Francoeur, Quang Vinh Phan, Zhijian Li, Manuel Tognon, Lara Brown, Michael I. Love, Guillaume Lettre, David B. Ascher, Christopher A. Cassa, Richard I. Sherwood, Luca Pinello

**Author notes:** Corresponding authors (C.A.C.), (R.I.S.), (L.P.).

## Abstract

CRISPR base editing screens are powerful tools for studying disease-associated variants at scale. However, the efficiency and precision of base editing perturbations vary, confounding the assessment of variant-induced phenotypic effects. Here, we provide an integrated pipeline that improves the estimation of variant impact in base editing screens. We perform high-throughput ABE8e-SpRY base editing screens with an integrated reporter construct to measure the editing efficiency and outcomes of each gRNA alongside their phenotypic consequences. We introduce BEAN, a Bayesian network that accounts for per-guide editing outcomes and target site chromatin accessibility to estimate variant impacts. We show this pipeline attains superior performance compared to existing tools in variant classification and effect size quantification. We use BEAN to pinpoint common variants that alter LDL uptake, implicating novel genes. Additionally, through saturation base editing of *LDLR*, we enable accurate quantitative prediction of the effects of missense variants on LDL-C levels, which aligns with measurements in UK Biobank individuals, and identify structural mechanisms underlying variant pathogenicity. This work provides a widely applicable approach to improve the power of base editor screens for disease-associated variant characterization.

## Introduction

Genetic variation contributes substantially to complex disease risk. While well-powered genome-wide association studies (GWAS)^1^ and rare variant analyses from cohort studies such as the UK Biobank (UKB)^2^ have associated thousands of loci and genes with clinical phenotypes, these observational approaches are often insufficient to identify causal variants. Perturbation-based methods enable evaluation of the impact of an individual variant in a common genetic background, isolated from genetically linked variants, and such testing can be performed in high throughput through multiplex assays of variant effect (MAVEs)^3^. Numerous types of MAVEs have been developed, including deep mutational scanning (DMS)^4^, saturation mutagenesis^5^, massively parallel reporter assays (MPRA)^6^, and CRISPR-based screens^7–9^.

CRISPR base editing screens have emerged as a uniquely powerful method to study variants in their endogenous genomic context. Base editors, fusions of Cas9-nickase and single-stranded cytosine or adenine deaminase enzymes^10,11^, enable site-specific installation of transition variants. As the majority of disease-associated variants are single-nucleotide transitions^12^, base editors enable the installation of functionally relevant variants in a precise and scalable way. Base editing screens have been employed to dissect coding variant effects as well as to evaluate GWAS-associated variant functions^13–28^.

However, base editing efficiency varies substantially depending on the local sequence context surrounding the target base, the specific Cas9 variant and deaminase used, and the cellular context^29^. Moreover, base edits can occur at multiple positions within the single-stranded DNA bubble created by the guide RNA (gRNA)-DNA binding on the opposite strand, therefore a single gRNA can install a variety of alleles, each with distinct efficiencies. While there have been efforts to predict editing outcomes using massively parallel base editor reporter assay data^29^, these predictions do not generalize well to unprofiled base editors and cellular contexts^20^.

In previous base editing screens, analysis of phenotypic outcomes is confounded by variable editing efficiencies and outcomes. Phenotypic effects of gRNAs with robust editing are exaggerated, and effects of variants that are not installed as efficiently are underestimated. Such confounding is especially pernicious when the target elements are coding variants, as a single gRNA may install distinct coding variants with different frequencies, and current analysis methods are unable to deconvolve such data.

Existing base editing screens have dealt with the heterogeneity in gRNA efficiency and genotypic outcomes in several ways. One approach that has been employed is to assume all editable nucleotides within the editing window are edited with uniform efficiency^13^. Two recent studies have profiled the gRNAs used in phenotypic base editing screening using a base editor reporter (or sensor) assay^20,21^ to filter gRNAs with low editing efficiency when analyzing their phenotypic data. Despite these initial efforts, the computational analyses of these screens have not yet been formalized, often relying on existing tools that were not designed specifically for base editor data with or without the target site reporter.

Here, we design an experimental-computational pipeline to improve the accuracy of variant effect estimation in base editing screens. By incorporating a target site reporter sequence into the gRNA construct, we simultaneously measure the editing efficiency of a gRNA and its phenotypic impact. We develop a computational pipeline, BEAN, that normalizes the phenotypic scores of target variants using genotypic outcome information collected from the target site reporter. Moreover, we extend BEAN to analyze densely tiled coding sequence base editing screen data, sharing information among neighboring gRNAs to obtain accurate phenotypic scores for each coding variant. BEAN provides a first-in-class integrated solution to experimental assessment of variant effects through base editing screens. We systematically benchmark BEAN against current state-of-the-art methods for the analyses of pooled CRISPR screens and show substantially improved performance of BEAN.

To leverage activity-normalized base editing screening, we have conducted screens assessing the impact of low-density lipoprotein cholesterol (LDL-C)-associated GWAS variants and low-density lipoprotein receptor (*LDLR*) coding variants on LDL-C uptake in HepG2 hepatocellular carcinoma cells. Genetic differences in LDL-C levels contribute substantially to coronary artery disease risk. Serum LDL-C measurements are quantitative and nearly uniformly measured in most biobanks, and thus they provide among the highest quality human phenotypic data for any trait. A trans-ancestry GWAS meta-analysis from the Global Lipids Genetics Consortium (GLGC) has identified >900 genome-wide significant loci associated with blood lipid levels, including >400 loci associated with LDL-C^30^. LDL-C GWAS loci overlap strongly with liver-enriched gene expression, nominating liver as the primary tissue driving LDL-C variant effects^31,32^. Yet, the causal variants and mechanisms by which many of these loci modulate LDL-C levels remain unknown.

LDL-C levels are also impacted by rare coding variants. In the most severe instances, inherited monogenic variants in several genes cause Familial Hypercholesterolemia (FH), a disease associated with extremely elevated LDL-C levels and premature cardiovascular disease^33^. The majority of genetic mutations known to cause FH occur in *LDLR*, a cell surface receptor that uptakes LDL, thus removing it from circulation^34^. Despite the effectiveness of lipid lowering therapies, FH patients are still 2-4-fold more likely to have coronary events than the general population^35^. Elevated LDL-C levels increase cardiovascular disease risk throughout life, so the early identification of at-risk individuals would have immense clinical utility^33^. However, many *LDLR* variants currently lack clinical interpretation. Of the 1,427 *LDLR* missense variants in the ClinVar database^36^, 50% are classified as variants of unknown significance (VUS) or to have conflicting interpretations of pathogenicity (“conflicting”), thus impeding FH diagnosis. Likewise, of the 758 unique *LDLR* missense variants carried by sequenced individuals in the UKB cohort, 69% are either unreported or have an uncertain annotation in ClinVar. Altogether, improved understanding of *LDLR* variant impacts would enable earlier diagnosis and treatment for a large number of at-risk individuals.

We have modeled the impacts of both common GWAS-associated and rare *LDLR* coding variants through base editing installation followed by cellular uptake of fluorescent LDL-C in HepG2 cells, which provides a scalable flow cytometric assay to measure a key contributing factor of serum LDL-C levels^37^ given the majority of serum LDL-C is cleared in liver^38^. By applying our experimental-computational pipeline to this screen model, we identify LDL uptake-altering GWAS-associated variants and characterize their downstream impact on chromatin accessibility, transcription factor binding, and gene expression that leads to differential LDL uptake. We nominate causal variants that alter LDL-C uptake through impacting the genes *OPRL1*, *VTN*, and *ZNF329,* which have not previously been connected with LDL-C levels.

Through saturation tiled base editing of *LDLR*, not only do we accurately distinguish known pathogenic vs. benign variants, we find strong correlation between missense variant functional scores and the LDL-C levels of patients in the UKB who carry these variants. We combine functional scores with structural modeling to mechanistically classify deleterious variant impacts, revealing a key, conserved tyrosine residue in each LDLR class B repeat that interacts with the neighboring repeat to maintain structural integrity. Altogether, BEAN provides a widely applicable tool to characterize single-nucleotide variant functions.

## Results

### A base editing reporter profiles endogenous editing outcomes

To enable accurate interrogation of variant effects at scale, we built a platform to perform dense, high-coverage base editing screens that accounts for variable editing efficiency and genotypic outcomes. To maximize coverage of variants in base editing screens, we built lentiviral adenine (ABE8e)^11,39^ and cytosine (AID-BE5)^29^ deaminase base editor (BE) constructs using the near-PAM-less SpCas9 variant, SpRY^40^. Both BEs showed native genomic editing activity, as measured in HepG2 cells by ASGR1 splice site editing followed by flow cytometric anti-ASGR1 antibody staining, with ABE8e-SpRY showing considerably more robust maximal activity (**Supplementary Fig. 1a**). Editing efficiency was increased by 5-10% by prior lentiviral integration of constitutively expressed BEs and by transient dosing of cells with the histone deacetylase valproic acid immediately after BE and gRNA transduction (**Supplementary Fig. 1b-c**), and thus these treatments were implemented in all screens.

Base editing efficiency is known to vary depending on Cas9 binding efficiency as well as the local sequence and chromatin context surrounding the target base^29,41,42^, and thus we expected gRNAs to vary substantially in editing efficiency across target sites. To account for this variability, we synthesized and cloned each gRNA paired with a 32-nt reporter sequence comprising the genomic target sequence of that gRNA into lentiviral base editor vectors (**Fig. 1a**, **Methods**), akin to previously published CRISPR mutational outcome reporter constructs^20,21^. When introduced into cells, the gRNA can edit both its native genomic target site and the adjacent target site (reporter) in the lentiviral vector, which can be read out using next-generation sequencing (NGS).

**Figure 1.**
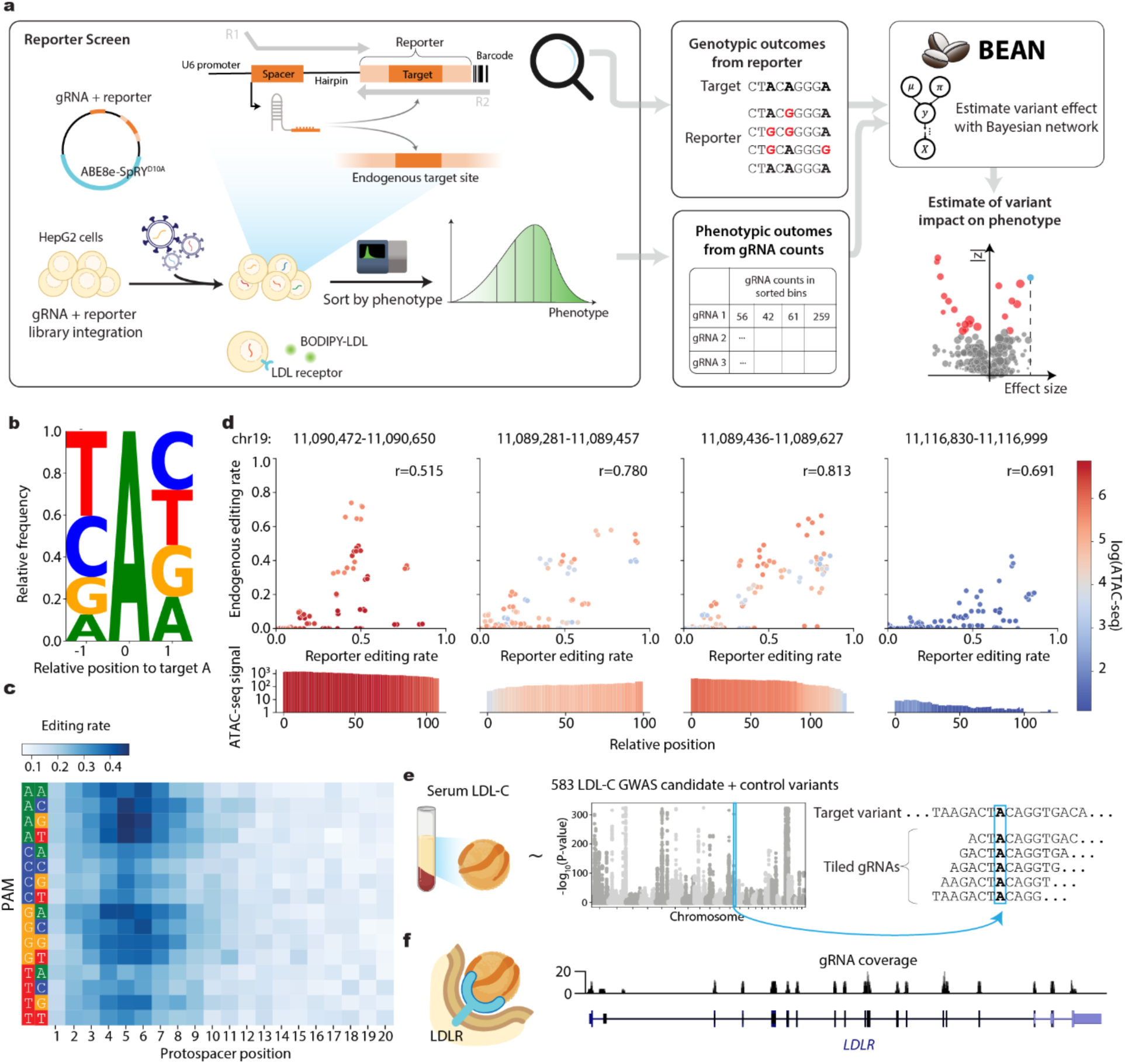
Activity-normalized base editing screening pipeline. **a)** Schematic of activity-normalized base editing screening process and analysis by BEAN. A library of gRNAs, each paired with a reporter sequence encompassing its genomic target sequence, is cloned into a lentiviral base editor expression vector. Lentiviral transduction is performed in HepG2, followed by flow cytometric sorting of four populations based on fluorescent LDL-cholesterol (BODIPY-LDL) uptake. The gRNA and reporter sequences are read out by paired-end NGS to obtain gRNA counts and reporter editing outcomes in each flow cytometric bin. BEAN models the reporter editing frequency and allelic outcomes and gRNA enrichments among flow cytometric bins using BEAN to estimate variant phenotypic effect sizes. **b)** Adjacent nucleotide specificity of ABE8e-SpRY editing represented as a sequence logo from 7,320 gRNAs; the height of each base represents the relative frequency of observing each base given an edit at position 0. **c)** Average editing efficiency of ABE8e-SpRY by protospacer position and PAM sequence **d)** Scatterplots comparing nucleotide-level editing efficiency between the reporter and endogenous target sites for a total of 49 gRNAs across four loci across 3 experimental replicates. The accessibility of the four loci as measured by ATAC-seq signal in HepG2 is shown in the top panel, and the scatterplot markers are colored by the accessibility of each nucleotide. Pearson correlation coefficients are shown as *r*. **e)** Schematic of the LDL-C variant library gRNA design for selected GWAS candidate variants with a Manhattan plot showing variant P-values from a recent GWAS study^50^. gRNAs tile the variant at five positions with maximal editing efficiency (protospacer positions 4-8). **f)** gRNA coverage of the LDLR tiling library across LDLR coding sequence along with 5’ and 3’ UTRs and several regulatory regions.

We designed two gRNA libraries using this approach to improve understanding of the genetics of LDL-C levels. The first library (LDL-C GWAS library) targets 583 variants associated with LDL-C levels from the UK Biobank GWAS cohort (**Methods**, **Supplementary Table 1**). We included fine-mapped variants with posterior inclusion probability (PIP) > 0.25 from either the SUSIE or Polyfun fine-mapping pipelines^43,44^, and also variants with PIP > 0.1 within 250 kb of any of 490 genes found to significantly alter LDL-C uptake from recent CRISPR-Cas9 knockout screens^37,45^. We designed five tiled gRNAs for each variant allele that place the variant in positions shown to induce most efficient editing with ABE8e (**Fig. 1e**)^46^. Positive control gRNAs which ablate splice donor and acceptor consensus sites in six genes found to have significantly altered LDL-C uptake upon knockout^37^, and 100 non-targeting negative control gRNAs that tile 20 synthetic variants were included, for a total of 3,455 gRNAs.

The second library (*LDLR* tiling library) targeted the *LDLR* gene (**Supplementary Table 2**). Taking advantage of the flexible PAM recognition of SpRY, every possible gRNA targeting the *LDLR* coding sequence on both strands was included. Lower density gRNAs, tiled every 2-3-nt, targeted the the 50-nt flanking each *LDLR* exon, the *LDLR* 5’ and 3’ UTR, promoter, and two intronic enhancers (**Fig. 1f**). This library also contained 150 non-targeting negative control gRNAs, for a total of 7,500 gRNAs.

We first assessed editing outcomes through lentiviral transduction of each library in HepG2 cells followed by NGS of gRNA-reporter pairs 10-14 days afterwards. We developed an end-to-end computational toolkit for base-editing screens, BEAN, which includes the ability to perform quality control and quantify editing outcomes from raw reads among other functionalities. Importantly, the quantification step is designed to account for self-editing of the spacer sequence, which we found to occur at appreciable frequency and with modest correlation with reporter editing frequency (LDL-C GWAS library median 31%, Pearson r=0.36, LDLR tiling library median 18%, r=0.31, **Supplementary Fig. 2**). We used BEAN to profile the previously uncharacterized PAM-less base editors ABE8e-SpRY and AID-BE5-SpRY on reporter data from the >10,000 gRNAs in both libraries (**Fig. 1b-c**, **Supplementary Fig. 3**). The result clearly recapitulated the hallmark positional preferences of these base editors^5,9^ the NRY PAM preference of the SpRY enzyme^10,11^, and the relative depletion of editing at AA dinucleotides by ABE8e. Notably, the average maximal positional ABE8e-SpRY editing frequency at protospacer positions 3-8 across dinucleotide PAM sequences ranges from 32% to 46%, indicating the ability of this enzyme to install variants efficiently across a wide variety of genomic locations.

To validate that editing of the reporter provides an accurate surrogate for endogenous editing, we generated a library where both the reporter and endogenous target site are sequenced following the editing by 49 gRNAs across four loci surrounding *LDLR* with varying levels of HepG2 chromatin accessibility (**Supplementary Table 3**). We demonstrate that nucleotide-level and allele-level reporter editing fractions correlate well with endogenous target site editing fractions (**Fig. 1d**, **Supplementary Fig. 4**, average Pearson correlation across 4 loci is r=0.70 for per-nucleotide editing rate r=0.70, per-allele editing rate r=0.69), and the reporter shows higher correspondence than BE-Hive predictions^29^ (Nucleotide r=0.44, allele r=0.64) (**Supplementary Fig. 5**). Notably, while reporter editing correlates with endogenous editing at all four loci, we found that endogenous editing frequency also depends on the accessibility of the target region, as has been previously reported for Cas9-nuclease^47–49^ and base editors^41,42^. Yet, current computational analyses do not model these dependencies, motivating the development of a tailored modeling framework.

We then performed fluorescent LDL uptake screens with each library in ≥5 biological replicates, ensuring >500 cells per gRNA at all stages. We used simulation to determine the optimal flow cytometric sorting scheme, accounting for variability in gRNA editing rate, gRNA coverage, gDNA sampling and PCR amplification (https://github.com/pinellolab/screen-simulation/). Based on our simulation result that finer bin widths improves sensitivity (**Supplementary Fig. 6**, **Supplementary Note 1**), we flow cytometrically isolated four populations per replicate with the very low (0-20% percentile), low (20-40%), high (60-80%), and very high (80-100%) LDL uptake (**Fig. 1a**), performing NGS on gRNA and reporter pairs in each sorted population. We observed robust replicability (median Spearman 𝜌=0.84 for LDL-C GWAS library, 0.88 for *LDLR* tiling library) in gRNA counts across replicates (**Supplementary Fig. 7**), indicating technical reproducibility.

### Activity-normalized base editing screen analysis with BEAN

We postulated that the gRNA editing outcomes provided by the reporter together with the accessibility of the target region could improve the quantification of variant phenotypic effects in our pooled BE screens. To do so, we developed a novel analysis method, BEAN (Base Editor screen analysis with Activity Normalization), to quantify the effect of each variant from gRNA abundance in sorted populations along with genotypic outcome information provided by reporter editing. BEAN assumes that the observed phenotypic distribution in a population of cells for each gRNA derives from a mixture of cells with unedited and edited alleles (Fig. 2). The proportion of cells carrying a given gRNA that possess a particular genotype is inferred based on the editing outcome observed in reporter as well as chromatin accessibility of the target locus using a Bayesian network. The distribution of cells with each gRNA prior to sorting is modeled as a Gaussian mixture for each underlying genotype produced by that gRNA. Because multiple gRNAs may induce the same genotypic outcome at different frequencies, BEAN uses this redundancy to build confidence in the predicted phenotypic impacts of a given genotype. As the output for each variant, BEAN provides its effect size i.e. the posterior mean phenotypic shift along with the corresponding z-score, and 95% credible interval (CI). We also note that BEAN can be adapted to an arbitrary number and arrangement of sorting bins and other base editing enzymes including those with uncharacterized editing preferences, and can accommodate screens without reporter or accessibility information (**Methods**).

**Figure 2.**
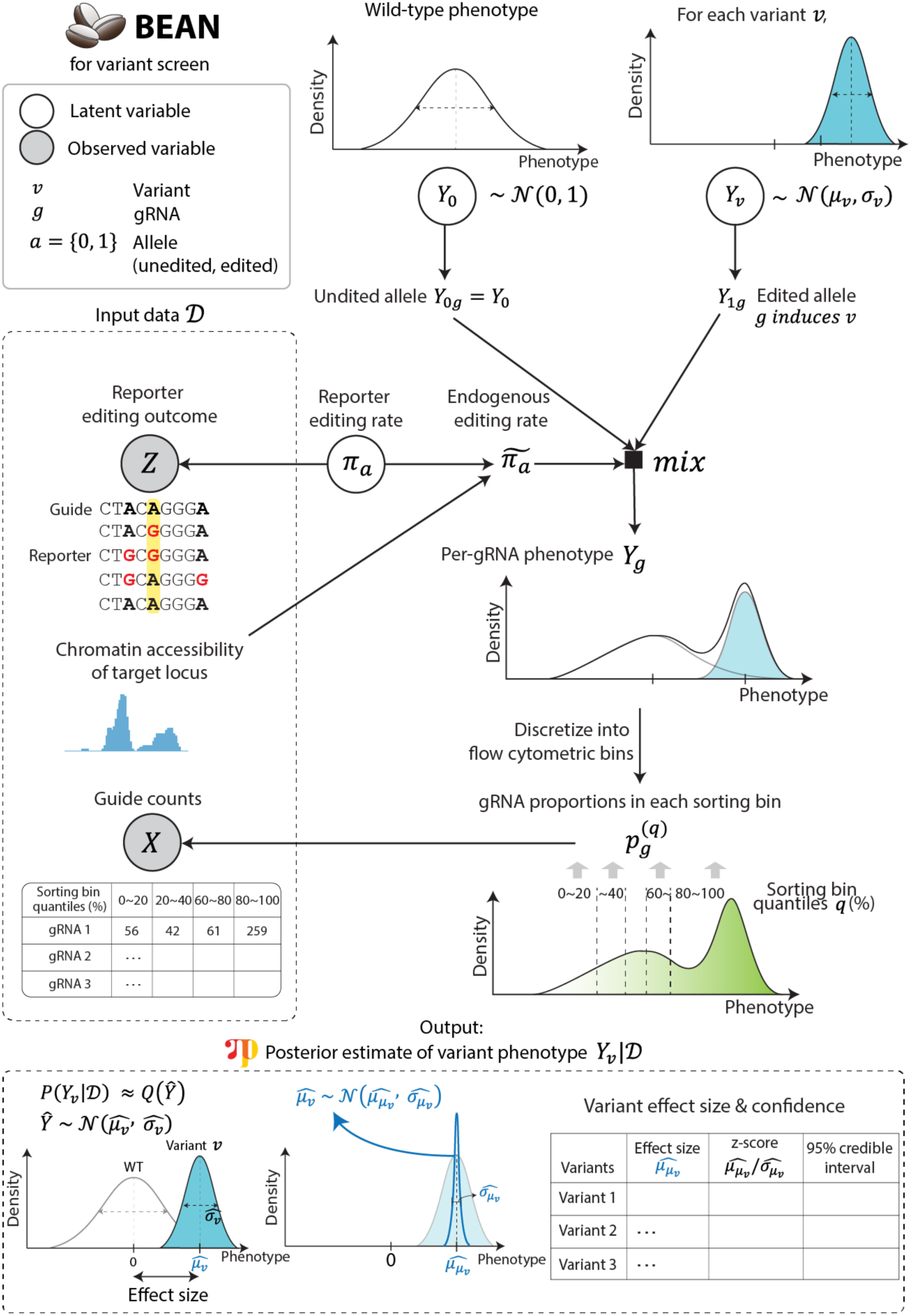
BEAN models variant effects from activity-normalized base editing screens. Simplified schematic of BEAN Bayesian network that models input reporter editing outcomes and gRNA counts. The Bayesian network model recapitulates the data generation process starting from a variant-level phenotype Y_v_ and models per-gRNA phenotypes as a Gaussian mixture distribution of edited and unedited (wild-type) allele phenotypes. The weights of the mixture components are modeled to generate reporter editing outcomes. gRNA abundance in each sorting bin is then calculated by discretizing the gRNA phenotype based on the experimental design into the phenotypic quantiles, and is modeled to generate the observed gRNA counts using an overdispersed multivariate count distribution (see Methods). BEAN outputs the parameters of the posterior distribution of mean phenotypic shift as Gaussian distribution with mean 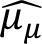 (effect size), along with negative-control adjusted z-score and credible interval (CI), where 𝒟 is the input data.

BEAN only assumes population-level consistency between editing of the reporter and endogenous target site. We hypothesized that variation in editor expression or cellular state may lead certain cells to be more amenable to editing than others. In this scenario, “jackpot” cells would be more likely to have editing at both endogenous and reporter loci. To assess this possibility, we compared the enrichment of a gRNA in the highest vs. lowest sorted LDL uptake quantile bin with the difference in reporter editing observed in cells sorted into these bins, reasoning that endogenous editing should be highest in the cells sorted into the enriched bin. We indeed observed such correlation for *LDLR* and *MYLIP* splice-ablating gRNAs (Spearman ρ=0.32, **Supplementary Fig. 8**), suggesting the existence of cell-level factors leading to “jackpot” cells with higher editing at both endogenous and reporter loci. However, the correlation between phenotypic and reporter editing enrichment was weaker when considering all positive control gRNAs (Spearman ρ=0.13). We thus concluded that incorporating the jackpot effect into BEAN would be unlikely to improve model performance.

### BEAN identifies LDL uptake altering GWAS variants

We applied BEAN to the LDL-C GWAS library screen. From the reporter data, variant editing efficiency per gRNA is highly variable with average edit fraction of 34.0%. Encouragingly, most target variants are edited at high efficiency by at least one of the five targeting gRNAs (median maximal editing of 60.4%, **Supplementary Fig. 9**).

First, we compared the performance of BEAN and five published CRISPR screen analysis methods at distinguishing the effects of positive control splice-altering variants versus negative control non-targeting gRNAs^23,51–54^ (**Methods,** Fig. 3a). To dissect the contributions of individual features to BEAN performance, we included two reduced versions of BEAN: one that considers reporter editing but not chromatin accessibility (BEAN-Reporter), and another that ignores the reporter, assuming uniform gRNA editing efficiency (BEAN-Uniform) (**Methods, Supplementary Fig. 10**). BEAN outperforms other evaluated methods at this classification task (Fig. 3b, **Supplementary Fig. 11**), and this improved performance is accentuated when the data is subsampled for fewer replicates, demonstrating its ability to maintain robustness even with less data. Importantly, BEAN shows improved performance (mean AUPRC=0.90 across 15 2-replicate subsamples) over BEAN-Reporter (mean AUPRC=0.87), which in turn outperforms BEAN-Uniform (mean AUPRC=0.85), supporting the value of accurately modeling target site editing. Intriguingly, even BEAN-Uniform outperforms alternative approaches, likely due to more accurate modeling of sorting bins, suggesting the utility of BEAN in sorting screens without reporter.

**Figure 3.**
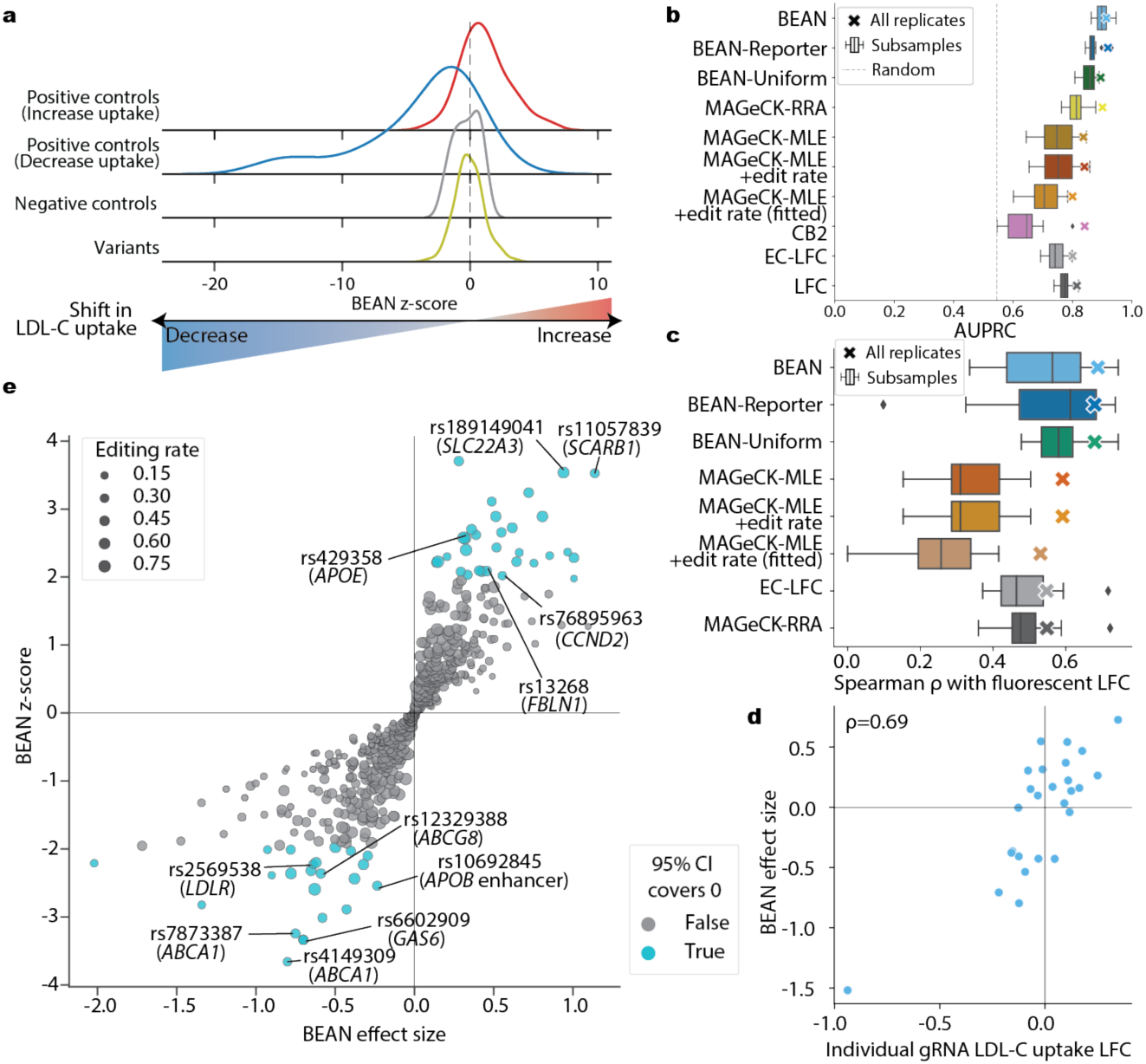
BEAN improves variant impact estimation from the LDL-C GWAS library screen. **a)** Ridge plot of BEAN z-score distributions of positive controls, negative controls, and test variants. **b)** AUPRC of classifying *LDLR* and *MYLIP* splicing variants vs. negative controls. Metrics for all 5 replicates are shown as markers and metrics of 15 two-replicate subsamples among the 5 replicates are shown as box plots. **c)** Spearman correlation coefficient between BEAN effect size and log fold change of LDL-C uptake following individual testing of 26 gRNAs. Metrics for all 5 replicates are shown as marker and metrics of 15 two-replicate subsamples among the 5 replicates are shown as box plots. **d)** Scatterplot of BEAN effect size and log fold change of LDL-C uptake following individual testing of 26 gRNAs. Spearman correlation coefficient is denoted as ρ. **e)** Scatterplot of variant effect size and significance estimated by BEAN. Labels show rsIDs of selected variants and dbSNP gene annotations and a manual annotation for APOB enhancer in the parenthesis.

Having demonstrated robust performance of BEAN, we evaluated our ability to characterize common variants that alter LDL-C uptake. We identified 54 variants that significantly alter LDL-C uptake (95% CI does not contain 0, **Supplementary Table 4**). These variants include intronic variants in well-known LDL-C uptake mediators whose knockout altered LDL-C uptake in a recent genome-scale CRISPR screen^37^ such as *ABCA1*, *LDLR*, and *SCARB1* (Fig. 3e). We additionally identified coding/intronic variants in *APOE*, *CCND2*, *GAS6*, and *FBLN1* with strong genetic likelihood of causality (UKBB SUSIE fine-mapping PIP > 0.99 and/or the only variant in a fine-mapped credible set^30^), indicating that the effect of these variants on serum LDL-C is at least partially mediated by LDL-C uptake.

To validate the inferred effect sizes, we performed individual lentiviral ABE8e-SpRY transduction of HepG2 cells with gRNAs targeting 22 variants and 4 positive controls (**Supplementary Table 5**). We performed fluorescent LDL-C uptake profiling of each edited cell line mixed with an in-well control cell line in 6 biological replicates (**Methods**), allowing us to compare changes in LDL-C uptake with matched data from the screen. The individual LDL-C uptake log-fold-change (LFC) values showed strong correlation to the BEAN effect sizes (𝜇, Spearman R=0.69, Fig. 3c**-d**, **Supplementary Fig. 12**), showing more robust correlation than BEAN-Uniform (R=0.68), log fold change based on MAGeCK-RRA (R=0.51), and regression coefficients 𝛽 of MAGeCK-MLE (R=0.61, see **Methods**). These data demonstrate that BEAN enables accurate inference of variant effects on LDL-C uptake over a wide dynamic range.

To gain insight into a set of 20 variants for which the mechanism of LDL-C uptake alteration is less clear, we developed a pipeline to assess the cellular effects of variant installation (Fig. 4a). First, we asked which of these variants impact chromatin accessibility. We established an approach to perform pooled variant editing followed by ATAC-seq. High multiplicity of infection (MOI) lentiviral delivery of a pool of 20 ABE8e-SpRY gRNAs to HepG2 cells was followed by ATAC-seq and paired genomic DNA collection in three biological replicates in standard and serum-starved conditions. We performed multiplexed PCR enrichment of the regions surrounding each of the 20 edited variants followed by targeted amplicon sequencing by NGS. Differential representation of an alternate allele in ATAC-seq relative to gDNA sequencing implies differential accessibility of the alternate allele than the reference (Fig. 4b).

**Figure 4.**
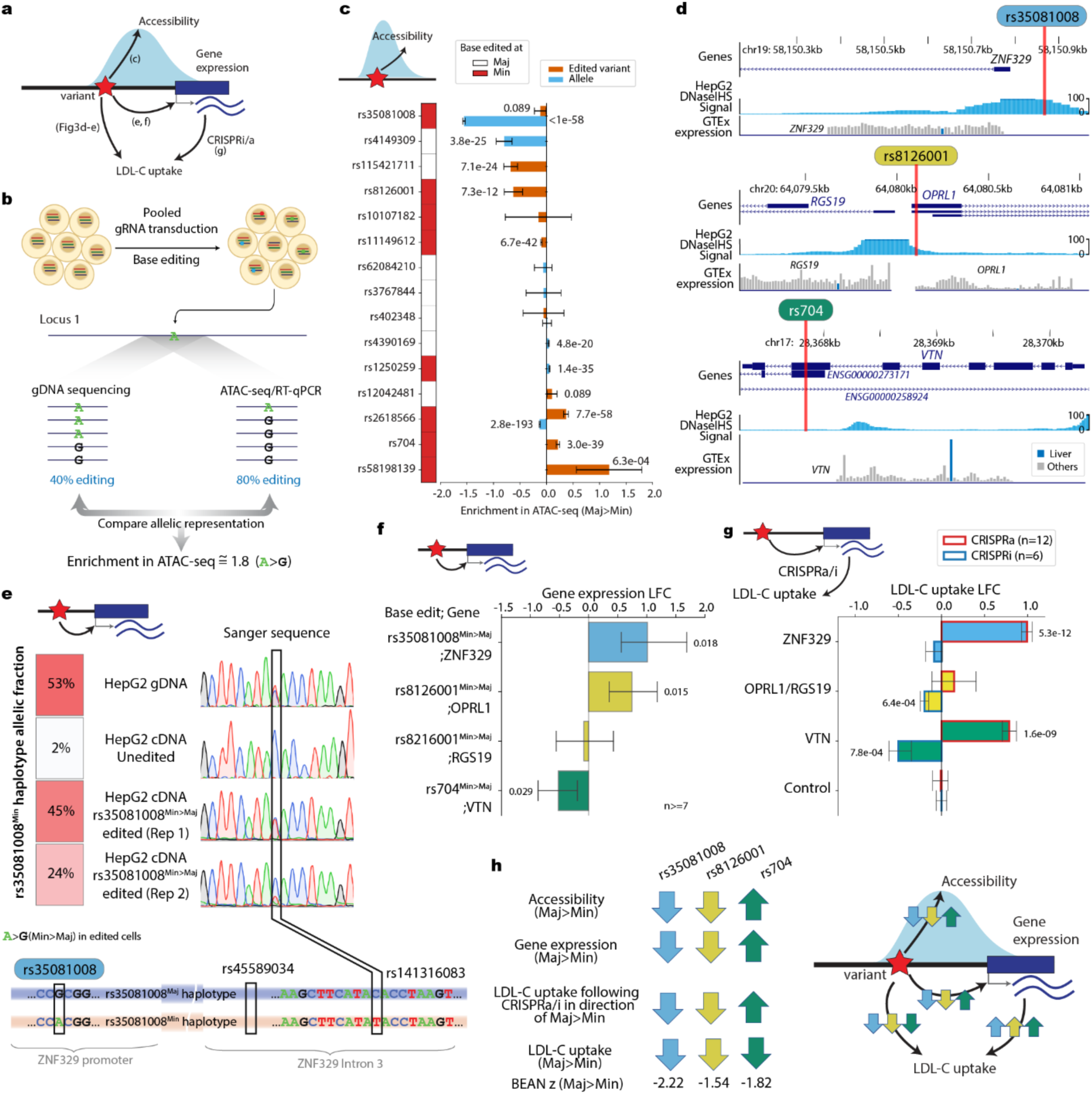
Functional characterization of LDL-C GWAS variants. **a)** Schematic of potential variant mechanisms and the figure panels showing data from each mechanistic experiment. **b)** Schematic of pooled ATAC-seq analysis to identify variants impacting accessibility. Differential representation of allele in gDNA and ATAC-seq reflects differential accessibility induced by the base edit or heterozygous reference allele. **c)** Change in ATAC-seq enrichment from the pooled ATAC-seq experiment. 95% confidence intervals are shown as the error bars. “Edited variant” denotes the enrichment by base edit and “Allele” denotes the enrichment by either of the heterozygous alleles, when available, translated uniformly to major (Maj) to minor (Min) allele direction. Variants where the base edit is conducted from minor allele are denoted as red in the color bar. Family-wise error rate (FWER) with Benjamini-Hochberg multiple correction is shown for each enrichment value where FWER < 0.1. **d)** Genomic tracks for three selected variants. DNaseIHS; DNase 1 Hypersensitivity. Multiple transcript variants of RGS19 and OPRL1 are shown in the middle panel. **e)** Fraction of *ZNF329* minor (Min) allele haplotype reads in gDNA and cDNA from untreated HepG2 and HepG2 with rs35081008^Min>Maj^ base editing. **f)** Change in gene expression following base editing of three selected variants from minor (Min) to major (Maj) allele. P-values of the one sample Student’s t-test of LFC vs. mean of 0 that are smaller than 0.05 are shown above each bar. **g)** Change in cellular LDL-C uptake following CRISPRa/i of proximal genes for three selected variants. P-values of the one sample Student’s t-test of LFC vs. mean of 0 that are smaller than 0.05 are shown above each bar. h) Summary schematic of characterization results.

Eight of the 20 variants are heterozygous in HepG2, and thus we could assess whether these variants reside in chromatin accessibility quantitative trait loci (caQTL)^55^, showing differential relative accessibility of the two haplotypes irrespective of base editing. We found five of these eight variants to be caQTLs (Fig. 4c). Two of these loci (rs35081008 and rs2618566) were also identified as caQTLs in a recent analysis of 20 human liver tissue samples^56^. Importantly, caQTL analysis cannot address the causality of the evaluated variant due to the presence of linked variants which could contribute to the differential ATAC-seq signal.

To assess whether individual variants alter chromatin accessibility, we evaluated whether base editing induces differential accessibility for any of the 20 tested variants. Technical issues including insufficient representation of the region, insufficient editing, and inability to phase heterozygous loci prevented assessment of five of the variants (see **Methods**). Of the 15 remaining variants, eight significantly altered chromatin accessibility when edited (family-wise error rate 0.1, Fig. 4c). Four such variants (rs11149612, rs35081008, rs8126001, and rs2618566) were in loci identified as liver tissue caQTLs. Because base editing only alters a single variant in a locus, this analysis establishes at least partial causality to the tested variant.

We performed deeper characterization of three variants whose editing alters both LDL-C uptake and chromatin accessibility. Rs704 is a missense coding variant in *VTN* and is the only variant in a fine-mapped credible set from LDL-C GWAS^30^, suggesting high likelihood of causality. The other two variants are in gene promoters—rs35081008 is in the *ZNF329* promoter, and rs8126001 is in the shared *OPRL1/RGS19* promoter (Fig. 4d). Both variants have moderate probability of causality from GWAS evidence (SUSIE PIP=0.49 for rs35081008, PIP=0.25 for rs8126001), with the remaining probability in the rs35081008 locus deriving from a linked variant (rs34003091) 19-nt upstream in the *ZNF329* promoter. None of the putative target genes has been previously found to alter LDL-C uptake, nor do they show significance in LDL-C burden analyses.

To investigate how the prioritized variants might affect transcription factors (TF) binding sites and thereby regulate proximal genes involved in LDL-C uptake, we adapted the MotifRaptor pipeline^57^. Briefly, MotifRaptor predicts both the binding strength and disruption scores of TF motifs at specified SNP loci (see **Methods**). Using the human TFs from the CIS-BP motif database^58^, for each variant, we ranked the TFs with high binding and potential disruption by the variant (**Supplementary Table 6**). For rs8126001, our approach prioritized two zinc finger TFs, ZNF333 and ZNF770 with enhanced binding site sequences due to the heterozygous minor allele in HepG2 cells (**Supplementary Fig. 13**). HepG2 ChIP-seq data^59^ further support the binding of these TFs at this locus, although the variant lies at the edge of the peaks (**Supplementary Fig. 14**). While definitive conclusions about these factors will require further experimental validation, our observations align with previous research^60^ suggesting that only a minority of causal variants directly alter canonical TF binding sequences and instead affect non-canonical sequences.

We confirmed through RT-qPCR analysis that editing the minor to major alleles of rs35081008 and rs8126001 leads to increased expression of *ZNF329* and *OPRL1* respectively (Fig. 4f), which is consistent with the increased chromatin accessibility induced by these edits. rs35081008 is heterozygous in HepG2, and we used two linked *ZNF329* intronic variants to assess allele-specific expression. In wild-type HepG2, only 2% of *ZNF329* transcripts derive from the minor allele haplotype (Fig. 4e), consistent with the diminished chromatin accessibility of this allele (Fig. 4c) and the status of rs35081008 as a liver eQTL. Editing rs35081008 from minor to major allele restores expression of this haplotype to 35% of total transcripts (Fig. 4e), providing further evidence that rs35081008^Maj^ results in increased expression of *ZNF329*.

We then performed CRISPRa and CRISPRi targeting to assess whether altered expression of the four candidate target genes alters LDL-C uptake. CRISPRa induction of *VTN* and *ZNF329* significantly increased LDL-C uptake, and CRISPRi repression of *VTN* and *OPRL1/RGS19* reduced LDL-C uptake (Fig. 4g). In our base editing experiments, rs704^Min^ shows decreased LDL-C uptake, so we surmise that this allele must have decreased expression or function, given that decreased *VTN* expression decreases LDL-C uptake (Fig. 4h). Prior biochemical characterization has shown decreased cellular binding capacity of rs704^Min^, suggesting a possible mechanistic explanation. Our data are consistent with rs35081008^Min^ decreasing *ZNF329* expression, which in turn decreases LDL-C uptake. Finally, our data are most consistent with rs8126001^Min^ decreasing *OPRL1* expression, which leads to decreased LDL-C uptake. This observation aligns with the higher predictive binding affinity of ZNF333, a transcriptional repressor^61,62^, with rs8126001^Min^ and the potential disruption of its binding with rs8126001^Maj^. In summary, through accurately quantifying impacts of disease-associated variants on LDL-C uptake, BEAN reveals genetic mechanisms underlying control of LDL-C levels.

### Saturation *LDLR* coding sequence tiling screening enables quantitative assessment of rare variant deleteriousness

We next adapted BEAN to the *LDLR* tiling library, enhancing the model to specifically assess the contributions of individual amino acid mutations rather than SNVs, by enabling a more comprehensive understanding of coding region alterations. Previous coding sequence base editing analyses have assumed that all editable bases within a window are edited, which leads to erroneous amino acid mutation assignments, or have analyzed gRNA-level signal only^15^. We aimed to exploit the combination of dense tiling afforded by ABE8e-SpRY and reporter editing outcomes to model the effects of coding variants more accurately.

The *LDLR* tiling screen showed high coverage of edited nucleotides and amino acids (92% of targetable nucleotides and 74% of the 860 *LDLR* amino acids in the *LDLR* coding sequence were edited at >10% frequency by at least one gRNA in the reporter, **Supplementary Fig. 15**). A total of 2,182 distinct variants were assessed, of which 874 are missense coding variants. Each gRNA produced an average of 2.6 distinct alleles, and each variant was covered by 5.8 gRNAs on average (**Supplementary Fig. 16**). Thus, ABE8e-SpRY tiling of *LDLR* resulted in a rich dataset of coding variants for the evaluation of their phenotypic impacts.

As opposed to the LDL-C GWAS analysis in which each gRNA was evaluated based on its editing frequency at a single target position, we adapted BEAN to account for multi-allelic outcomes. First, BEAN translates the edited alleles, i.e., aggregates nucleotide-level allele counts that leads to the identical amino acid transition into a single amino-acid level allele counts, while preserving nucleotide transition in non-coding regions. BEAN then filters for the translated alleles that are robustly observed (see **Methods**) for each gRNA (Fig. 5a). BEAN uses a Bayesian network to combine phenotypic information from all the gRNAs that produce a given allele. Importantly, the phenotype attributed to each gRNA is modeled as a mixture distribution of the alleles it generates, with the contribution of each allele weighted by its corresponding editing frequency.

**Figure 5.**
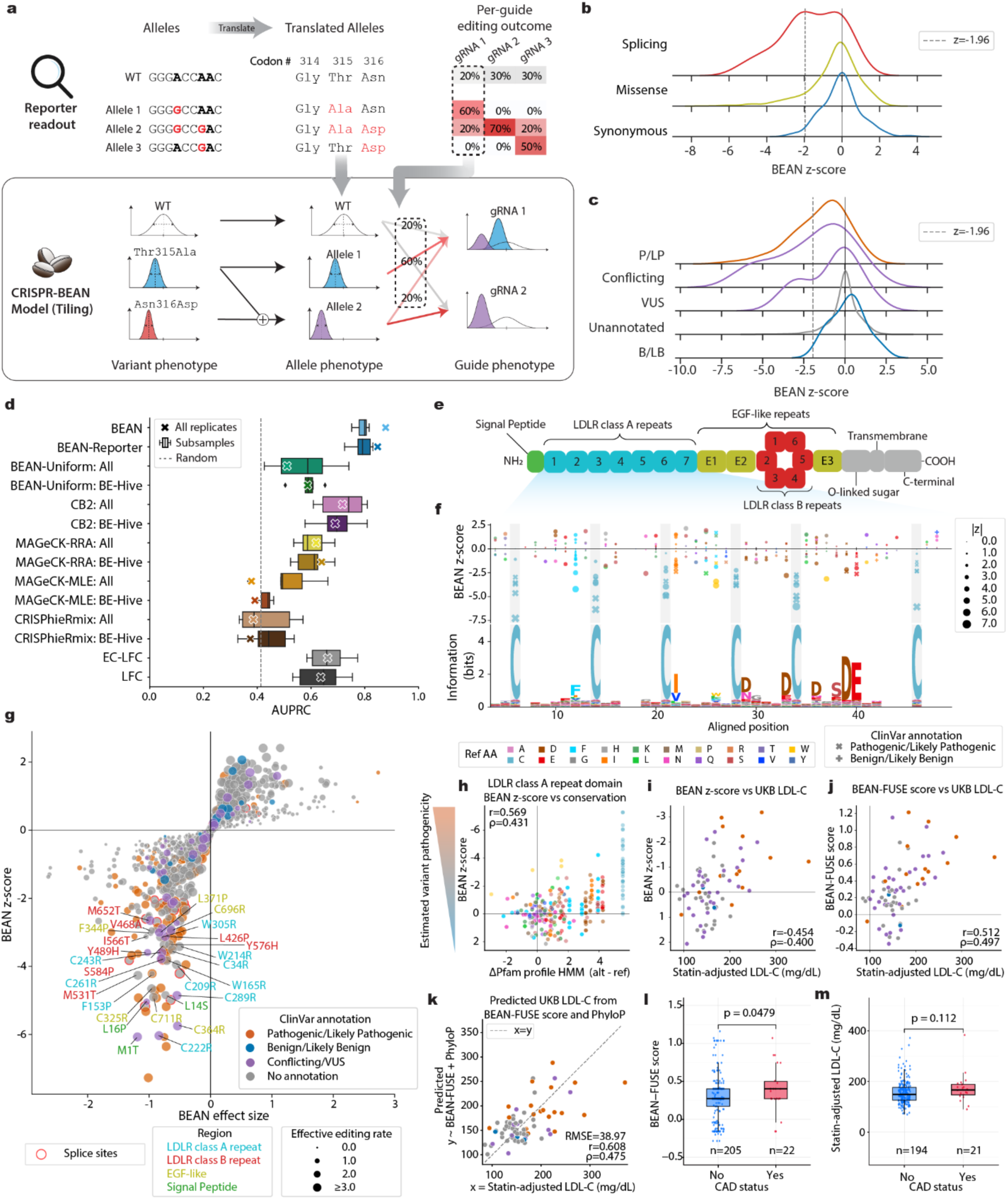
Dissection of *LDLR* variant effects through BEAN modeling of a saturation tiled base editing screen. **a)** BEAN model for coding sequence tiling screens. Reporter editing efficiencies are calculated at the amino acid-level when the edited nucleotides are in coding region. Phenotypes of gRNAs with multi-allelic outcomes are modeled as the Gaussian mixture of allelic phenotypes. If an allele consists of more than one variant, the phenotype of the allele is modeled as the sum of the component variants. A Bayesian network is used to model variant-level phenotypes, sharing phenotypic information from all available gRNAs. **b)** Ridge plot of BEAN z-score distributions of positive controls, negative controls, and variants. **c)** Ridge plot of BEAN z-score distributions of Clinvar variants annotated as pathogenic/likely pathogenic (P/LP), benign/likely benign (B/LB), conflicting interpretation of pathogenicity (conflicting), and Uncertain significance (VUS), and unannotated variants. **d)** AUPRC of classifying ClinVar pathogenic/likely pathogenic vs. benign/likely benign variants. The marker shows the metrics of each method run on 4 replicates with no failing samples. Boxplot shows the metrics of 6 2-replicate combinations of the 4 replicates. **e)** *LDLR* domain structure adopted from Oommen et al^71^. **f)** BEAN z-scores for variants in the 7 LDLR class A repeat domains aligned with the Pfam profile HMM logo. Highly conserved cysteines are highlighted in grey. **g)** Scatterplot of estimated variant effect sizes and z-scores. Labels of selected deleterious variants without ClinVar pathogenic/likely pathogenic annotations are shown. **h)** Scatterplot of LDLR class A repeat missense variant BEAN z-scores and ΔPfam profile HMM scores. Higher ΔPfam scores correspond to substitution from highly conserved to rarely observed amino acids. **i)** Comparison of mean statin-adjusted LDL-C level and BEAN z-score for variants observed in UKB and base editing. **j)** Comparison of mean statin-adjusted LDL-C level and BEAN-FUSE scores for variants observed in UKB and base editing. **k)** LDL-C levels of observed missense variants predicted by a regression model using BEAN-FUSE and PhyloP scores with 10-fold cross validation, compared with mean statin-adjusted LDL-C level in UKB. **l)** Boxplots of BEAN-FUSE functional scores for UKB individuals with variants observed in our base editing screen with or without CAD. **m)** Boxplots of statin-adjusted LDL-C levels of UKB invididuals with variants observed in our base editing screen with or without CAD. P-value of two-sided Wilcoxon rank-sum test is denoted. *r*; Pearson correlation coefficient, 𝜌; Spearman correlation coefficient

BEAN assigned significant z-scores (<-1.96, equivalent to 95% credible interval not covering 0) to 145 among 2,182 variants assessed from the *LDLR* tiling library (**Supplementary Table 7**), 131 of which decrease LDL-C uptake. 47 variants that significantly decrease LDL-C uptake are annotated in ClinVar as pathogenic/likely pathogenic, while 17 are ClinVar VUS/conflicting variants and none are ClinVar benign/likely benign (Fig. 5g), indicating that BEAN can reliably predict the pathogenicity for variants without a pathogenic or benign classification (Fig. 5b**-c**).

We compared the performance of BEAN at distinguishing ClinVar-annotated pathogenic from benign/likely benign *LDLR* variants to other available screen analysis methods^51–54^. To allow comparison of methods that do not account for editing outcomes, we assigned outcomes to each gRNA either by assuming all editable bases within the maximal editing window are perfectly edited^13^ (“All”) or by using the most frequent predicted outcome from BE-Hive^29^(“BE-Hive”). As in the LDL-C GWAS screen, BEAN showed better performance than any other method (Fig. 5d, **Supplementary Fig. 17**), and BEAN also outperformed the model variants that do not account for accessibility (BEAN-Reporter) or reporter editing outcomes (BEAN-Uniform), further justifying the modeling decision to explicitly leverage editing outcomes and accessibility. BEAN achieves an AUPRC of 0.88 at this task, indicating highly effective distinction of pathogenic and benign *LDLR* variants through scoring HepG2 LDL-C uptake proficiency.

To gain insight into mechanisms by which these variants disrupt LDL-C uptake, we examined BEAN z-scores for variants that reside within conserved functional domains (Fig. 5e**-f**, **Supplementary Fig. 18**). LDLR contains seven highly conserved LDLR class A repeats that bind to LDL. The LDLR class A repeat is structurally anchored by six highly conserved cysteines that form three disulfide bonds^63^. As expected, many of the missense variants with the strongest effects on LDLR function disrupt these cysteines (Fig. 5f). We find that cysteine mutating edits in each of the seven LDLR class A repeats disrupt LDLR activity (**Supplementary Fig. 19**), suggesting that structural integrity of all repeats is required for efficient LDL binding, although disruption is most impactful in repeats 3-7. Truncation experiments have reported that repeats 1 and 2 are dispensable for LDL binding^64^, in partial accord with our results^65^. To examine the relationship between conservation and function more comprehensively in these repeats, we compared the BEAN z-score of every installed variant with its change in amino acid conservation score from the Pfam profile HMM^66^ (see **Methods**). We observed strong concordance (Pearson r = 0.57 Fig. 5h), with N-terminal hydrophobic residues and C-terminal calcium-coordinating acidic residues within the repeats also showing particular functional importance, as expected from the known function of these domains.

Encouraged by the concordance between our screen and conservation scores within the LDLR class A repeats, we asked whether BEAN scores could predict functional impairment across the entire *LDLR* gene. We examined statin-adjusted LDL-C levels^67^ in the UK Biobank (UKB) for individuals with paired exome sequencing and lipid level data. To control for the contribution of other variants in genes known to impact serum LDL-C level, we filtered out individuals who harbor nonsynonymous *APOB* or *PCSK9* variants or multiple *LDLR* missense variants, leading to 9,819 individuals harboring 358 distinct *LDLR* missense variants. There are 76 distinct *LDLR* missense variants observed in our base editing data with UKB carriers. We observe robust concordance between the average carrier LDL-C and BEAN scores for these variants (Spearman ρ = 0.40, Pearson r = 0.45, Fig. 5i), suggesting that BEAN provides accurate quantitative prediction of the impact of *LDLR* missense variants on control of serum LDL-C levels in the human population.

As our base editing screen does not exhaust possible mutation types per position, we used the FUSE^68^ pipeline to impute the impact of unobserved variants at positions at which a different missense variant is scored. FUSE uses an amino acid substitution matrix derived from 24 deep mutational scanning datasets to impute functional scores for all possible missense variants at positions observed in base editing data (BEAN+FUSE score, see **Methods**). Applying FUSE to the 76 UKB variants with observed base editing data, BEAN-FUSE shows improved correlation with UKB carrier LDL-C (Spearman ρ = 0.50, Pearson r = 0.51, Fig. 5j). BEAN-FUSE correlation with UKB carrier LDL-C was robust but lower at all 358 missense *LDLR* variants with lipid measurements (Spearman ρ = 0.37, Pearson r = 0.35, **Supplementary Fig. 20**a). Altogether, BEAN-FUSE provides a pipeline to extend base editing screening to predict functional impairment for unobserved missense variants, although our data suggest that accuracy does decrease for unobserved variants.

As base editing provides orthogonal functional assessment to conservation, we asked whether the LDL-C levels of UKB variant carriers could be predicted with BEAN-FUSE scores and PhyloP 100way vertebrate conservation scores. Using XGBoost regression^69^, we achieved more robust correlation with UKB carrier LDL-C than either BEAN-FUSE or PhyloP alone at the 76 variants observed in the base editing screen (Spearman ρ = 0.48, Pearson r = 0.61, RMSE=39.0, Fig. 5k, **Supplementary Fig. 20**c) and at 358 variants with BEAN-FUSE score (Spearman ρ = 0.37, Pearson r = 0.31, RMSE=51.1, **Supplementary Fig. 20**b,d). This result demonstrates the potential utility of base editing data to improve quantitative phenotype prediction combined with computational prediction methods.

Individuals with pathogenic FH variants are at higher risk of coronary artery disease (CAD), even after controlling for LDL-C levels^70^. However, the vast majority of rare *LDLR* missense variants lack ClinVar pathogenic/likely pathogenic designations, preventing information about these potentially disease-causing variants from being shared with patients. Therefore, we asked whether CAD incidence within *LDLR* variant carriers could be stratified by functional scores. We found that for individuals with rare *LDLR* variants, functional scores processed by BEAN-FUSE were significantly higher for patients with prevalent or incident CAD (Wilcoxon rank-sum test, p = 0.0479, Fig. 5l). BEAN-FUSE scores provided more robust stratification of individuals with CAD than statin-adjusted LDL-C values for individuals with variants covered in the screen (Wilcoxon rank-sum test, p = 0.112, Fig. 5m). This demonstrates the advantage of quantifying genetic risk, which has a lifelong impact on LDL-C levels, over the snapshot provided by a single LDL-C measurement. Overall, we show that activity-normalized base editing screening can yield accurate quantitative estimation of *LDLR* variant pathogenicity in a large human cohort.

### Structural basis of LDLR missense variants

We further analyzed *LDLR* missense variants identified to significantly impair LDL-C uptake by BEAN to gain insight into mechanisms of their pathogenicity. We first examined variants with top z-scores that are unannotated or annotated as conflicting, or VUS in ClinVar. The top ranked variant, which shows even more significant loss-of-function than splice-ablating variants, alters the start codon, preventing full-length LDLR translation. Other top variants such as C222R, C261R, C289R, and C364R disrupt conserved disulfide bond-forming cysteines in LDLR class A repeats and EGF-like domains. Top-ranked variant in the of the signal peptide L16P substitute hydrophobic leucines with prolines in the transmembrane alpha helix, which is likely to distort the alpha helix^72^ and the neighboring L15P has been shown to reduce LDLR transport to the plasma membrane^73^. Neighboring L14S that also ranks high substitutes hydrophobic leucine with serine in the hydrophobic h-region central to the signal peptide^74^. Additionally, multiple variants disrupt calcium ion binding, which is key to LDLR class A repeat folding^75^ through the conversion of negatively charged amino acids (D/E) to glycine (G), thereby disrupting ionic interactions with side-chain carboxylates and calcium ions (D94G, E101G, E179G, D307G) in LDLR class A repeats (**Supplementary Fig. 22**). We also found that L371P, a VUS, disrupts a calcium ion interaction in the EGF-like domain by breaking the coordinate covalent bond between the calcium ion and the carbonyl group within the L371 main chain due to backbone distortion. Finally, we found that F153P significantly interfered with hydrophobic interactions between the aromatic ring and the attached saccharide on Q182.

We noticed that an appreciable number of deleterious variants that lack ClinVar pathogenic designation reside in the six LDLR class B repeats. The LDLR class B repeats, also known as YWTD repeats, form a propeller-like structure involved in the release of LDL following its endocytosis. To gain insights into unannotated variant impact, focusing on the LDLR class B repeats, we used the full wild-type LDLR structure from the AlphaFold Protein Structure Database^76,77^ and the MODELLER^78^-generated mutant structures to calculate changes in interatomic interactions using Arpeggio^79^. Additionally, we predicted the effects of variants on protein stability (ΔΔ𝐺, negative value indicates destabilization) with DDMut^80^ (**Supplementary Table 8**). We found that the 26 significant LDLR class B variants induce more destabilizing effects, disrupt more hydrophobic interactions, have lower relative solvent accessibility^81^ (0.041 of maximum residue solvent accessibility), and have higher wild-type residue depth as compared to the other observed variants in this region (Fig. 6a**-d**, **Supplementary Fig. 23**). Collectively, these observations strongly indicate that these significant LDLR class B repeat variants are predominantly buried within the protein core where they engage in extensive hydrophobic interactions essential for protein folding. Moreover, we found a conserved interaction across repeat domains in which a tyrosine (aligned position 5 in **Supplementary Fig. 18**b) holds neighboring propeller blades together through interactions with a hydrophobic residue of the neighboring repeat (Fig. 6e**-f**). We identified five of these variant pairs (Y442C with V481A, Y442C with V468A, Y489H with M531T, Y532C with L568P, and Y576H with V618A), where all nine positions have at least one variant that weakens their hydrophobic interaction and has a significant BEAN z-score. Among the top-ranked unannotated or ClinVar VUS and conflicting variants within LDLR class B repeats, the six most significant variants (L426P, Y489H, M531T, I566T, S584P, and Y576H) all disrupt residues that hold the propeller blades together through hydrophobic interactions (Fig. 6g**-i**, **Supplementary Fig. 24**). Further supporting the importance of hydrophobic interactions, the base editing screen installed additional missense variants at positions 531, 566, and 652 that conserve hydrophobicity. In all cases, mutation into hydrophobic residues has less severe impact from the base editing screen and DDMut-predicted destabilization than mutation into non-hydrophobic residues (Fig. 6j). For example, while we find M652T to be highly deleterious (BEAN z=-2.65), we find no functional disruption from the hydrophobicity-conserving M652V variant (BEAN z=+2.06). This analysis is supported clinically, as M652V is designated in ClinVar as “Likely Benign,” and the average UKB carrier LDL-C is below average (106mg/dL). In summary, structural analysis of rare *LDLR* variants identified by BEAN provides a basis for the missense variant impact through affecting structural integrity of LDLR, highlighting a central role for hydrophobic interactions that hold together adjacent beta blades of the LDLR class B repeat domain.

**Figure 6.**
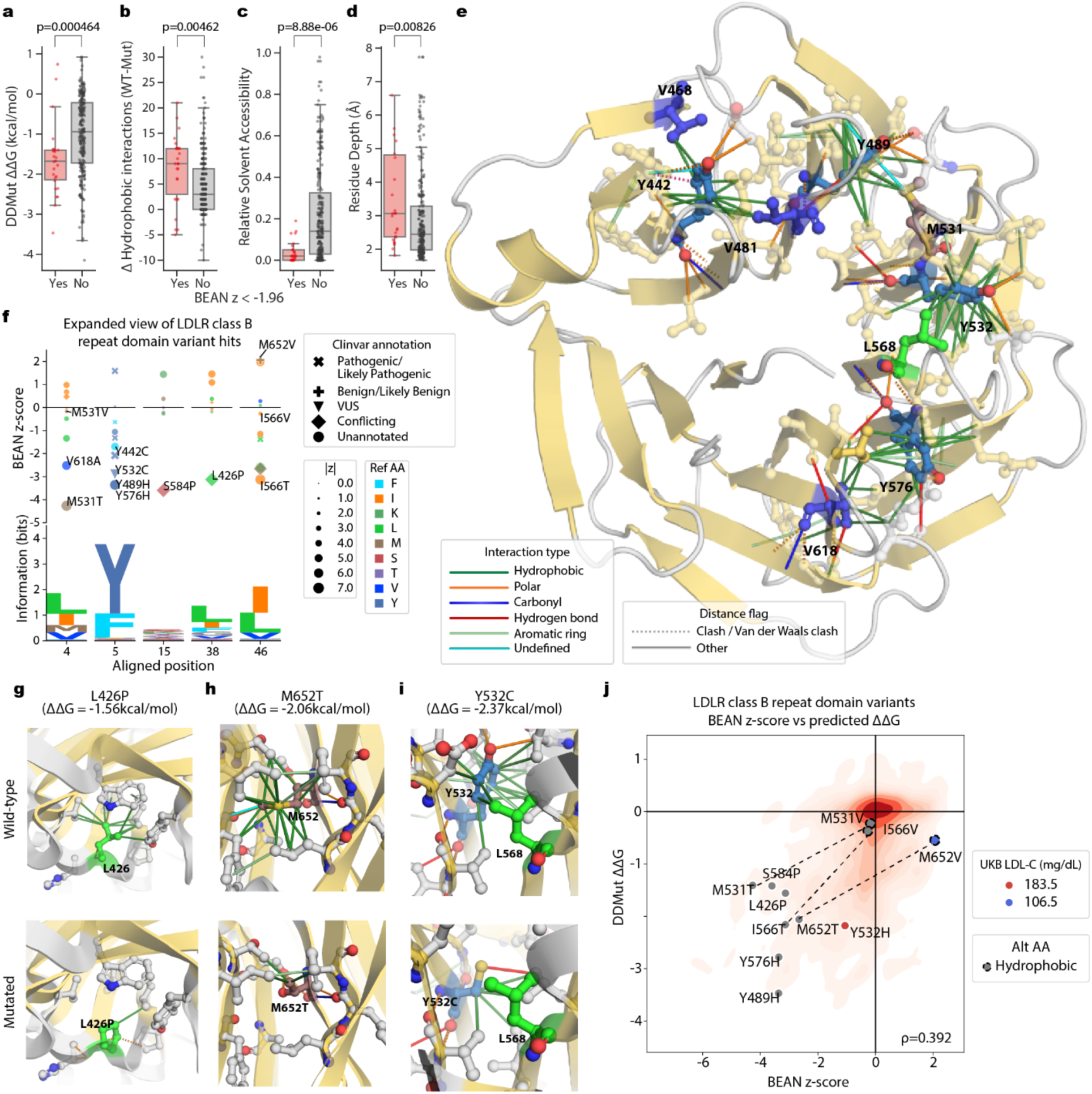
Deleterious variants in LDLR class B repeats weaken hydrophobic interactions. **a-d)** Boxplots of 26 significant (z < -1.96) and the rest of 259 variants observed in LDLR class B repeats. P-values of two-sided Wilcoxon rank-sum test are denoted. WT; wild-type, Mut; mutated. **e)** Conserved interactions involving tyrosine of which mutation showed significant BEAN scores. Simplified interaction types and distance flags as annotated by Arpeggio are shown in the legend. **f)** BEAN z-scores of positions with conserved hydrophobic residues are shown along with the LDLR class B repeat PFAM HMM logo. **g-i)** Local atomic interaction in wild-type and mutated structure for ClinVar conflicting variants or VUS L426P, M652T, and Y532C. Residues in the variant positions are colored by the reference amino acids. Residues that interact with the variant position are shown. Variant position and interacting residues are colored by elements (O: red, N: blue, S: yellow). **j)** Contour plot of BEAN z-score against ΔΔ𝐺 predicted by DDMut for 872 missense variants. Positions with distinct observed missense variants that disrupt and conserve hydrophobic sidechains are connected by dashed line.

## Discussion

In this work, we develop a framework to improve variant effect quantification in base editing screens through accounting for variable genotypic outcomes estimated by gRNA efficiencies and chromatin accessibility. The activity-normalized approach is straightforward to apply, simply requiring synthesis of gRNA-reporter pairs, which can be cloned and screened using standard experimental procedures. This approach should prove particularly useful when employing new CRISPR enzymes and deaminases, as it allows for simultaneous characterization of editing preferences and phenotypic screening. We note that, while the screens described herein utilized flow cytometric phenotypic readouts, BEAN should also be suitable in dropout and enrichment paradigms by performing reporter analysis at an early timepoint prior to extensive phenotypic selection. We provide an open source implementation of BEAN in the comprehensive Python package *bean* with end-to-end functionality from base editing screen sequencing data to variant effect quantification at https://pypi.org/project/crispr-bean/.

Our results show that careful Bayesian modeling of the data generation process can substantially improve analytical power. We show that incorporation of reporter editing outcomes (BEAN-Reporter) and accessibility (BEAN) improves classification over the minimal model (BEAN-Uniform). In our work, we take into account the dependence of editing on loci accessibility from measurements taken from 49 gRNAs at four loci and use the relationship to improve our model (we provide a guide on fitting this relationship in **Supplementary Note 2**). Higher-throughput measurements and incorporation of additional epigenetic features influencing base editing could refine the inference of endogenous editing efficiency. It’s also worth noting that BEAN currently does not consider off-target editing, an omission that may affect the evaluation of phenotypic impacts for certain gRNAs.

BEAN also makes certain assumptions regarding how data is distributed. It assumes that phenotypic readout follows a Gaussian distribution and that alleles with multiple variants show additive effects. We also present an approach to modeling multivariate gRNA and allele count data by employing a Dirichlet-Multinomial distribution, building on the Negative Binomial modeling of counts used in prior methods^82,83^ (**Supplementary Note 3**).

We show that BEAN outperforms existing analysis methods at classifying GWAS variants with phenotypic impacts. Applying BEAN to the LDL-C GWAS library screen, we found it superior to existing methods at distinguishing positive control splice-altering variants. Furthermore, BEAN enabled accurate inference of variant effects on LDL-C uptake that were recapitulated using individual lentiviral gRNA transduction.

We used BEAN to uncover common variants that modulate LDL-C levels through altering liver cell expression/function of three previously unappreciated genes, *OPRL1*, *VTN*, and *ZNF329*. It is unclear why individuals with rare deleterious variants in these genes do not show altered LDL-C levels, but given that these genes have evidence of selective constraint^84^, we speculate that variants that are tolerated in the population may have weak functional and phenotypic effects^85^. The GWAS effect sizes for these variants are small, so they are unlikely to represent therapeutic targets; nonetheless, their elucidation contributes to understanding of the complex genetic underpinnings of lipid homeostasis.

We additionally demonstrate that dense activity-normalized base editing screens can improve characterization of coding variants in *LDLR*. Prior coding sequence-targeted base editing screens have used more restrictive PAMs^13,15–18,20–23,25–27^, thus limiting their breadth. By combining ABE8e-SpRY, which we show to have robust activity across the vast majority of PAMs, with BEAN, which enables sharing of information across adjacent gRNAs to boost power, we obtain accurate phenotypic measurements for an average of one variant per amino acid. This resolution is less than that of DMS, which can evaluate all possible missense variants at each position^4,86^. However, base editing is considerably less work-intensive and less expensive to perform and is far more scalable, allowing assessment of sets of genes in a single experiment. We provide pathogenicity assessment of 874 *LDLR* missense variants, most of which do not have prior clinical designation. Structural characterization of identified *LDLR* variants reveals distinct domain-specific characteristics of the most deleterious variants, including a central role of hydrophobic interactions gluing adjacent LDLR class B repeat domain’s beta blades.

Past coding variant editing screens have focused on binary classification of pathogenic and benign variants, and BEAN effectively distinguishes these variant classes in *LDLR*, while also making predictions about dozens of variants of unknown significance and conflicting annotation. Notably, we also show that BEAN scores associate with quantitative serum LDL-C levels measured in patients with rare *LDLR* variants in the UKB. Functional assays are accepted as evidence of pathogenicity or benignity in clinical variant interpretation guidelines such as those published by the American College of Medical Genetics and the Association for Molecular Pathology^87,88^. However, these frameworks have focused on classifying larger effect pathogenic and benign variants in a binary fashion, and these approaches have not been designed to offer risk predictions for quantitative traits. Given that CAD risk depends proportionally on the level of lifelong serum LDL-C exposure^89,90^, our ability to assign quantitative estimates of clinical risk to *LDLR* variants has strong clinical significance that, if replicable in other disease-associated genes, may merit adopting an assessment paradigm for clinical risk based on quantitative traits. We additionally show, albeit on a small cohort, that *LDLR* functional scores associate more closely with CAD incidence than LDL-C levels. This result is consistent with the residual CAD risk of FH patients even after LDL-reductive therapy^91^ and reinforces the value of *LDLR* pathogenicity analysis above and beyond LDL-C monitoring.

We show that base editing-derived functional scores correlate moderately with evolutionary conservation (Pfam, PhyloP) as well as structural disruption (DDMut) analyses. We show preliminary evidence that integrating BEAN and PhyloP scores improves prediction of LDL-C levels. Thus, functional screening offers at least partially orthogonal information to other forms of evidence that have been used to build computational pathogenicity predictors^92–94^. We anticipate that principled integration of distinct lines of evidence including MAVE data will improve pathogenicity prediction, since each evidence type is an imperfect surrogate for the effects of a variant over the lifespan of an individual. For example, our LDL-C uptake screening fails to measure how variants impact uptake of other lipoproteins by LDLR^95^, interaction with PCSK9^96^, or expression or function across environmental conditions or cell types, and thus its accuracy may be improved by integrating orthogonal methods. In conclusion, activity-normalized base editor reporter screening with BEAN markedly improves variant impact quantification from CRISPR base editing screens. Given the importance of variants of weak effect in complex disease, this approach promises to accelerate the characterization of human disease-associated variants in their native genomic context.

## Methods

### Establishing Cell Lines

HepG2 cells were obtained from American Type Culture Collection (ATCC). HepG2 cells were infected with lentiviral constitutive base editor vectors pLenti_ABE8e-SpRY-P2A-BFP_HygroR and pLenti_AID-BE5-SpRY-P2A-BFP_HygroR. After Hygromycin selection, cells were sorted twice to enrich for BFP expression.

### Screen library design

The LDL-C GWAS library was constructed to include gRNAs targeting variants associated with LDL-C levels from the UK Biobank GWAS cohort. Fine-mapped variants with posterior inclusion probability (PIP) > 0.25 from either the SUSIE or Polyfun fine-mapping pipelines (updated in December 2019, downloaded from https://www.finucanelab.org/data) were included, as well as variants with PIP > 0.1 within 250 kb of any of 490 genes found to significantly alter LDL-C uptake from recent CRISPR-Cas9 knockout screens^1,2^. All HepG2 haplotypes derived from the phased HepG2 genome sequence were targeted, and thus multiple allelic variants were targeted at certain genomic locations. Variants were assigned to ABE or CBE sub-libraries according to the identity of the affected nucleotide, and variants were included even in cases such as transversions and variable-length variants in which the edited variant would not exactly match the alternate allele identity. Five gRNAs that position the variant in protospacer positions 4-8 were included. gRNAs that contain TTTT stretches were removed from the library. Positive control gRNAs were designed to ablate all feasible splice donor and acceptor sites for ABE and to install 16 stop/gain variants for CBE, using five gRNAs per target site using the same logic as for variants. A total of 100 negative control non-targeting gRNAs were included in each sub-library, designed as 20 sets of five tiled gRNAs using the same logic as for variants. Paired reporter sequences were designed as the 32-nt genomic target sequence centered on the 20-nt gRNA.

The LDLR tiling library was constructed to include all gRNAs targeting coding regions (tiling density of 1) on both strands. The 26-nt intronic region surrounding each LDLR exon was tiled at ½ density (on both strands as in all subsequent cases), and the 24-nt distal to this region was tiled at 1/3 density. 5’ and 3’ UTR regions were tiled at 1/3 density. The two strongest LDLR intronic enhancers, both in intron 1, were also tiled at 1/3 density. In all cases, gRNAs were designed to match the HepG2 genomic sequence, and gRNAs were designed to target all HepG2 haplotypes when HepG2 is heterozygous at that sequence. gRNAs that contain TTTT stretches were removed from the library. 150 negative control non-targeting gRNAs were included. Paired reporter sequences were designed as the 32-nt genomic target sequence centered on the 20-nt gRNA.

Libraries were designed in the following template:

TGGAAAGGACGAAACACCG[19-20-nt gRNA]

GTTTAAGAGCTATGCTGGAAACAGCATAGCAAGTTTAAATAAGGCTAGTCCGTTATCAACTTGAAAAAGTGGCACCGAGTCGGTGCTTTTTTT[32-nt target (6-nt upstream, 20-nt gRNA, 6-nt PAM)][4-nt barcode]

AGATCGGAAGAGCACACGNNNNNNNNNNNNNN

Where the final 14 N’s are a variable primer sequence enabling pooling of sublibraries into a single synthesis order. The gRNA libraries were ordered from Agilent.

### Base editing screening

The gRNA libraries were cloned into either CRISPRv2FE-ABE8e-SpRY-BsrGI or CRISPRv2FE-AIDBE5-SpRY-BsrGI. Libraries were amplified using NEBNext Ultra II Q5 mastermix, cloned using NEBuilder HiFi DNA Assembly mastermix, and electroporated into Endura competent cells (Biosearch Technologies) for propagation. Lentivirus was produced in HEK293T cells for each library using TransIT-Lenti transfection reagent, titered, and incubated with 6.25*10^6 ABE8e-SpRY stable HepG2 and AID-BE5-SpRY stable HepG2 cells respectively per replicate at an MOI of 0.3-0.5. Five or more biological replicate screens were performed for each library. After 24 hours, media containing the lentivirus was removed and fresh DMEM media + 2mM VPA was added to allow for construct integration and promote base editing. After another 48 hours of media + VPA treatment, media with 1:20,000 Puromycin was added and cells underwent selection for the next 5-7 days and were split as needed.

Following complete selection as defined by complete death of a concurrent untreated control, cells were started on the 2-day LDL uptake protocol: on day 0, library cells were split, counted, and replated onto 15-cm plates at 1*10^5 cells/cm2. On the evening of day 1, DMEM media was removed and replaced with Optimem to induce overnight serum starvation. On the morning of day 2, 1:400 BODIPY FL-LDL (Thermo Fisher Scientific) + Optimem was added and incubated with cells for 4-6 hours. After this incubation period, plates were trypsinized, stained with 50 ng/mL DAPI, and sorted based on BODIPY-LDL fluorescence levels into 4 bins (top 20%, top 20-40%, bottom 20%, and bottom 20-40%). gDNA was then collected from each sorted population as well as an unsorted bulk population and prepared for next generation sequencing.

### Library preparation and next generation sequencing

To prepare for next generation sequencing of samples, genomic DNA collected from each sorted population and an unsorted bulk population was used as the input for a PCR1 reaction aimed at amplifying the integrated construct spanning the gRNA, as well as adding different inline PE1 barcodes to specific samples for downstream analysis. The ideal total genomic DNA input per sample for PCR1 was 20ug of DNA, but if less than 20ug of genomic DNA was collected for a specific sorting population, then the total genomic DNA yield was input into PCR1. NEBNext Ultra II Q5 mastermix was used. Please refer to **Supplementary Table 9** for specific PCR1 primers used depending on the specific reporter and corresponding sub-experiment described above in the paper. Following PCR1, reactions were individually PCR purified using a standard QIAquick PCR Purification Kit. Next, a qPCR2 was performed from 0.25ul of each purified sample in a 15ul qPCR reaction to determine the optimum number of cycles for PCR2, and the products of qPCR2 were run on a gel to confirm lack of primer dimer and the confidence of CT cycles from qPCR2. PCR2 cycle counts were chosen to be 2-3 cycles less than the qPCR2 CT for the corresponding sample, with a minimum number of PCR2 cycles being 7. In order to set up PCR2, half of each sample’s purified PCR1 product was then used with NEBNext Ultra II Q5 mastermix (please refer to the **Supplementary Table 9** for specific PCR2 primers used. It is important to note that, despite differences in primers between sub-experiments, NEBNext i7 primers were always used in PCR1, and NEBNext i5 primers were always used in PCR2. The unique combination of these two barcodes for each specific sample is what allows for downstream identification of reads post sequencing. Following PCR2, samples were again PCR purified with a QIAquick PCR Purification Kit, and then purified samples were run on a 2200 Agilent Tapestation to identify the quantity of product as well as any unwanted byproducts and primer dimers that might have occurred throughout NGS preparation. Samples were then pooled based on their molarity, and the pool was purified to remove unwanted products using SPRI-Select beads from Beckman Coulter. The SPRI bead: sample ratio for pool purification was chosen based on the quantity and size of unwanted byproducts relative to the desired product, and varied among 0.8-0.9 for all experiments. Following bead purification, the pooled sample was sequenced using Illumina Nextseq used paired-end sequencing with >50-nt read 1 and >36-nt read 2.

### Comparison of endogenous and reporter editing

Endogenous sublibraries A-D targeting four distinct loci across *LDLR* (Supplementary Table 3) were cloned and run as the screens (see Screen Procedure above), but were run in six-well format with 400,000cells per sample six well being infected. Following the standard lentiviral infection protocol and puromycin selection process, samples were harvested at the end of selection as only editing data was desired. After gDNA isolation, a reporter-based library preparation for next generation sequencing was performed (see Library preparation for next generation sequencing protocol above), together with an additional library preparation for amplifying endogenous editing sites: PCR1 samples to determine endogenous editing were set up with Ultra II Q5 mastermix and 2.5ug of genomic DNA input and a unique primer mix for each of the four LDLR endogenous libraries. These primer mixes contained three unique R1 forward primers and one common R2 reverse primer that would allow amplification segments from the endogenous LDLR containing all desired editing sites targeted within that library. Upon completion of PCR1, samples were PCR purified using a QIAquick PCR Purification Kit and prepared following the standard library preparation protocol above. For specific primers used, please see Supplementary Table 9). Following the preparation of these endogenous samples, both endogenous and reporter samples were run on an 2200 Agilent Tapestation and pooled + purified accordingly to prepare for next generation sequencing (see Library preparation and next generation sequencing section).

Endogenous target site reads are mapped to the reference amplicon sequences using CRISPResso2 with base editing mode and custom mismatch score matrix that tolerates A to G mutation generated by ‘*bean-count*’ command of *bean* software that implements all the computational functions of the proposed BEAN workflow as a Python package. Bean is available at https://pypi.org/project/crispr-bean/. The paired gRNA and reporter library is mapped to the expected gRNA and reporter sequences using ‘*bean-count*’ function of the *bean* package. Position-wise reporter base edits are tested for significance against control data without editing using the function ‘*bean.annotate.filter_alleles.filter_alleles’* from the *bean* package. This function conducts Fisher’s exact test and was used to filter for edits with Bonferroni-corrected P-value < 0.05 and odds ratio > 5.

### Read mapping

FASTQ files are first demultiplexed by matching the corresponding 8nt pair-end index sequences (I2 sequences “NNNNNNAG” are partially degenerative). Then, the FASTQ files are further demultiplexed by exact matching of the 3-6nt barcodes and the U6 hairpin stub (“GGAAAGGACGAAACACCG”). Paired end reads in the demultiplexed FASTQ files are mapped to gRNA sequences and read for the reporter editing outcome using the ‘*bean-map-samples’* command from *bean*. There, read pairs where either R1 or R2 with average quality Phred score lower than score 30 are discarded. Read pairs with good quality are mapped to the spacer sequence where the spacer positions in R1 perfectly match with any of the provided gRNA sequences while being masked for the editable positions to account for self-editing. As 26%-54% of the reads are shown to have undergone R1-R2 recombination (Supplementary Fig. 25), we assign the read to two categories, where both R1 spacer sequence and R2 barcode sequence uniquely matches to a gRNA in the library and where on the R1 spacer sequence has the unique match to the gRNA sequence. For the reads that have the correct R1-R2 matching (barcode matched reads), we count the allele-level editing outcome by comparing the reference reporter sequence to the R2 reporter sequence by globally aligning two sequences using the modified ‘*CRISPResso2Align.global_align*’ function of CRISPResso2^3^ for base editing. The matched gRNA count, all gRNA count regardless of R1-R2 matching, and per-guide reporter allele counts for the matched gRNAs are stored as the output per each sequencing sample.

### Quality control of reporter screen data

To exclude failing samples and outlier guides, ‘*bean-qc*’ from *bean* is run on the mapped gRNAs and edited allele counts. Specifically, samples with median Spearman correlation of gRNA count smaller than 0.8, median log fold change of positive control guides (gRNAs targeting splicing sites for both LDL-C GWAS and LDLR tiling library) in top 20% and bottom 20% quantile smaller than -0.1, or median gRNA editing rate in reporter smaller than 0.1 are labeled as low-quality. The gRNA editing rate is calculated as the target variant cognate (A to G in ABE) editing rate in LDL-C GWAS library and mean cognate (A to G in ABE) editing rate in editable base (A for ABE) protospacer position 3-8. Outlier gRNAs and replicate pairs are defined as the gRNAs with a median absolute deviation larger than 5 and RPM (reads per million) ≥ 10000 among replicates.

### Profiling and visualization of base editing preference

LDLR tiling library editing outcome in bulk samples were analyzed to profile base editing preferences, and avoid bias in sequence context that comes from having A in designated position in LDL variant library. The heatmap of editing preference across spacer position and PAM was generated by the function ‘*bean.pl.editing_patterns.plot_by_pos_pam*’ from *bean.* This considered the mean editing efficiency of A to G transitions within protospacer positions 1-20 for 7320 targeting gRNAs with more than 10 reads in any bulk sample. The average editing range per dinucleotide in the PAM was calculated as the maximal editing rate in protospacer positions 3-8, where the the relative editing rate is the highest. To quantify the context preference, the mean A to G editing efficiency of the –1 and +1 bases of the intended target base in protospacer position 3-8 was calculated. The context preference logo was generated from the normalized mean editing efficiency across replicates by the nucleotide in –1 and +1 positions with the Logomaker package.

### Prediction of editing outcomes with BE-Hive

We used the Python implementation of BE-Hive^4^ (https://github.com/maxwshen/be_predict_bystander) to predict editing outcomes of the *LDLR* tiling library. We initialized the model with “mES” as cell type and “ABE8” as the editor due to the lack of HepG2 cell type model. For each spacer, we extracted a 50nt long sequence around the starting position of the spacer in the hg38 genome, with 20nt before the spacer start and 30nt after,on the same strand. These 50nt sequences were used as the input to the BE-Hive to predict likely editing outcomes. To calculate allele-level edit rates for each spacer, we summed up the probability of any editing outcomes with the same editing patterns in position 0-18 (0-based) relative to the start of the spacer. Similarly, to calculate base-level edit rate, we summed up the probability of any editing outcomes with identical base edits in position 3-8, relative to the start of the spacer.

## BEAN models

### Variant and gRNA-level phenotypic modeling with reporter

BEAN models the phenotype of cells with variant 𝑣, 𝑌_*v*_, as Normal distribution, where the wild-type cells have standard normal phenotypic distribution 𝑌_0_ and the variant effects are quantified in a relative scale, using 𝑌_0_ as reference. For the cells with a gRNA, their phenotype is modeled as a mixture of allelic distributions produced by the gRNA, reflecting the heterogeneous outcome from a gRNA.

For variant screens (LDL-C GWAS library), we aggregate alleles into two categories: alleles with or without observed edits at the target variant. The non-edited component in these models is fixed to have a wild-type phenotypic distribution. That is, the phenotype 𝑌_*g*_ of cells with gRNA 𝑔 that induces variant 𝑣 with editing rate 𝜋 is modeled as follows:

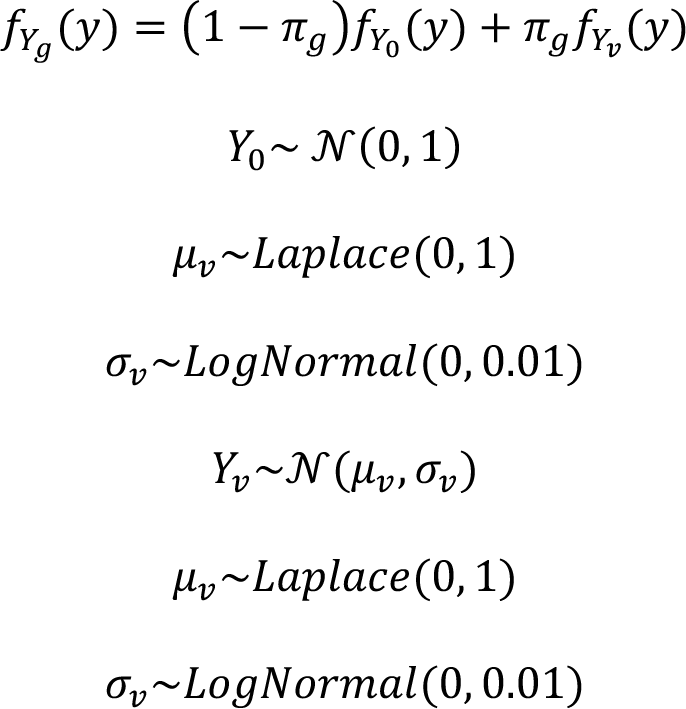

where 𝑓_*Y*_ indicates the probability density function of 𝑌. The prior for 𝜇_*v*_ and 𝜎_*v*_ are set to be narrow based on the assumption that most variant would have close to wild-type effect size of mean 0 and standard deviation 1.

For saturation tiling screen, as bystander edits are more likely to have phenotypic effect, BEAN accounts for more than one non-wild-type allele where each allele may include one or more variants. Here, we use the term “allele” to refer to the multiple editing outcome produced by base editing, and we aggregate multiple nucleotide-level variants that lead to the same coding sequence amino acid mutations together. To account for splicing and noncoding region variants, variants that fall outside coding regions are not aggregated.

We denote with 𝐴(𝑔) = {𝑎|𝐴𝑙𝑙𝑒𝑙𝑒 𝑎 𝑖𝑠 𝑝𝑟𝑜𝑑𝑢𝑐𝑒𝑑 𝑏𝑦 𝑔} the set of alleles produced by gRNA 𝑔 that is robustly observed, here in at least 10% of the gRNA read counts across 30% of the samples after above-mentioned aggregation. However, we note that users have the flexibility to set their own robustness thresholds in ‘*bean-filter*’ of *bean* package. The phenotype of a given allele 𝑎 is defined as the sum of phenotypic effect of non-wild-type nucleotide and amino acid level variants. Finally, the phenotype of cells with gRNA 𝑔 is modeled again as the mixture distribution of allelic phenotype for the alleles it induces (𝑎 ∈ 𝐴(𝑔)) as follows:

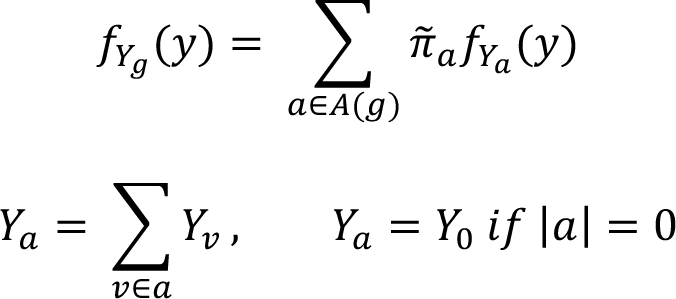

where 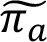 is the endogenous editing rate, estimated from 𝜋_*α*_, the reporter editing rate, of allele 𝑎. The non-edited allele phenotype and the priors for 𝜇_*v*_ and 𝜎_*v*_ are identical to the variant screen modeling. The identity of the alleles and their frequency in reporters are learned from per-gRNA reporter allele counts in pre-sort (bulk) sample. Within this modeling framework, the allelic editing frequency in the reporters is proportionately adjusted based on chromatin accessibility of the intended gRNA target locus to better estimate the endogenous allele frequency, while allowing for deviation from the scaled values. For each gRNA, allele editing rate 𝝅_***g***_ = (𝜋_*g*0_, …, 𝜋_*g*_|*A*(*g*|) and per-gRNA allele count 𝒁_𝒈_ = (*A*_*g*0_, …, *A*_*g|A(g)|*_) are modeled as the Dirichlet and Multinomial distributions :

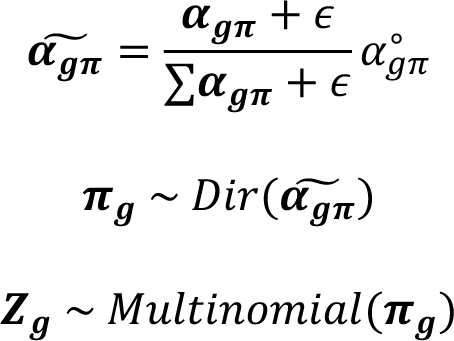

Where 𝜶_𝝅_ is initialized as 1⃗, 𝜖 = 1*e*^−5^ and 𝛼^∘^_*gπ*_ is the precision parameter that is fitted from the data (**Supplementary Note 3**). This approach partially follows DESeq2^5,6^ approach of dispersion parameter estimation for the Negative Binomial distribution. The reporter editing rate 𝝅_𝒈_ is further scaled by accessibility to be used as the endogenous editing rate 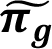 through a function 𝑓. This function 𝑓 is learned a priori from the paired reporter and endogenous editing rate data while the deviation of 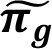 from *f*(π_***g***_) is fitted per gRNA. The deviation 𝜖_π*g*_ below accounts for the incomplete correlation between endogenous and reporter editing rates.

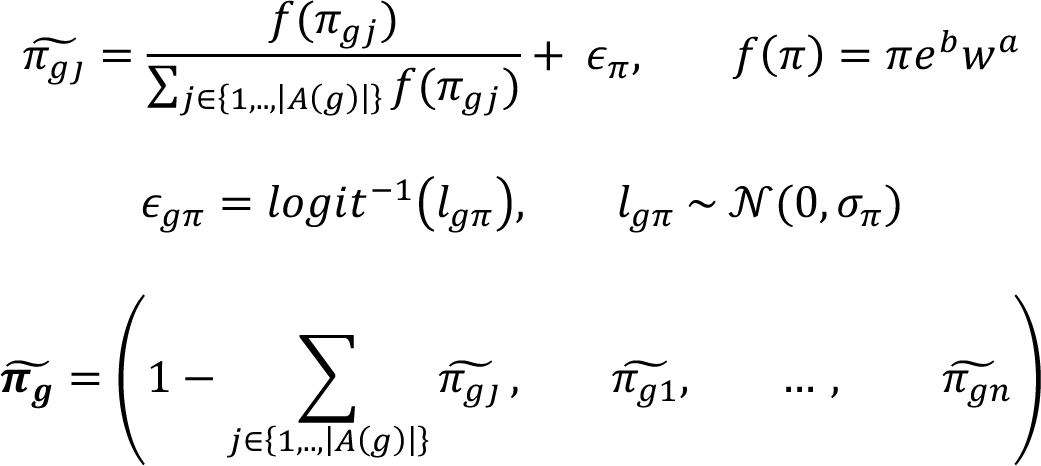

𝑓(𝜋) is fitted from the data generated for comparison of endogenous and reporter editing based on the regression 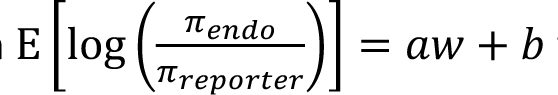 where 𝑤 is 𝑙𝑜𝑔(𝑎𝑐𝑐𝑒𝑠𝑠𝑖𝑏𝑖𝑙𝑖𝑡𝑦 𝑠𝑖𝑔𝑛𝑎𝑙 + 1) and the resulting coefficients α = 0.2513 and *b* = −1.9458 are used for the analyses presented in this paper. The residual of the regression is fitted as the Normal distribution, which is used as the prior for the logit-scale deviation 𝑙_π_ (see full detail in **Supplementary Note 2**).

In modeling base editing data screens with reporter data, we have built and evaluated two variants of the BEAN model that utilize less information than the original model. The first variant, BEAN-Uniform assumes a single component Normal distribution of cellular phenotype, reflecting the assumption that all gRNAs would have the same editing efficiency.

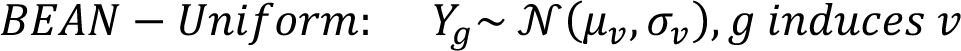

While BEAN utilizes both reporter data and accessibility to estimate endogenous editing efficiency from the reporter data, the second variant, BEAN-Reporter focuses only on the incorporation of the reporter data without accessibility information. That is, for BEAN-Reporter, **π = π**.

### Sorting screen and gRNA count data modeling

Sorting screens sorts the pool of cells with different gRNA and editing outcomes into distinct bins based on the phenotype they’re sorted on prior to sequencing. To model the sorting procedure, the proportion of cells that falls within sorting quantile bins for each gRNA is calculated analytically. This process allows for the determination of the relative fraction of cells with the gRNA that falls into each sorting bin, which is then used as the concentration parameter of Dirichlet-Multinomial distribution. Dirichlet-Multinomial distribution is chosen to model the gRNA read count across sorting bins that is over-dispersed multinomial count distribution, which we confirm from our data (see **Supplementary Note 3**). The gRNA read counts across sorting bins 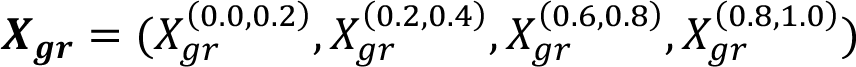 and the barcode-matched gRNA read count ***X^b^_gr_*** for gRNA 𝑔 and replicate 𝑟 are modeled as following:

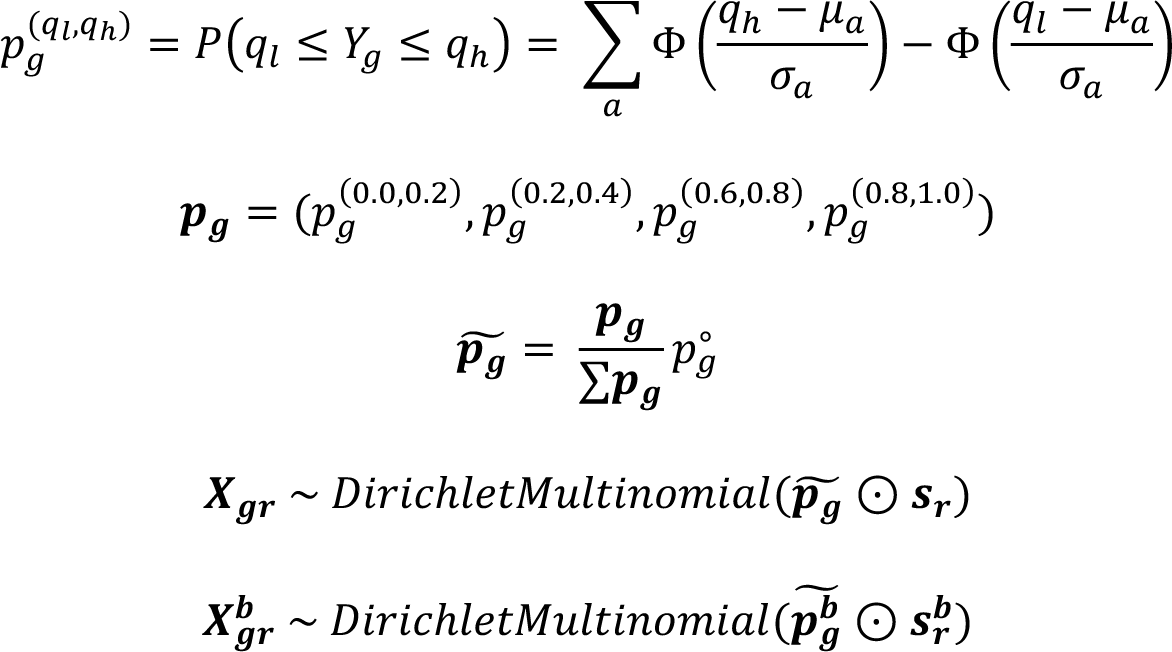

where ⊙ denotes element-wise multiplication. Here, *𝒑_g_* is scaled as 𝜶_𝝅_ by the data-fitted precision parameter 𝑝^∘^_*g*_ (**Supplementary Note 3**) then scaled by the sample-specific size factor ***s_r_*** = 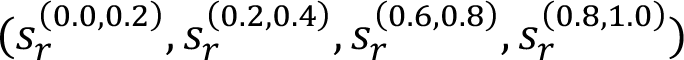, where the sample size factor is calculated as in DESeq2^5,6^. For sample 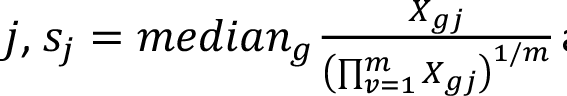 and the same function is used to calculate size factor for barcode-matched read counts for sample 𝑗, 𝑠*^b^_j_* with *𝑋^b^_gj_*. We note that 𝑿^𝒃^ is not used in the inference when 4 %4 𝒈𝒓 benchmarking against other methods to make sure we provide the same input. Samples marked as low-quality and gRNA and replicate pair with ≤10 total reads, or identified as outliers are excluded from inference (see **Quality control of reporter screen data**).

The parameters 𝜇_*g*_, 𝜎_*g*_, 𝛼_π_, 𝑙_π_ of posterior distributions are fitted using stochastic variational inference (SVI) of Pyro using 2000 steps with decaying learning rate starting from 0.01 with variational distribution that mirrors the model. Specifically, the posterior phenotypic distribution of each variant is fitted as a Normal distribution with a posterior standard deviation parameter and mean parameter which has Normal posterior distribution:

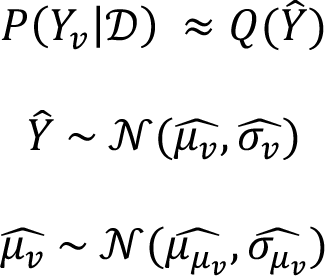

Where 𝒟 is observed data for the model and 𝑄 is the variational distribution. Negative control variants are used to control the significance of variant effect, by fitting the shared phenotypic distribution of negative controls as a single normal distribution. Subsequently the results are scaled so that the fitted negative control distribution is transformed to a standard normal.

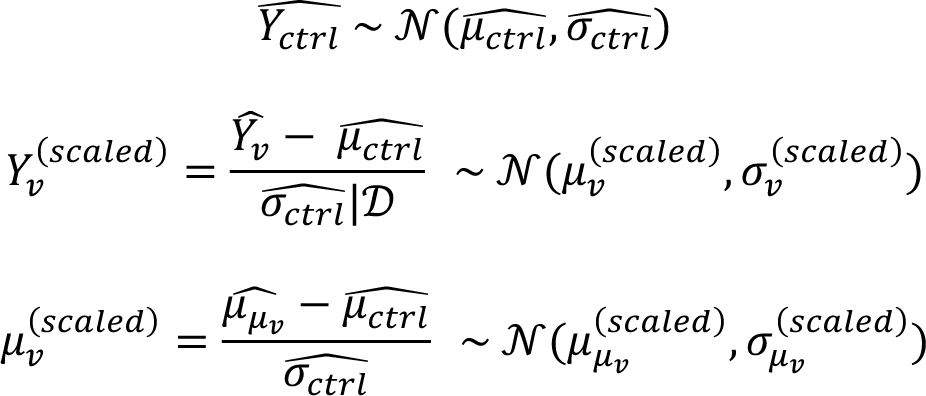

^𝑌à^In order to control for false discovery with negative control variants, the standard deviations of variants 𝜎(LJ’DMN) are scaled so that the standard deviation of 𝜇_;_, where 𝑛 are the negative control variants, is equal to 1. In the LDL-C variant screen, 20 negative control variants (each tiled by 5 gRNAs) are used and for LDLR tiling screen 175 synonymous variants are used as the negative control variants:

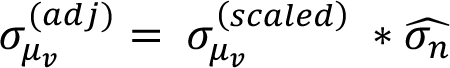

Where 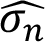 is fitted as the standard deviation estimate of 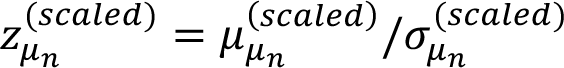 with ‘stats.norm.fit’ of Python’s SciPy package with setting location parameter to 0. The model’s output includes various parameters relating to the phenotype of the variant, such as the mean and standard deviation of variant phenotype 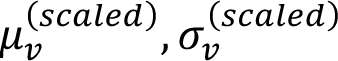 and scaled and significance-adjusted phenotypic mean distribution parameters 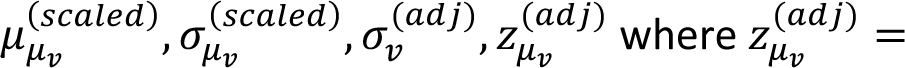 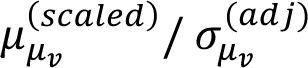 are reported together with estimated endogenous editing efficiency for each variant. For ‘variant’ mode, the mean targeting gRNA editing rate is reported and for ‘tiling’ mode, effective editing efficiency is reported and calculated as 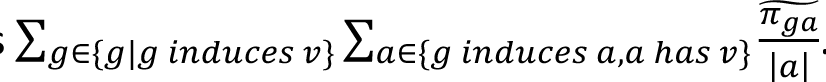. The model, variational distribution and inference procedure are available as the default options of ‘*bean-run*’ command of *bean* software. Specifically, BEAN-Uniform is run with ‘*--uniform-edit*’ and full BEAN model is run by specifying ‘*--scale-by-acc*’ argument.

### Benchmarking of CRISPR Pooled Screen Analysis Methods

We reviewed and selected several CRISPR pooled screen analysis methods for benchmarking against BEAN, based on their availability and applicability to our experimental design and sorting screens. BAGEL^7^ was not applicable as it required positive and negative control target genes as the input. ACE^8^ was designed for gene essentiality screens. Gscreend^9^ required a single unsorted population to be compared against the multiple treatment samples. Consequently, we chose MAGeCK-RRA^10^, three running modes of MAGeCK-MLE^11^, CRISPRBetaBinomial (CB2)^12^, and CRISPhieRmix^13^. CRISPhieRmix is only used for benchmarking the *LDLR* tiling screen as it requires the negative control gRNAs as LDL-C GWAS library benchmarking uses negative control variant label to evaluate classification performance. Huang et al., (2021)^14^ didn’t offer their method available as software, but we incorporated their efficiency correction concept as efficiency-corrected log fold change (EC-LFC). Log fold change of variants calculated from MAGeCK-RRA was included as the baseline. We believe that these methods represent the state-of-the-art based on multiple recent benchmark studies.

We used two modes of MAGeCK v0.5.9.4, MAGeCK-RRA^10^ and MAGeCK-MLE^11^. MAGeCK-RRA takes treatment and control samples and evaluates if the rank of log fold change of gRNA abundance is not uniformly distributed. We used the paired mode with bottom 20% and top 20% quantile bin samples of each replicate as the paired treatment and control.

MAGeCK-MLE^11^ uses Negative Binomial generalized linear model with log link to output the coefficients, that can be interpreted as the log fold change of the gRNA abundance following the unit increase in the covariate that is provided in the input design matrix. MAGeCK-MLE is the only method that we benchmakred against that can use all 4 quantile bins of our sorting screens. We assigned 0, 1, 3, 4 to 0-20%, 20-40%, 60-80%, 80-100% quantile bin samples as the input covariate values. We further benchmarked MAGeCK-MLE where it uses the gRNA activity (‘*---guide_efficiency_file*’) or fits the gRNA activity (‘*--guide_efficiency_file --update-efficiency*’). As MAGeCK-MLE assumes gRNA efficiency in guide_efficiency_file to scale from -1 to 0.25, the editing efficiency is normalized to the range. In case the gRNA has not enough reads and is not assigned of the editing rate, it is assigned to the editing rate of 0.5 before the scaling. All runs are ran both with (‘*--genes-varmodeling* 1000’) or without (default) dispersion fitting.

CB2^12^ models the gRNA counts using the beta-binomial distribution in which the variance can be either large or smaller than the mean to quantify gRNA abundance for CRISPR pooled screen data analysis. It uses Fisher’s combined probability test to estimate the gene-level significance. We installed the CB2 package (v1.3.4) and benchmarked the performance by comparing the bottom 20% and top 20% quantile bins.

CRISPhieRmix^13^ is a hierarchical mixture model for analyzing CRISPR pooled screen data by assuming that the majority of genes does not impact phenotype. It builds a two-group mixture model to identify the impactful target genes. Specifically, log 2 fold change between bottom and top 20% quantile bins is calculated from DESeq2^6^ and used as the input to compare the bottom and top samples.

EC-LFC was calculated by dividing the variant log fold change calculated by MAGeCK-RRA^10^ by the variant editing efficiency. For the LDL-C GWAS library, mean editing efficiency of the targeting gRNAs are used as the variant editing efficiency. For *LDLR* tiling library, effective editing efficiency was used as the variant editing efficiency.

### Benchmark on LDL-C GWAS and *LDLR* tiling library

For both LDL-C GWAS library and *LDLR* tiling library, the classification performance AUPRC of distinguishing positive controls against negative controls are evaluated. For the benchmark, 6 biological replicates of LDL-C GWAS library and 4 LDLR tiling library with no failing samples are used, and barcode-matched reads are ignored during inference in BEAN runs. For replicate subsample analysis, all possible 2-replicate combinations are subset to be analyzed by each method. For LDL-C GWAS library, its positive control variants, which are the splice sites of the genes that changes the LDL-C uptake is used as the positive control variants and the 20 non-targeting negative control variants are used as the negative control variants. For LDLR tiling library, ClinVar “pathogenic” or “pathogenic/likely pathogenic” annotated variants are classified against ClinVar “benign” or “benign/likely benign” annotated variants. As each method has different strategy to assign gRNA to variant thus scores different set of variants, we evaluate the recall as how much of the all ABE-discoverable Pathogenic/Likely Pathogenic variants are identified as Pathogenic.

### Cloning and testing of individual gRNAs

#### Base Edit

Oligonucleotides including protospacer sequences were ordered in the following format: GGAAAGGACGAAACACCG [19-20-bp protospacer —remove initial G for any 20-bp protospacer with one natively] GTTTAAGAGCTATGCTGGAAAC (see Supplementary Table 9). Using NEBuilder HiFi DNA assembly, ABE8e-Cas9NG designated oligonucleotides were cloned into CRISPRv2FE-ABE8e-Cas9NG, while ABE8e-SPRY designated oligonucleotides were cloned into CRISPRv2FE-ABE8e-SpRY-BsrGI. To make base edited cell lines, the gRNA constructs were packaged into lentivirus and transduced into HepG2 ABE8e-SPRY-BFP cells seeded at 4×10^4^ cells/cm^2^ on 6-well plates in two replicates with 8 µg/ml polybrene. Two days post-transduction, cells were treated with 500 ng/ul puromycin and selected for approximately one week. HepG2 Base Edited cells were seeded 1:1 with HepG2-mcherry cells to achieve a total density of 1.08×10^5^ cells/cm^2^ on a 96 well plate in at least two technical replicates of two biological replicates and incubated overnight. The next day, the media was replaced with optiMEM and cells were incubated overnight. Approximately 4-6 hours prior to flow cytometric analysis, cells were treated with 2.5 mg/mL BODIPY-LDL in optiMEM. Cells were trypsinized and analyzed for presence of mCherry and LDL uptake using a Beckman CytoFLEX flow cytometer. LDL uptake of each base edited cell line was normalized to the LDL uptake of the mCherry cells within the same well. Differential LDL uptake between base edited and control cells was further normalized using data from the ABE8e and SPRY sgCTRL lines.

#### CRISPRi

Oligonucleotides including protospacer sequences (**Supplementary Table 9**) were ordered in the following format: GGAAAGGACGAAACACCG [19-20-bp protospacer—remove initial G for any 20-bp protospacer with one natively] GTTTAAGAGCTATGCTGGAAAC were cloned into a pHR-U6-gRNAFE-Zim3-dCas9-P2A-Hygro backbone through NEBuilder HiFi DNA assembly. To make CRISPRi cell lines, the gRNA constructs were packaged into lentivirus and transduced into HepG2 cells seeded at 4×10^4^ cells/cm^2^ on 48-well plates in two replicates with 8 µg/mL of polybrene. Two days post-transduction, cells were treated with 125 µg/ml Hygromycin B and were selected for approximately one week. LDL uptake experiments were performed as described above, seeding CRISPRi cell lines 1:1 with HepG2-tTA-BFP cells as the internal control.

#### CRISPRa

Oligonucleotides including protospacer sequences (**Supplementary Table 9**) were ordered in the following format: GGAAAGGACGAAACACCG [19-20-bp protospacer —remove initial G for any 20-bp protospacer with one natively] GTTTAAGAGCTAGGCCAACATG. Using NEBuilder HiFi DNA assembly, oligonucleotides were cloned into a pLenti U6-2xMS2gRNA MCPp65 PuroR backbone. To make CRISPRa cell lines, the gRNA constructs were packaged into lentivirus and transduced into HepG2 dCas9-10xGcn4-mChe + scFv-Sbno1-Nfe2l1-Krt40-BFP cells and seeded at 4×10^4^ cells/cm^2^ on 6-well plates in two replicates with 8 µg/ml polybrene. Two days post-transduction, cells were treated with 500 ng/ml puromycin and selected for approximately one week. LDL uptake experiments were performed as described above, seeding CRISPRa cell lines 1:1 with HepG2 wt cells as the internal control.

#### Pooled ATAC-seq

A pool of 20 gRNAs was cloned into CRISPRv2FE-ABE8e-Cas9NG or CRISPRv2FE-ABE8e-SpRY-BsrGI (see Cloning and testing of individual gRNAs), packaged into lentivirus and transduced into HepG2 ABE8e-SpRY-BFP cells seeded at 4×10^4^ cells/cm^2^ on 6-well plates in three replicates with 8 µg/mL of polybrene. Cells were treated with VPA and selected with Puromycin as in screens. Once selected, two sets of 1*10^6^ cells for each of the three replicates as well as an unedited control replicate were seeded to 6-well plates. The next day, one well per replicate was fed with DMEM + FBS and the other with optiMEM (serum-starved). 24 hours later, the wells were trypsinized, and 1*10^5^ cells were used for ATAC-seq while the remaining cells were used for bulk genomic DNA isolation using the Purelink Genomic DNA mini kit (Life Technologies). ATAC-seq was performed using the Active Motif ATAC-Seq kit according to manufacturer’s instructions.

To obtain valid primers to amplify the loci surrounding the 20 target variants, Primer3^15^ was used to generate 5 candidate primer sets within +-150-nt from each variant. Primer-Dimer.com was used to calculate a deltaG (dG) interaction matrix for all candidate primers. Primers with average dG <= -7 were removed. Then, recursive pairwise filtering was performed to iteratively remove the primer with the worst dG interaction until no pairwise dG <= -7 remains. This recursive filtering was performed 300 times, and the run with the most primers remaining was used. The primer set for each variant with highest minimum dG was selected. Primers were all ordered from IDT preceded by NNN to randomize initial nucleotides in NGS. We provide the amplicon sequence of 20 loci in Supplementary Table 9.

Genomic DNA and ATAC-Seq products from 8 total samples (3 experimental and 1 control, both in serum and starved conditions) were amplified using two primer pools, each composed of 10 primer sets to synchronize annealing temperature. 2.5 ug of gDNA/half of ATAC-Seq product was used in 100 uL reactions for 32 cycles (gDNA) or 35 cycles (ATAC-Seq). Tapestation was used to pool the two PCR products for each sample, and these 16 pools were used as input to the NEBNext UltraII DNA Library Prep to prepare NGS libraries. Libraries were sequenced using 150-nt single-end sequencing using Illumina Nextseq.

#### Pooled ATAC-seq analysis

For each sample, the ATAC-seq reads were mapped to amplicon sequences (**Supplementary Table 9**) from 20 loci via Bowtie2^16^ (v2.5.1). ‘bowtie2-build’ was used to build indices for the amplicon sequences for each of 20 loci and reads are mapped onto the indices with default parameters. In-house Perl script was used to parse the SAM output from Bowtie2 and to demultiplex the reads by the locus they mapped to with default options. Demultiplexed reads are then profiled for the target base editing rate using CRISPResso2^3^ (v.2.2.9) using average read quality cutoff of Phred score 30 and assigned of base ‘N’ if per-base quality is lower than Phred score 20. For each variant, reads are assigned to the reference allele or alternate allele based on the base identity at the target SNP position. In case there exists a neighboring variant that allows phasing as HepG2 is heterozygous for the variant, the reads are counted per phase based on the identity of the neighboring variant. We note that for two of the variants examined (rs3767844 and rs4390169), whether the base is the result of editing or is the reference allele was ambiguous (that is, variants are heterozygous in HepG2 and two reference alleles of A and G, then we cannot assign reads with G in the variant position to the edited reference A or unedited reference G). For the variants, we simply compare two observed bases and treat the effect as caQTL. For rs771555783, rs76895963, and rs116734477, edited reads are not detected due to insufficient representation of the loci, and thus excluded from the enrichment analysis.

We first identified the variants with significant editing observed in treatment samples compared to the control samples where base editors are not treated. This is done by assessing the significance of coefficient for 𝑖𝑠_𝑡𝑟𝑒𝑎𝑡𝑚𝑒𝑛𝑡 in the following Binomial regression with ‘*GLM*’ module of Python *statsmodels* package^17^, where *Edited_j_ Unedited_j_* is the read counts of edited and unedited variants in sample 𝑗, and 𝑖𝑠_𝑡𝑟𝑒𝑎𝑡𝑚𝑒𝑛𝑡_*j*_ is the indicator variable for the sample 𝑗 being treatment sample.

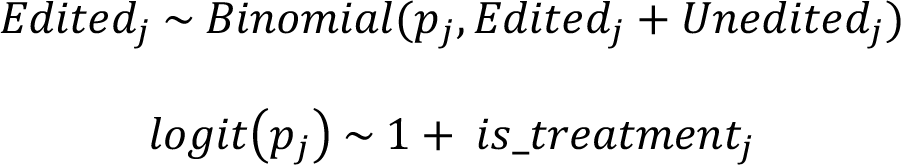

Significantly edited variants should show higher proportion of edited reads (𝑝_*j*_) in treatment samples compared to the control samples. For all significance testing, Benjamini-Hochberg family-wise error rate (FWER) value of 0.1 is used as the threshold, where multiple testing correction is performed with ‘*stats.multitest.multipletests*’ function of Python *statsmodels* package^17^.

For the variants with significant observed editing, we calculated the enrichment of the editing in ATAC-seq compared to the gDNA sample, which indicates the editing opened the chromatin at the variant loci and increased its capture rate for ATAC-seq. The enrichment of edited allele is calculated as the Binomial regression coefficient of edited and unedited read counts for each variant. The proportion of edited read count is regressed on whether the sequencing sample 𝑗 is from ATAC-seq (𝑖𝑠_𝐴𝑇𝐴𝐶_*j*_ = 1) or gDNA (𝑖𝑠_𝐴𝑇𝐴𝐶_*j*_ = 0), and the regression coefficient of 𝑖𝑠_𝐴𝑇𝐴𝐶_*j*_ is used as the accessibility enrichment of the variant editing. We condition for replicate and condition specific effect, along with the interaction effect between condition and ATAC-seq sample to examine if the variant only alters accessibility under either one of two conditions.

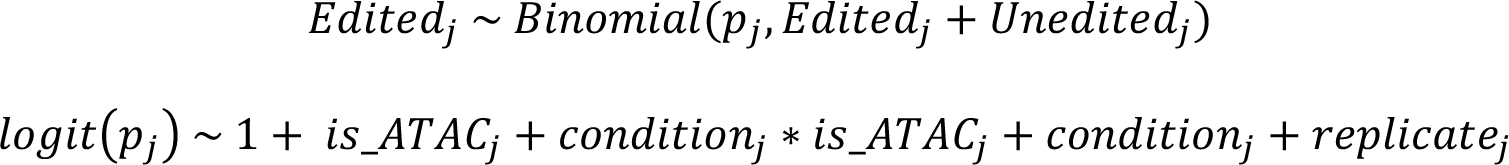

We also calculated the caQTL effect of the variants that are heterozygous in HepG2. Here, whether one allele has higher enrichment in ATAC-seq sample is examined as the regression coefficient for 𝑖𝑠_𝐴𝑇𝐴𝐶_*j*_ of following regression, again conditioned on experimental condition and replicate.

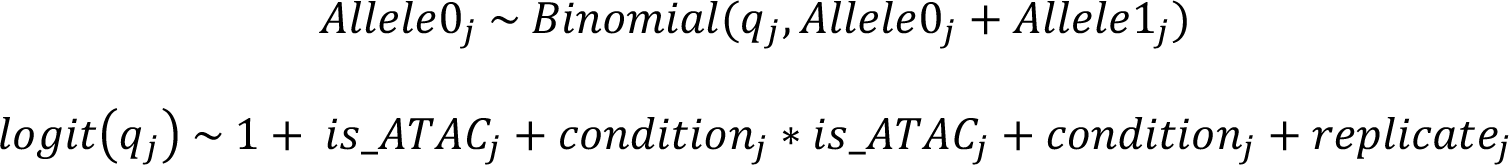

Here, 𝐴𝑙𝑙𝑒𝑙𝑒0_*j*_ and 𝐴𝑙𝑙𝑒𝑙𝑒1_*j*_ are the read counts of alleles 0 and 1. The regression coefficient for 𝑖𝑠_𝐴𝑇𝐴𝐶_*j*_ is used as the accessibility enrichment of allele 0. When the enrichment is shown uniformly in major to minor allele, enrichment values and confidence intervals calculated for the opposite direction is inverted of their sign.

### MotifRaptor

We adapted the MotifRaptor^18^ pipeline to investigate how prioritized genetic variants may influence nearby genes involved in LDL-C uptake. For each variant, we retrieved genomic sequences spanning 61 bp centered around the SNP location, using the hg38 genome assembly as a reference. Each sequence was mutated by substituting the major allele with the minor allele at the SNP position, yielding both a reference and an alternative sequence for each variant.

Subsequently, to evaluate the potential for transcription factor (TF) binding, we employed all the human TF position weight matrices (PWMs) from the CIS-BP database^19^ to scan each pair of reference and alternative sequences. This motif scanning generated binding scores at each sequence position, serving as predictive indicators of TF binding potential.

We then compared these scores for each TF across the reference and alternative alleles within every sequence pair. This comparative step is crucial for determining a variant’s impact on TF binding.

Specifically, higher binding scores for the alternative sequence indicate an increase in TF binding potential, while lower scores suggest a decrease. To quantify these changes, we calculated a ’disruption score’ as follows:

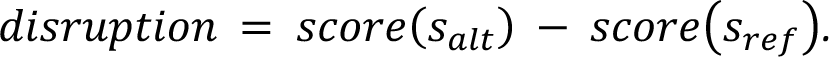

This score helps capture the directional change each variant induces, where a negative value signifies reduced TF binding potential and a positive value indicates an increase.

### Pfam profile HMM scores

Pfam profile HMM files of PF00057, PF00058 and PF00008 for LDLR class A repeat, LDLR class B repeat, and EGF-like domain, respectively, are downloaded from Pfam^20^ to generate sequence logo through Skylign^21^. Match emission score from the profile HMMs is used to calculate Δ𝑃𝑓𝑎𝑚 score. Match emission score is the negative log probability to observe the amino acid from multiple sequence alignment for a given position, thus lower score corresponds to high conservation and lower Δ𝑃𝑓𝑎𝑚(𝑟𝑒𝑓 − 𝑎𝑙𝑡) = −(𝑃𝑓𝑎𝑚_*alt*_ − 𝑃𝑓𝑎𝑚_*ref*_) corresponds to higher reference amino acid conservation and lower chance to observe alt amino acid.

### LDLR repeat domain alignment

LDLR class A repeats is aligned as shown in a previous study to align for all Cysteine residues. Alignments for LDLR class B repeats and EGF-like domain were obtained with Clustal Omega^22–24^ by aligning domain sequences with seed alignments from Pfam PF00058 and PF00008.

### UK Biobank data processing

#### Study participants

The UK Biobank^25^ is a prospective cohort of over 500,000 individuals recruited between 2006 and 2010 of ages 40-69. Drawing from 469,803 participants with whole exome sequencing (WES) data, we included 443,353 participants with available LDL cholesterol measurements in this study. Patients with homozygous variants and participants with more than one rare variant across *LDLR* and with any rare variant in *APOB* and *PCSK9* were not considered for these analyses.

#### Variant inclusion and quality control

Exon coordinates were determined for *LDLR*, *APOB*, and *PCSK9* using MANE transcripts^26^, with an additional 5nt retained upstream and downstream of each coding region to capture splice-site variants. Exome sequencing was performed for UKB participants as previously described. Analysis was conducted on the Research Analysis Platform (ukbiobank.dnanexus.com). We extracted gene-level VCF files from the WES joint-called pVCFs using bcftools^27^ (v1.15.1) using the Swiss Army Knife app, then normalized to flatten multiallelic sites and align variants to the GRCh38 reference genome.

Variants in low complexity regions, segmental duplications, or other regions known to be challenging for next generation sequencing alignment or calling were removed from analysis (National Institute of Standards and Technology Genome in a Bottle Consortium^28^ difficult regions), as were variants with an alternate allele frequency greater than 0.1% in the UK Biobank cohort. Further filtering removed variants in which more than 10% of samples were missing genotype calls and variants that did not appear in the UK Biobank cohort. To mitigate differences in sequencing coverage between individuals who were sampled at different phases of the UK Biobank project, variants were only retained in the final set if at least 90% of their called genotypes had a read depth of at least 10.

The canonical functional consequence of each variant was calculated using Variant Effect Predictor (VEP, v99)^29^. Non-coding variants outside of essential splice sites were not considered in the analysis.

Computational scores are provided by VEP, including the PhastCons conservation score^30^ (PhastCons100way_vertebrate). When multiple PhastCons conservation scores are available for a coding variant, the mean of the available scores was used.

#### Clinical endpoints and endophenotypic data

Coronary artery disease and myocardial infarction cases were aggregated from hospital records (primary or secondary diagnosis), death registries (primary or secondary cause of death), and self-reported data. Age of onset was estimated based on date of onset and birth date when not directly provided, and cases with uncertain or unavailable onset data were excluded.

Patient-level LDL-C values were ascertained from the UK Biobank data files. Estimated untreated LDL-C levels were obtained using adjustments for lipid-lowering therapies were used in analyses, as described in the supplement of this manuscript.

#### ClinVar assertions

ClinVar clinical assessments were identified from the tab delimited version of ClinVar released on 04/04/2023. In this analysis, we use ‘pathogenic’, ‘likely pathogenic’, and ‘pathogenic/likely pathogenic’ classifications as ‘P/LP’ collectively, and ‘benign’, ‘likely benign’, and ‘benign/likely benign’ classifications as B/LB.

### BEAN-FUSE scores

We make use of the FUSE (Functional Substitution Estimation) pipeline^31^ to improve the estimation of variant functional effects and to impute effects of variants which have not been screened. FUSE makes use of related measurements within and across experimental assays to jointly estimate variant impacts. After functional scores have been estimated by the BEAN pipeline, the full set of scores are processed by FUSE, which first collectively estimates the mean functional effect per amino acid residue position within the assay, using shrinkage estimation. FUSE then makes estimates for individual allelic variants within the amino acid residue position, based on a functional substitution matrix derived from deep mutational scanning data across many genes. The result is a full set of estimated variant functional effects for both: 1) the original variants screened in the assay, and 2) other possible variants which were not screened but fall within amino acid residues which had variants covered in the screen.

### Prediction of UKB LDL-C level

UKB LDL-C level of variants observed in the base editing and had high confidence (𝜎_μ_ < 0.5) was predicted using XGBoost^32^ Python package and with default option with 10-fold cross validation implemented in scikit-learn^33^ ‘model_selection.cross_val_predict’. The LDL-C levels of UKB variants that are unobserved or not observed with enough confidence (𝜎_μ_ > 0.5) was predicted by the XGBoost model that is trained on the variants observed with 𝜎_μ_ < 0.5.

### Structural analysis

The protein structures were visualized and the screenshots were generated using PyMOL^34^ (v2.5.2). Relative solvent accessibility (RSA)^35^ and residue depth were calculated using the DSSP module in BioPython (v1.79)^36^ to capture the local 3D accessibility of residues. The wild-type atomic interactions between residues were calculated by Arpeggio using the LDLR AlphaFold2 structure (position 1-860) from the AlphaFold Protein Structure Database^37^. Additionally, interactions with calcium ions and saccharides were calculated using PDB structure 1N7D (position 65-714 after renumbering according to Uniprot P01130). These interactions were also computed for mutant structures generated using MODELLER^38^ 10.3. Subsequently, the change in interactions was determined by subtracting the interactions in the mutant from those in the wild-type. Within each of the LDLR class B, LDLR class A, and EGF-like domains, two-sided Wilcoxon rank-sum tests were conducted to compare the features calculated for deleterious variants (identified by BEAN z-scores below -1.96) against those of other variants. DDMut^39^ is a deep learning model that predicts protein stability change induced by mutation, ΔΔ𝐺, based on the local atomic environment and interactions in wild type and mutated residue. LDLR Alphafold2 structure is used as the input to predict the ΔΔ𝐺 of variants observed in our LDLR tiling screen.

Molecular interactions were visually represented using a color-coded scheme to differentiate between interaction types. Hydrophobic interactions were depicted in ’Forest’; polar interactions were depicted in ’Orange’; Carbonyl interactions were depicted in ’Blue’; hydrogen bonds were depicted in ’Red’; Aromatic ring interactions including Methionine Sulfur-π, Donor-π, Cation-π, and Amide-Ring interactions, were depicted in ’Pale Green’; Undefined interactions were depicted in ’Cyan’; Coordinate covalent bonds were depicted in ’Purple’; and Ionic interactions were depicted in ’Yellow’. Moreover, line type specifies distance flag from Arpeggio output, where thin dashed dashed lines represent Van der Waals (vdW) Clashes, where the vdW radii between two atoms cause steric clashes. When such clashes co-occurred with other interactions mentioned earlier, they were portrayed with dashed lines, using the color code corresponding to the additional interaction except for undefined interaction type. Additionally, all ionic interactions and coordinate covalent bonds with ions were consistently represented by yellow and purple dashed thick lines. Other interactions were all represented by solid lines.

## Acknowledgments

The authors thank Grigoriy Losyev and Allison James for technical assistance and funding from UM1HG012010 (R.I.S., L.P.), 1R01HL164409 (C.A.C., R.I.S., L.P.), 1R01GM143249 (R.I.S.), R01HG010372 (C.A.C., T.Y.), American Cancer Society (R.I.S.), American Heart Association (R.I.S.), National Organization for Rare Diseases (R.I.S.), 1R35HG010717-01 (L.P.), National Health and Medical Research Council of Australia (GNT1174405; D.B.A. and Y.Z.), and the Victorian Government’s Operational Infrastructure Support Program (Y.Z. and D.B.A.). We are indebted to the UK Biobank and its participants (UK Biobank application #41250 and IRB protocol 2020P002093). We thank Qian Qin, Cameron Smith, and Logan Blaine for help on BEAN model implementation and representation, Kendell Clement for advice on CRISPResso2 usage, Zain Patel for insights on transcription factor binding analysis, Soojung Yang for advice in structural analysis, and Hanna Boen for helping endogenous editing comparison.

## Author contributions

R.I.S. conceived the experimental design and J.R. and L.P. conceptualized BEAN. S.B. collected screen data. J.R. developed BEAN and M.J., M.I.L. and L.P. advised on design and implementation of BEAN. J.R. and T.Y. processed and analyzed data. T.Y. performed BE-Hive and FUSE analysis. M.F., Q.V.P., and R.I.S. performed downstream characterization of LDL-C GWAS variants. T.Y., L.B., and C.A.C. obtained and analyzed UKB data. Y.Z. led structural analysis of LDLR variants with J.R. and D.B.A. J.R. and Z.L. benchmarked classification performance. M.T. and L.P. performed analysis on variant impact on transcription factor binding. G.L. advised on library design. J.R. and R.I.S. drafted the manuscript. R.I.S, L.P., and C.A.C. provided guidance and supervised this project. All the authors wrote and approved the final manuscript.

## Code availability

Bean source code is available at https://github.com/pinellolab/crispr-bean. The scripts used to generate the figures and analyses presented in the manuscript have been deposited here: https://github.com/pinellolab/bean_manuscript.

## Data Availability

The data used in this manuscript have been provisionally deposited on Zenodo (doi: 10.5281/zenodo.8270605). Controlled access, patient-level data from the UKB may be requested at https://ams.ukbiobank.ac.uk/ams/

## Additional information

Supplementary Information is available for this paper. Correspondence and requests for materials should be addressed to (C.A.C.), (R.I.S.), (L.P.).

