## Supplementary Notes for "Joint genotypic and phenotypic outcome modeling improves base editing variant effect quantification"

**Supplementary Note 1. Experimental design decisions from simulation results**

We have developed reporter screen simulation software (https://github.com/pinellolab/screen-simulation) to guide experiment and computational model design. The software simulates the gRNA read counts given the number of cells per gRNA, gRNA editing rate distribution, variant effect sizes distribution, cell sorting quantile cutoffs, and number of reads per sample. Given the parameters, variant editing, cellular phenotype, and cell sorting are simulated. From the samples of sorted cells, PCR is simulated where gDNA is sampled and amplified. gRNA editing rate is sampled per target variant from the observed reporter editing rate distribution of LDL-C GWAS library, where a target variant in LDL-C GWAS library is sampled at a time to assign 5 gRNA editing rate values to the gRNA targeting a variant in the simulation. This reporter editing rate is then scaled by the sampled endogenous target site accessibility as in CRISPR-BEAN. For each cell with a gRNA, it is edited for the target variant with the probability of the simulated editing rate. If the edit is installed, the phenotype of the cell will be shifted of its mean by the effect size, from the standard normal distribution. Cells will then be sorted into the given phenotypic quantile cutoffs.

In order to study the single-nucleotide variants that have lower phenotypic changes than knock-down by Cas9 or transcriptional activation/deactivation by CRISPRa/i, we hypothesized that the finer partitioning of cell population into more phenotypic quantile bins will enhance the detection of variants with smaller effect size.

Specifically, 700 target variants which are each targeted by 5 distinct gRNAs are analyzed, and 100 variants have nonzero effect sizes of 0.1, 0.2, …, 1.0, 10 per effect size value, and the rest of the 600 variants have 0 effect size. 10 simulations of each condition are aggregated to calculate the sensitivity of detecting variants of each nonzero effect size value.

We showed that sorting into four 20% quantile bins (0-20, 20-40, 60-80, 80-100%) consistently shows better recall compared to traditional top/bottom sorting (0-30, 70-100%) given the same total read depth of 1M across cell coverage per gRNA and number of experimental replicates from simulated data (**Supplementary Fig. 6**).

**Supplementary Note 2. Fitting endogenous editing rate from loci accessibility and reporter outcome**

BEAN takes account for the observation that endogenous editing rate $\tilde{\boldsymbol{\pi}}$ is roughly proportional to the reporter editing rate $\boldsymbol{\pi}$ and is scaled by the loci accessibility. In the model, BEAN fits a function $f$ that maps the reporter editing rate to the endogenous editing rate by assuming the proportional relationship with error $\widetilde{\pi_{gj}} = f(\pi_{gj}) = c\pi_{gj} + \epsilon_g$ for non-wild-type allele $j$ and the scaling factor $c$ is the function of accessibility.

The scaling factor $c$ is fitted from data by fitting the ratio between the observed nucleotide-level editing rate $\pi_{gi}$ in reporter and endogenous target site editing rate $\widetilde{\pi_{gi}}$ in the mini-tiling screen data. Specifically, linear model of $r = log\left(\frac{\widetilde{\pi_{gi}}+0.05}{\pi_{gi}+0.05}\right)$, $\mathrm{E}[r] = aw + b$ was studied where $w = window\_mean(\log(accessibility\ seq\ signal + 1))$ and $window\_mean$ is the mean log-transformed accessibility signal of 100bp upstream and downstream of the edited nucleotide. ATAC-seq, DNase I hypersensitivity, and LMNB ChIP-seq were tested as the accessibility measures that correlates with endogenous editing activity as reported in a previous study[1]. We observed the ratio $r$ correlates the most with the log-transformed ATAC-seq signal (ENCODE[2] ENCFF262URW.Pearson R=0.540) compared to DNase I hypersensitivity signal (ENCODE[2] ENCFF113VII, R=0.473) or LMNB ChIP-seq signal (ChIP-Atlas[3,4] SRX4654321, R=0.221). Thus, we chose ATAC-seq signal as the accessibility input to BEAN. The coefficients were fitted to be $a = 0.2513, b = -1.9458$ with Python package statsmodels[5]' statsmodels.robust.robust_linear_model.RLM (**Supplementary Fig. 26a**)

The fitted relationship was used to transform allele editing rate in the reporter to the rate in the endogenous editing. To account for the deviation of $r$ from the predicted, $\epsilon_g$ =$logit\left(\sum_{i\neq 0}\widetilde{\pi_{gj}}\right) - logit\left(\sum_{i\neq 0}\pi_{gj}\right)$ is fitted during the inference time per gRNA $g$, and its prior is set to be $\epsilon_g \sim \mathcal{N}(0, \sigma_\epsilon)$

where $\sigma_\epsilon = 0.655$ is the observed standard deviation of the residual $logit(\widetilde{\pi_{gi}}) - logit\left(f(\pi_{gi})\right)$ using the $f$ from the above fitted relationship (**Supplementary Fig. 26b**).

**Supplementary Note 3**. Over-dispersed multinomial count data modeling with Dirichlet-Multinomial distribution

Additionally, we extend the univariate count modeling as Negative Binomial[12,13] or Beta-binomial[14] distribution into multivariate counts of each gRNA in different sorting bins and alleles that is produced by each gRNA as Dirichlet-multinomial distribution. We show from the simulation result that although the gRNA count in each sorted sample is generated using Negative Binomial distribution, Dirichlet-multinomial modeling of gRNA counts across sorting bins shows comparable recall of variants (**Supplementary Fig. 27**).

We follow the DESeq[6,7] procedure of utilizing depth-normalized sample mean and variance to fit the total concentration estimate $\alpha_g^\circ = \sum_k \alpha_g^{(k)}$ for each gRNA $g$ and trend-fitting the concentration estimates with depth-normalized total counts, where $\alpha_g^{(k)}$ is the concentration parameter of Dirichlet-Multinomial distribution $X_g \sim DirMult(n_g, \boldsymbol{\alpha} = (\alpha_g^{(1)}, \dots, \alpha_g^{(d)}))$ of a specific gRNA $g$. Subscript $g$ is omitted below until otherwise described for simplicity.

First, we obtain formula to estimate per-gRNA $\alpha^\circ$ values using method-of-moments. For $X = (X^{(1)}, \dots, X^{(d)})$ and $k \in \{1, \dots, d\}$,

$$\mu^{(k)} = E\left[X^{(k)}\right] = np^{(k)}$$

$$V^{(k)} = Var\left(X^{(k)}\right) = np^{(k)}\left(1 - p^{(k)}\right)\left(1 + \frac{n-1}{1+\alpha^\circ}\right)$$

Where $p^{(k)} = \frac{\alpha^{(k)}}{\alpha^\circ}$. Given $n$, we can get the method-of-moment estimates of $p^{(k)}$and $\alpha^\circ$ as

$$\widehat{p^{(k)}} = \frac{\widehat{\mu^{(k)}}}{n}$$

$$\widehat{\alpha^\circ} = \left( \sum_{k \in \{1 \ldots d\}} \frac{n-1}{\frac{\widehat{V^{(k)}} - \widehat{\mu^{(k)}}}{n\widehat{p^{(k)}}\left(1-\widehat{p^{(k)}}\right)} - 1 + \frac{1}{1-\widehat{p^{(k)}}}} - 1 \right) / d$$

Next, as multiple observations from different replicates does not share $n$ across replicates, we follow DESeq's count normalization procedure to obtain within-group variances and means as follows. Here, $s_r^{(k)}$ is the size factor of the sample in $r^{th}$ replicate and $k^{th}$ sorting bin calculated as in DESeq and described in the Method section of the main text.

$$\widehat{\mu^{(k)}} = \sum_{r=1 \ldots R} \frac{X_r^{(k)} / s_r^{(k)}}{R}$$

$$\widehat{V^{(k)}} = \frac{\sum_{r=1 \ldots R} \left( X_r^{(k)} / s_r^{(k)} - \widehat{\mu^{(k)}} \right)^2}{R}$$

We report overdispersion of counts from comparing within-group variances and and multinomial variances (**Supplementary Fig. 21a, d**). Furthermore, we observed strong linear trend between the normalized total count $\hat{n} = \sum_k \widehat{\mu^{(k)}}$ and $\widehat{\alpha^\circ}$ (**Supplementary Fig. 21b-c, e-f**) in log scale. The linear trend between $\log(\hat{n})$ and $\log(\widehat{\alpha^\circ})$ is fit with `scipy.optimize.curve_fit` of Python package SciPy[8].

$$\tau_g = \log\left(\widehat{\alpha_g^\circ}\right) \sim \beta_0 + \beta_1 \log(\hat{n}_g)$$

$$\log\left(\widehat{\alpha^\circ}_{tr}\left(\hat{n}_g\right)\right) = \widehat{\beta_0} + \widehat{\beta_1} \log(\hat{n}_g)$$

In the manuscript, we take $\widehat{\alpha^\circ}_{tr}\left(\hat{n}_g\right)$ as the final $\alpha^\circ$ estimate. We provide the option to shrink the $\widehat{\alpha_g^\circ}$ for each gRNA $g$ towards the trend-fitted $\widehat{\alpha^\circ}_{tr}\left(\hat{n}_g\right)$ with `--shrink-alpha` argument of `bean-run` that utilizes Normal-Normal conjugacy. Specifically, we consider the trend-fitted concentration estimate $log\left(\widehat{\alpha^\circ}_{tr}\left(\hat{n}_g\right)\right)$ as the mean of the prior distribution of $\kappa_g$, which is the mean of the normal distribution that gives rise to the MoM-fitted $\tau_g = \log\left(\widehat{\alpha_g^\circ}\right)$ for individual gRNA.

$$\kappa_g \sim \mathcal{N}\left(log\left(\widehat{\alpha^{\circ}}_{tr}(\hat{n}_g)\right), \eta\right)$$

$$\tau_g | \kappa_g \sim \mathcal{N}(\kappa_g, \nu)$$

$$\eta = \frac{\sum_{g \in G}\left(\tau_g - log\left(\widehat{\alpha^{\circ}}_{tr}(\hat{n}_g)\right)\right)^2}{|G| - 1}$$

$$\log\left(\widehat{\alpha_g^{\circ}}\right)_{shrink} = E\left[\kappa_g \middle| \tau_g\right] = \frac{\eta}{\nu + \eta}\tau_g + \frac{\nu}{\nu + \eta} log\left(\widehat{\alpha^{\circ}}_{tr}(\hat{n}_g)\right)$$

Here, $G$ is the set of gRNAs.
