## Supplementary Figures for "Joint genotypic and phenotypic outcome modeling improves base editing variant effect quantification"

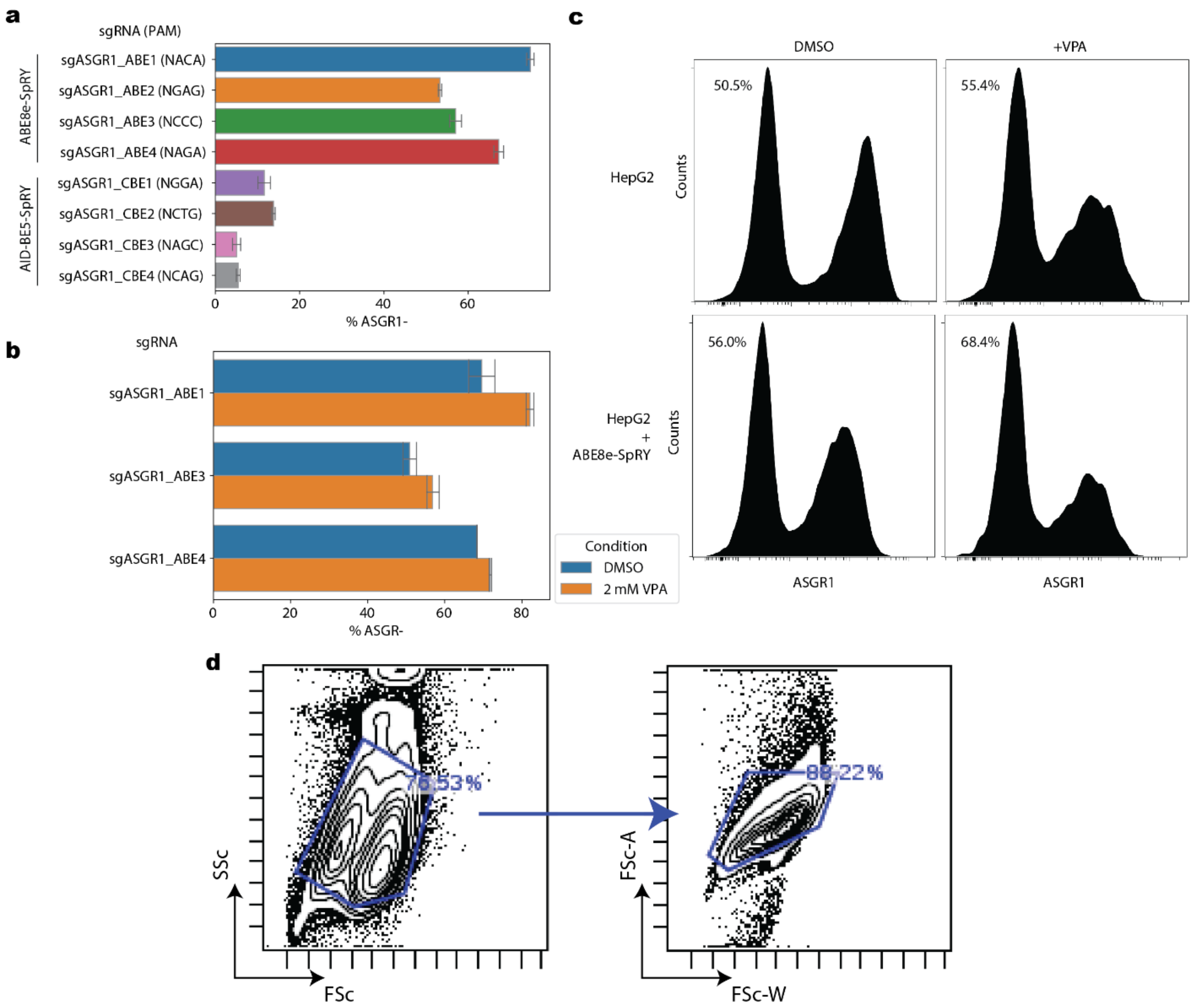


Supplementary Figure 1. Optimization of SpRY base editing. a) gRNA editing efficiency in HepG2 with ABE8e-SpRY and AID-BE5-SpRY for ASGR1 splice site-targeted gRNAs, measured by the fraction of ASGR1 negative cell counts (% ASGR -) quantified by flow cytometry from two experimental replicates. b) gRNA editing efficiency with and without valproic acid (VPA) treatment from 4 experimental replicates. c) Flow cytometry signal with and without stable ABE8e-SpRY integration and valproic acid (VPA) treatment. Fraction of ASGR negative cell counts in each condition is labeled as percentage in the panel. d) Example flow cytometric gates used in all analysis and sorting experiments to filter for single cells.

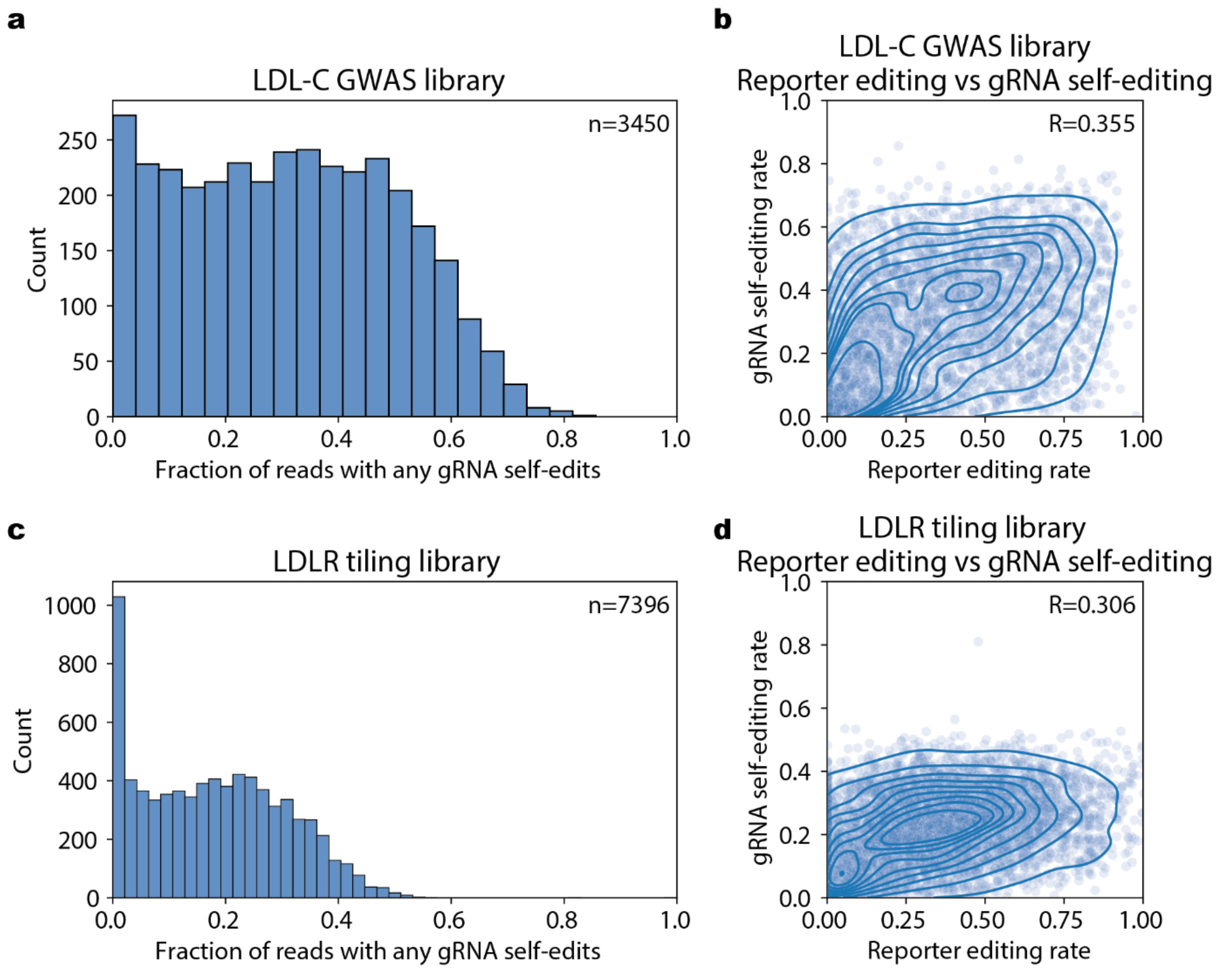


Supplementary Figure 2. gRNA self-editing rates and correlation with reporter editing rate. a) Histogram of LDL-C GWAS library gRNA self-editing rate. b) Correlation of gRNA self-editing with reporter editing rate in LDL-C GWAS library. c) Histogram of LDLR tiling library gRNA self-editing rate. d) Correlation of gRNA self-editing with reporter editing rate in LDLR tiling library. gRNAs with more than 10 counts in the bulk sample of any replicates are considered and any A to G editing in the spacer sequence is considered self-editing. Pearson correlation coefficients are shown in b) and d) as R.

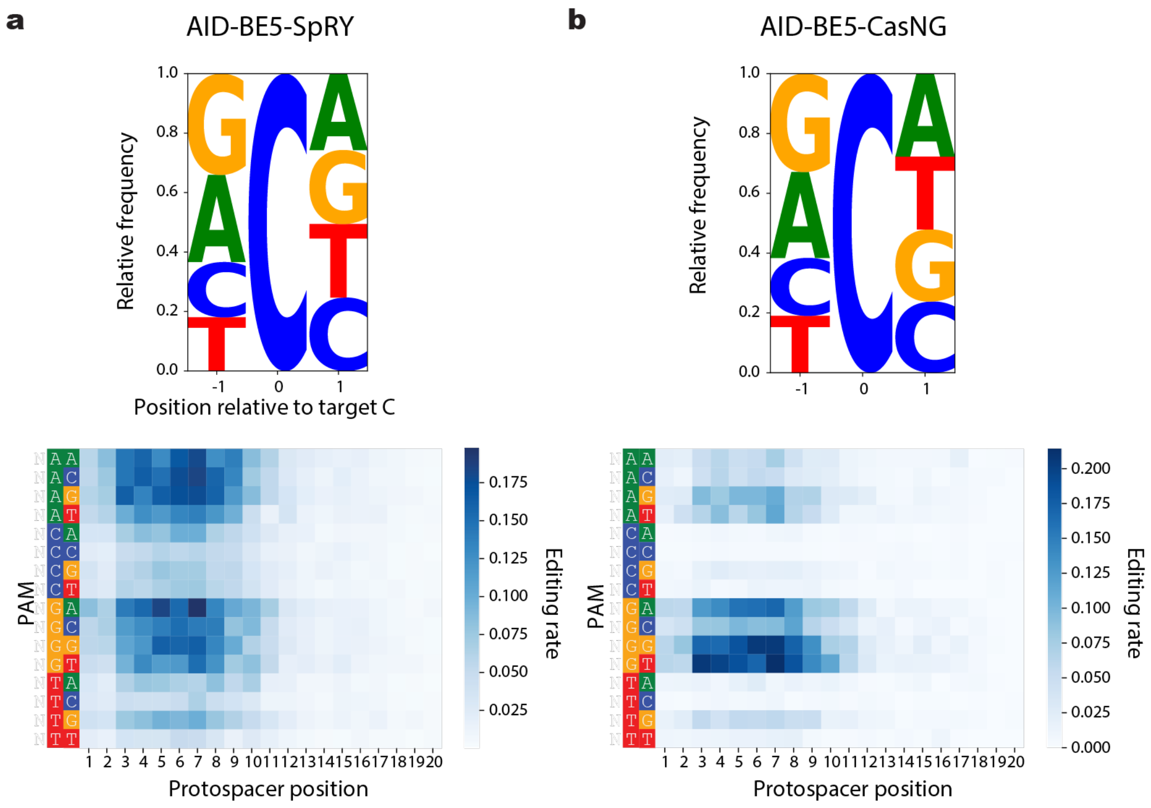


Supplementary Figure 3. Base editor editing preference profile and context specificity. Deamination motif and PAM-dependent editing preference of a) AID-BE5-SpRY from 7294 gRNAs and b) AID-BE5-Cas9NG from 7299 gRNAs with more than 9 read counts across any replicates of bulk samples. Context specificities of two enzymes are represented as sequence logos. The height of each base represents the relative editing efficiency with each base. Mean editing efficiencies by protospacer position and PAM sequence are shown in the bottom panels.

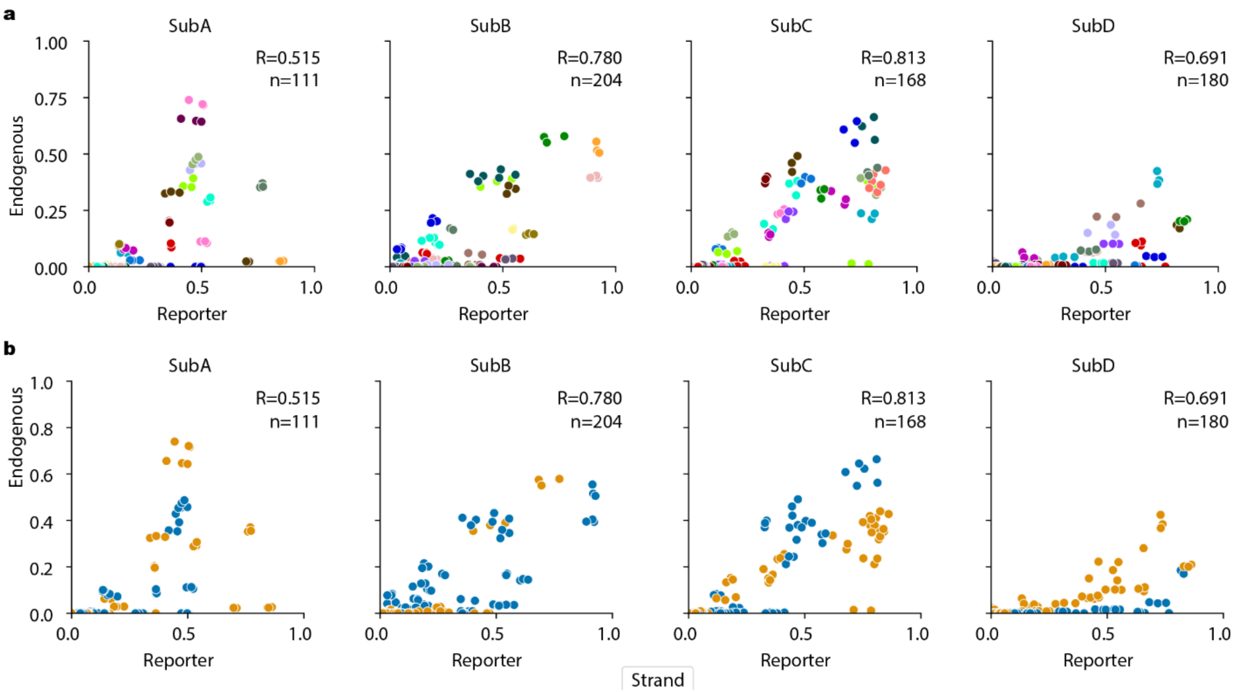


Supplementary Figure 4. Nucleotide-level editing comparison of reporter and endogenous locus. a) Nucleotide level editing rates colored by nucleotide edit per gRNA. Each point plots per-replicate editing rates of nucleotide level edits made by distinct gRNAs. b) The same plot colored by gRNA strand.

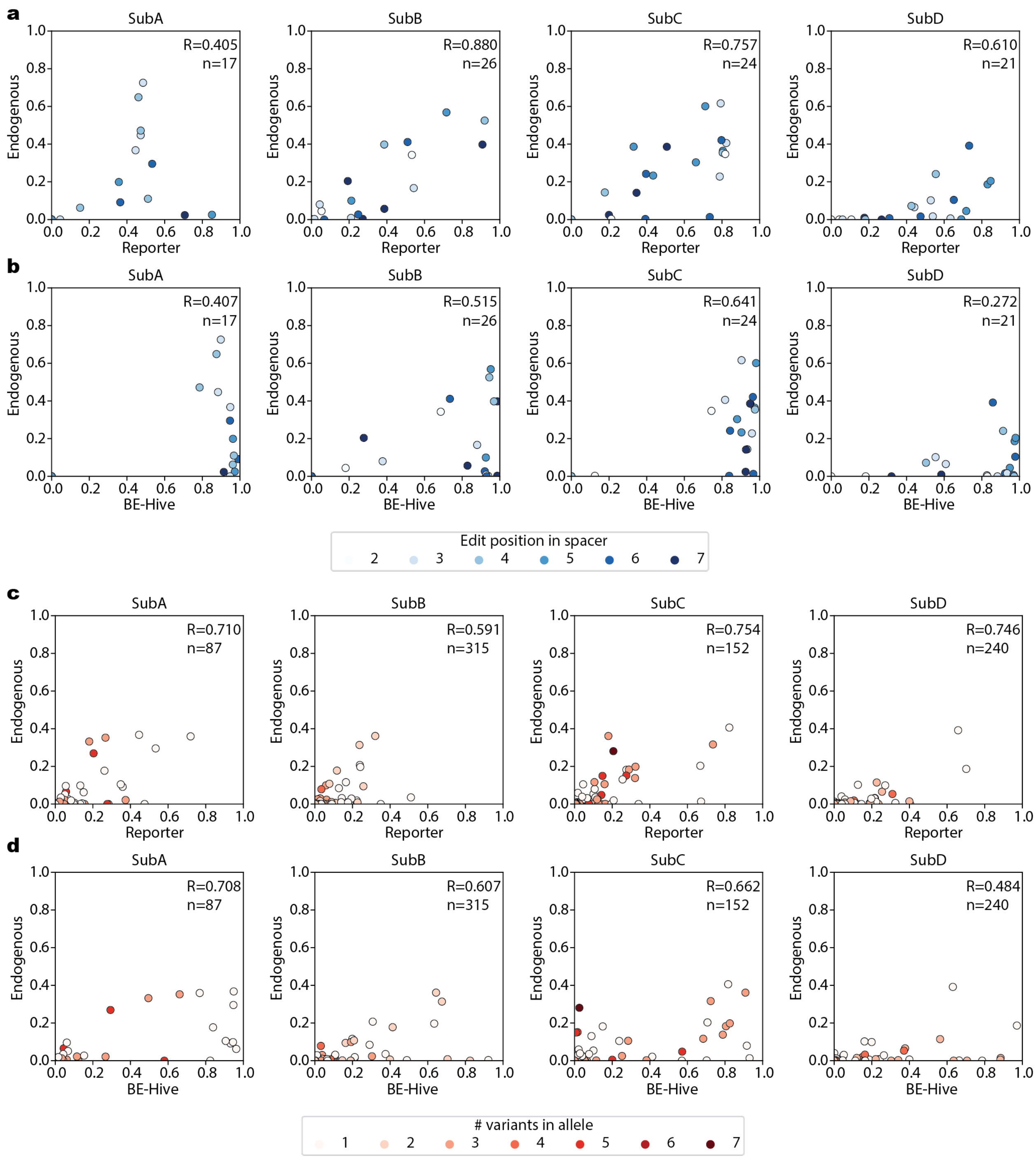


Supplementary Figure 5. Endogenous target site editing rate comparison with reporter and BE-Hive predicted editing outcomes. Mean endogenous and reporter edit rates across three replicates are plotted. a-b) Nucleotide-level editing rates in a) reporter and b) BE-Hive plotted against those in endogenous loci within protospacer position 2 to 7 (1-based, inclusive). b) Allele level editing rates in c) reporter and d) BE-Hive plotted against those in endogenous loci. Alleles are defined within editing window within protospacer 1 to 19 (1-based, inclusive). SubA-D denotes sublibraries A-D. Pearson correlation coefficients are denoted as R and the number of points is denoted as n.

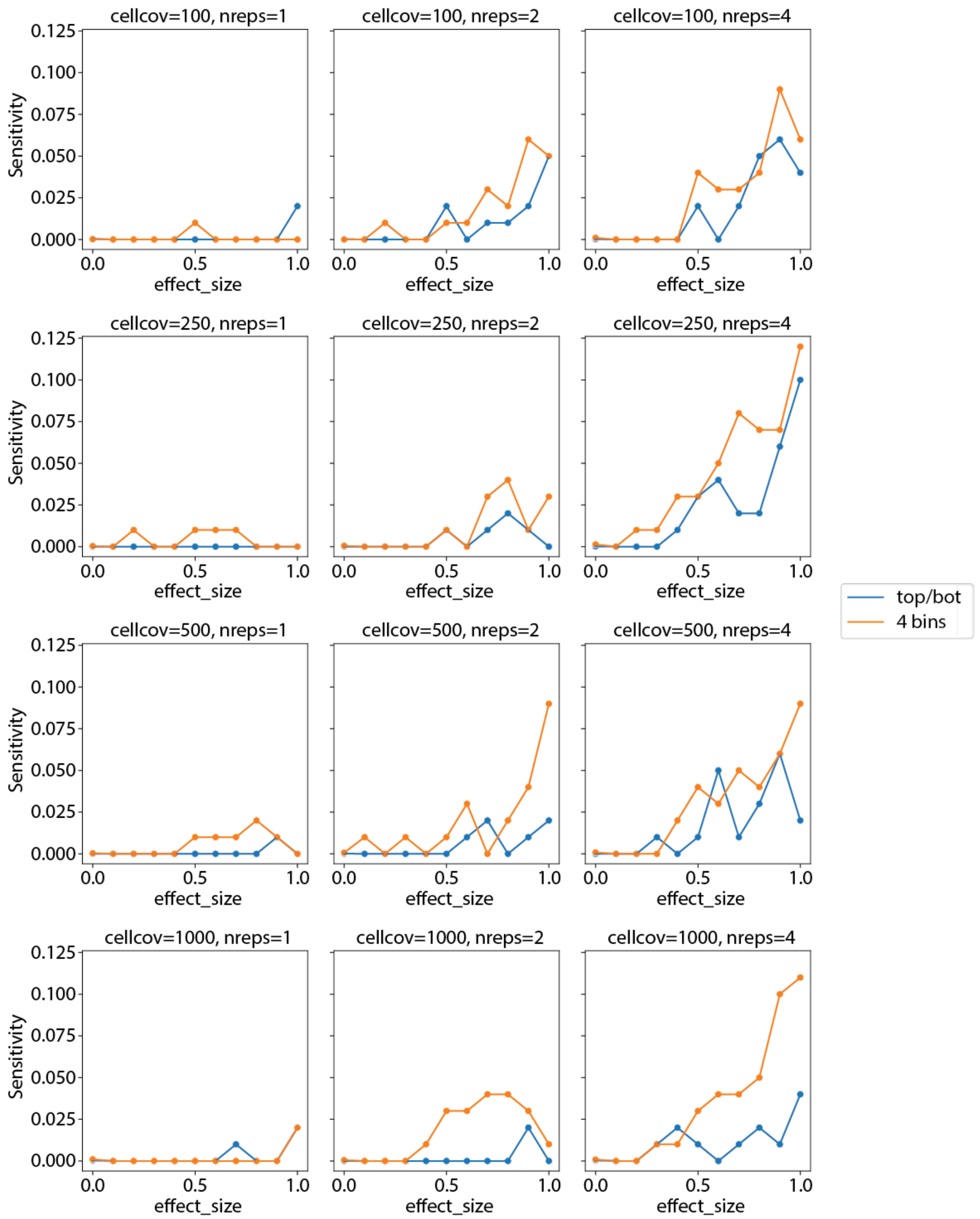


Supplementary Figure 6. Sort bin simulation analysis. Comparison of average recall calculated from 100 variants per effect size across 10 simulation runs. from top/bot sorting scheme (0-30%, 70-100% phenotypic quantiles) to finer 4-bin sorting scheme (0-20%, 20-40%, 60-80%, 80-100% phenotypic quantiles) of cells in simulated data given the same total read depth of 10M. cellcov: Average number of cells that has the gRNA. nreps: Number of simulated experimental replicates. Screen data is analyzed by MAGeCK-RRA paired mode with FDR threshold 0.05.

**a**

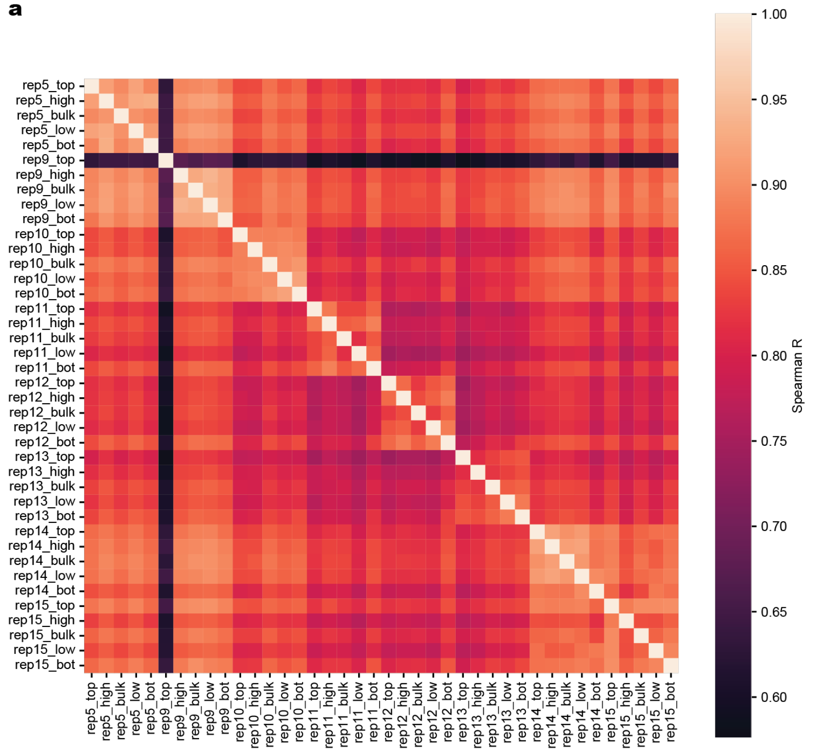


**b**

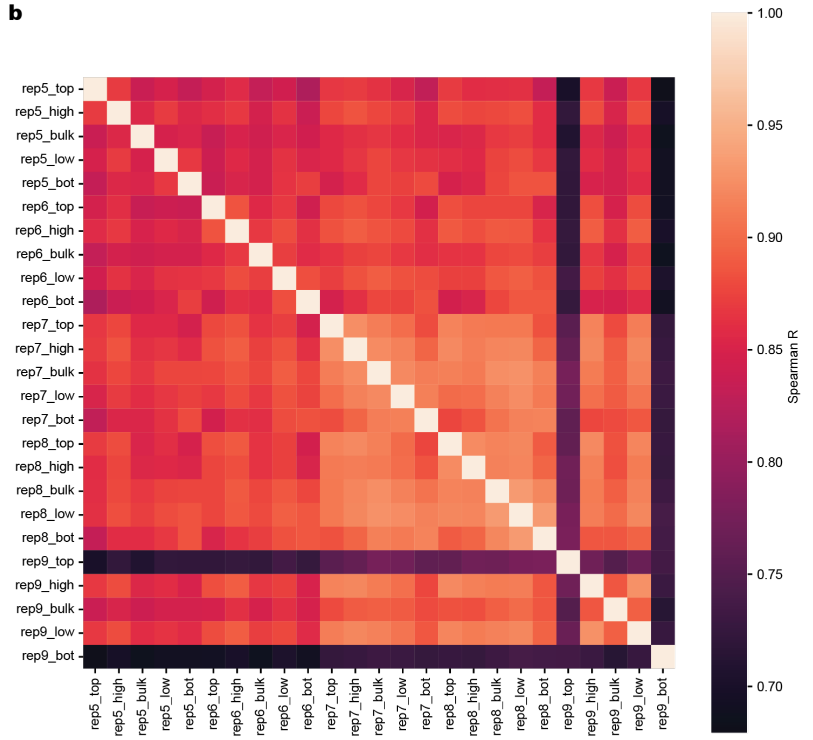


Supplementary Figure 7. Base editing screen read correlations. Spearman correlation coefficient of a) gRNA counts of LDL-C GWAS library and b) LDLR tiling library samples after filtering out for gRNAs with less than 10 read counts. Top; 80-100%, high; 60-80%, low; 20-40%, bot: 0-20% percentile LDL-C uptake. Rep; experimental replicate.

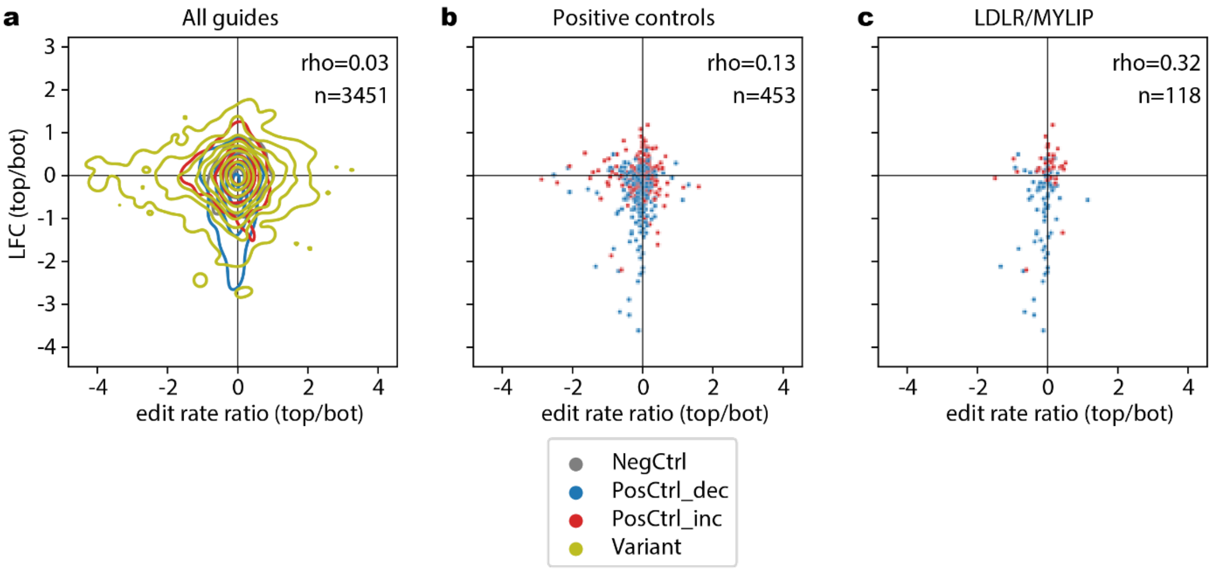


Supplementary Figure 8. Jackpot analysis. Scatterplots of editing rate enrichment plotted against gRNA abundance enrichment of a) all gRNAs, b) positive control gRNAs, c) strongest positive control gRNAs targeting LDLR or MYLIP splicing sites. Enrichment of both editing efficiencies and gRNA abundance is calculated as the log fold chance of the measures in the highest and lowest 20% quantile bin. Spearman correlation coefficients are shown as rho. NegCtrl; negative control gRNAs, PosCtrl_dec; positive control gRNAs that is expected to decrease LDL-C uptake, PosCtrl_inc; positive control gRNAs that is expected to increase LDL-C uptake.

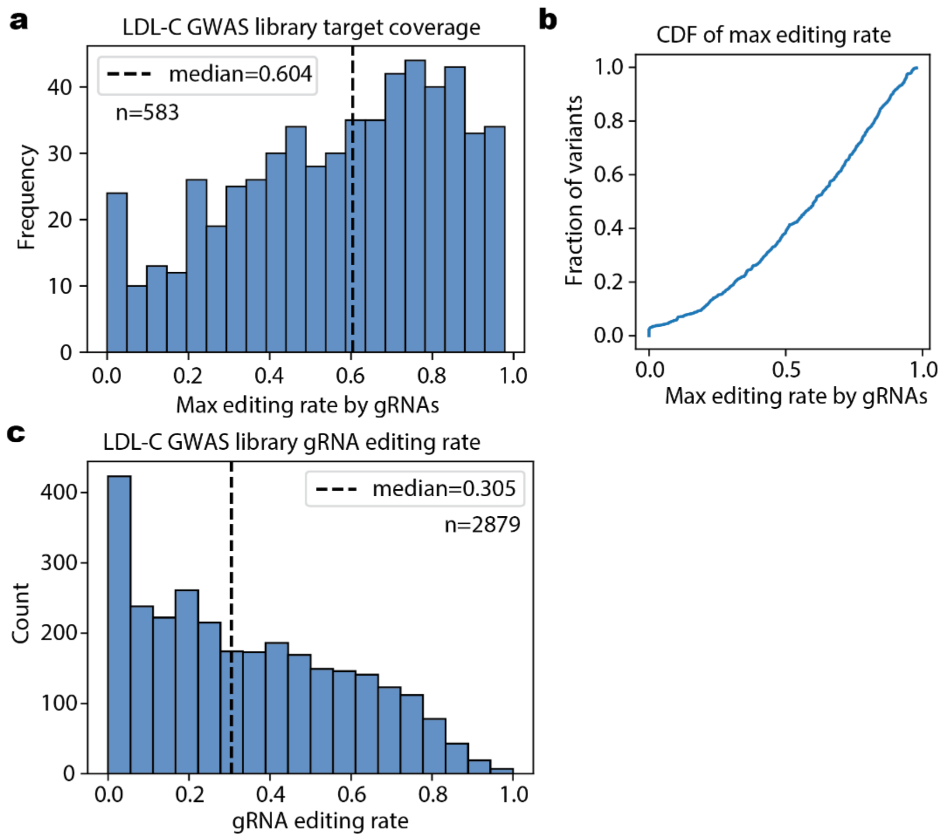


Supplementary Figure 9. LDL-C GWAS variant editing coverage. a) Histogram of per-target variant coverage calculated by the maximal editing rate of the variant by any gRNAs targeting the variant, where gRNA editing rates are calculated as the mean editing rate in bulk samples across replicates where more than 10 reads are observed. b) Cumulative distribution function of the same distribution plotted in a). c) Per-gRNA editing rate of the target variant in bulk samples across replicates where more than 10 reads are observed.

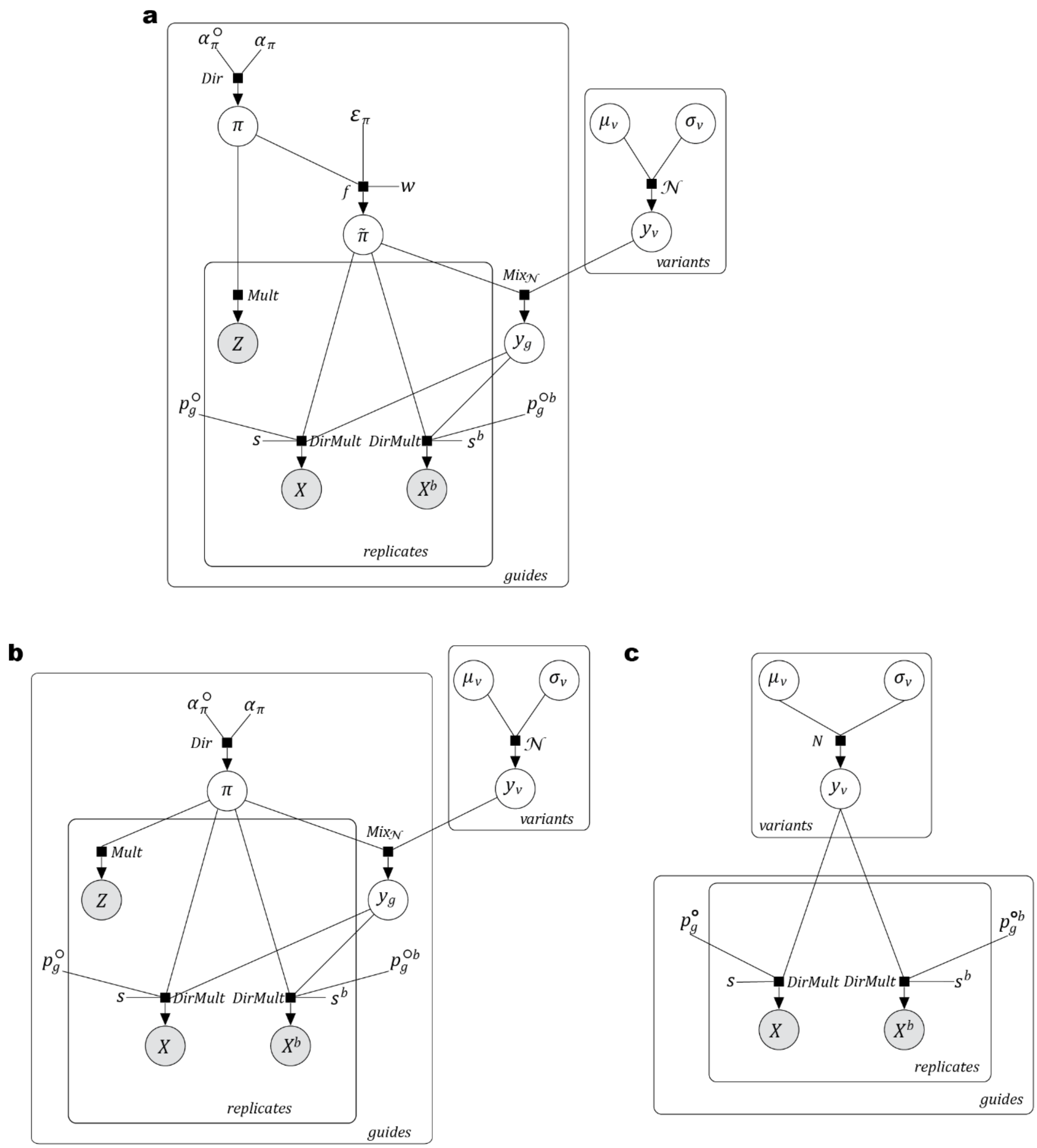


Supplementary Figure 10. BEAN modeling logic. Plate diagrams of a) BEAN b) BEAN-Reporter, c) BEAN-Uniform. $X^b$ and all parameters with superscript $b$ is not used for benchmark analyses.

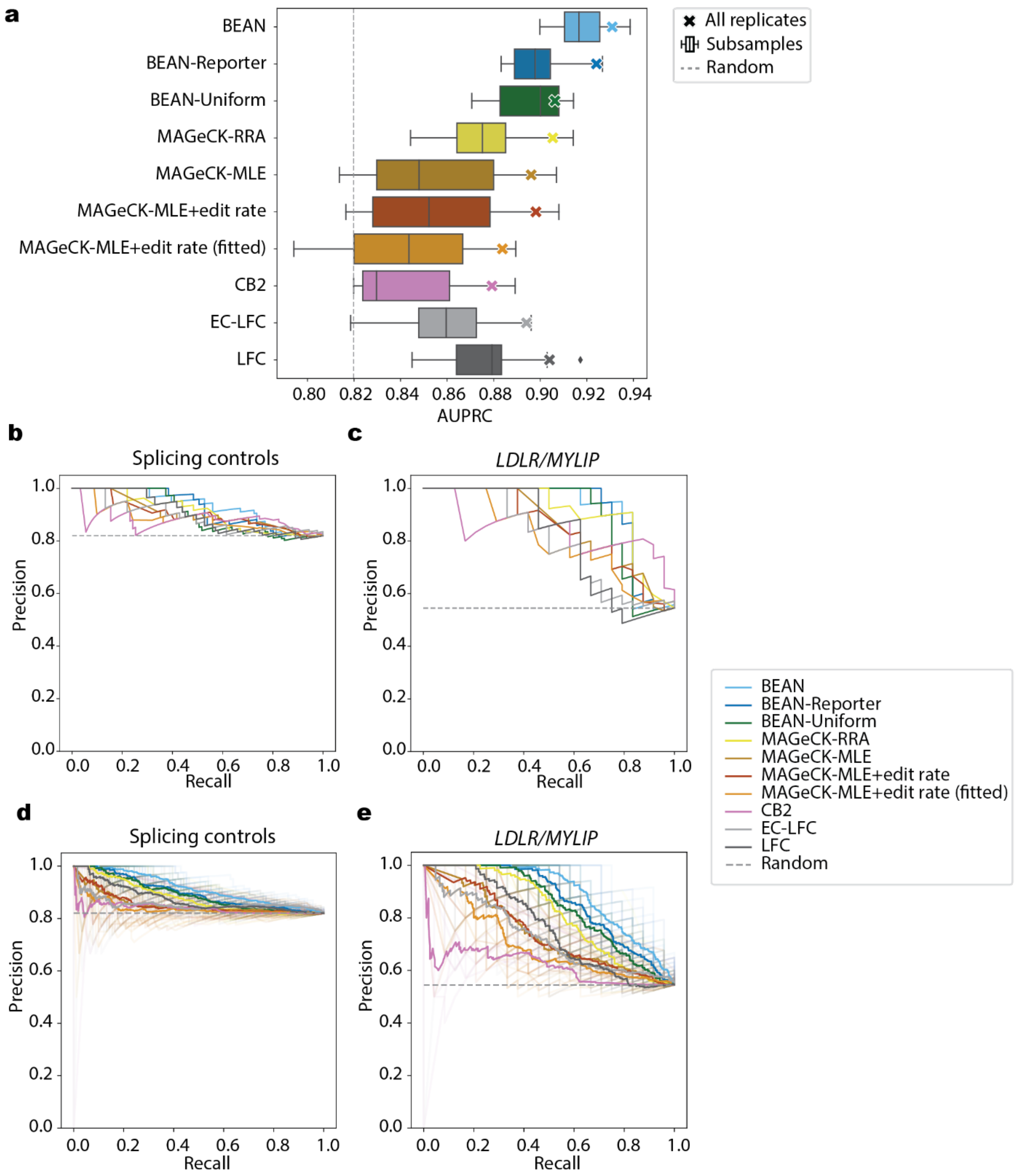


Supplementary Figure 11. LDL-C GWAS library quality control analysis. a) AUPRC plot for classifying positive splicing control variants against negative control variants. b) Precision-Recall curve for classifying all positive control splice sites of against negative controls for all replicates with no failing samples. c) Precision-Recall curve for classifying splice sites of *LDLR/MYLIP* against negative controls for all replicates with no failing samples. d) Precision-Recall curve for classifying all positive control splice sites of against negative controls for 2-replicate subsample of the data. Mean Precision value for a recall across 15 subsample runs are plotted as solid line. e) Precision-Recall curve for classifying splice sites of

*LDLR*/*MYLIP* against negative controls for 2-replicate subsample of the data. Mean Precision value for a recall across 15 subsample runs are plotted as solid line.

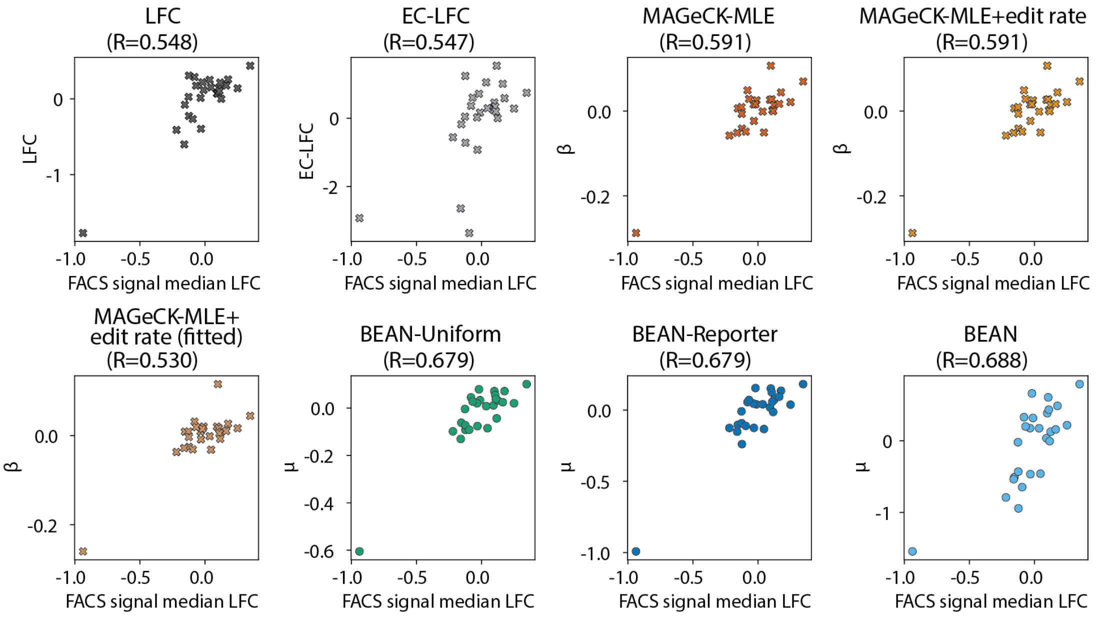


Supplementary Figure 12. Comparison of LDL-C uptake from distinct LDL-C GWAS screen analysis approaches and individual gRNA testing. Scatterplot and Pearson correlation coefficients (R) of effect size estimates and the log fold change (LFC) of fluorescence signal following individual transfection of 22 gRNAs.

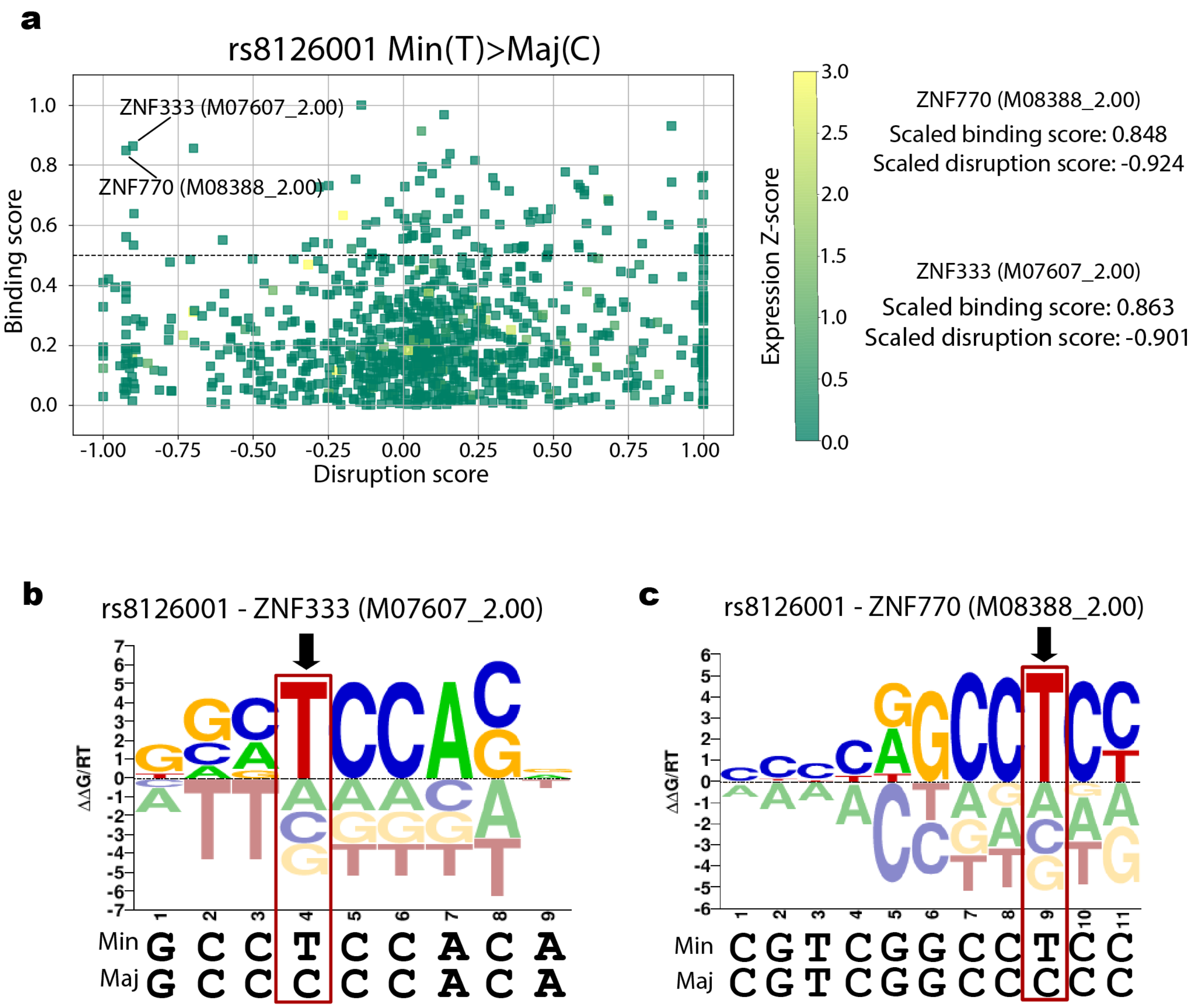


Supplementary Figure 13. MotifRaptor analysis of candidate variant transcription factor binding disruption. a) Disruption plotted against binding score of motifs of rs8126001 minor allele transition to major. b-c) Identified ZNF333 and ZNF770 motif aligned with rs8126001 loci with minor and major alleles.

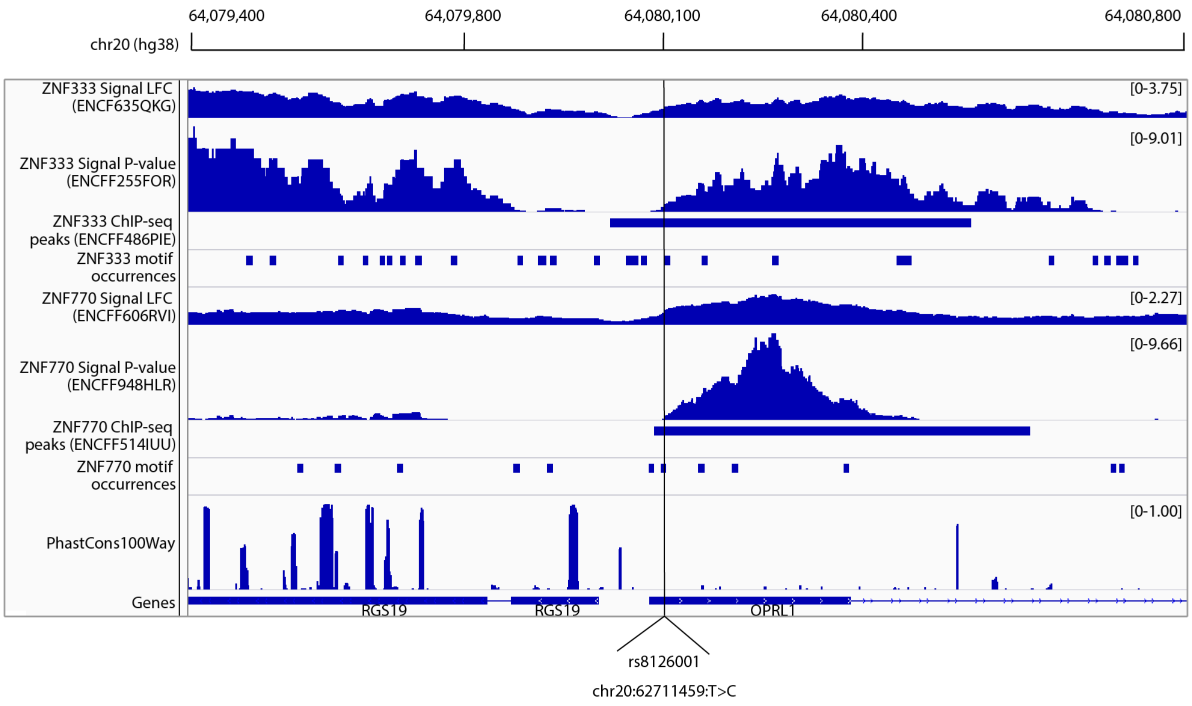


Supplementary Figure 14. ChIP-seq signal LFC, signal P-values, peaks and motif occurrences of ZNF333 and ZNF770 around rs8126001. PhastCons100way conservation scores and gene annotations are displayed together. ENCODE accessions are shown in parenthesis.

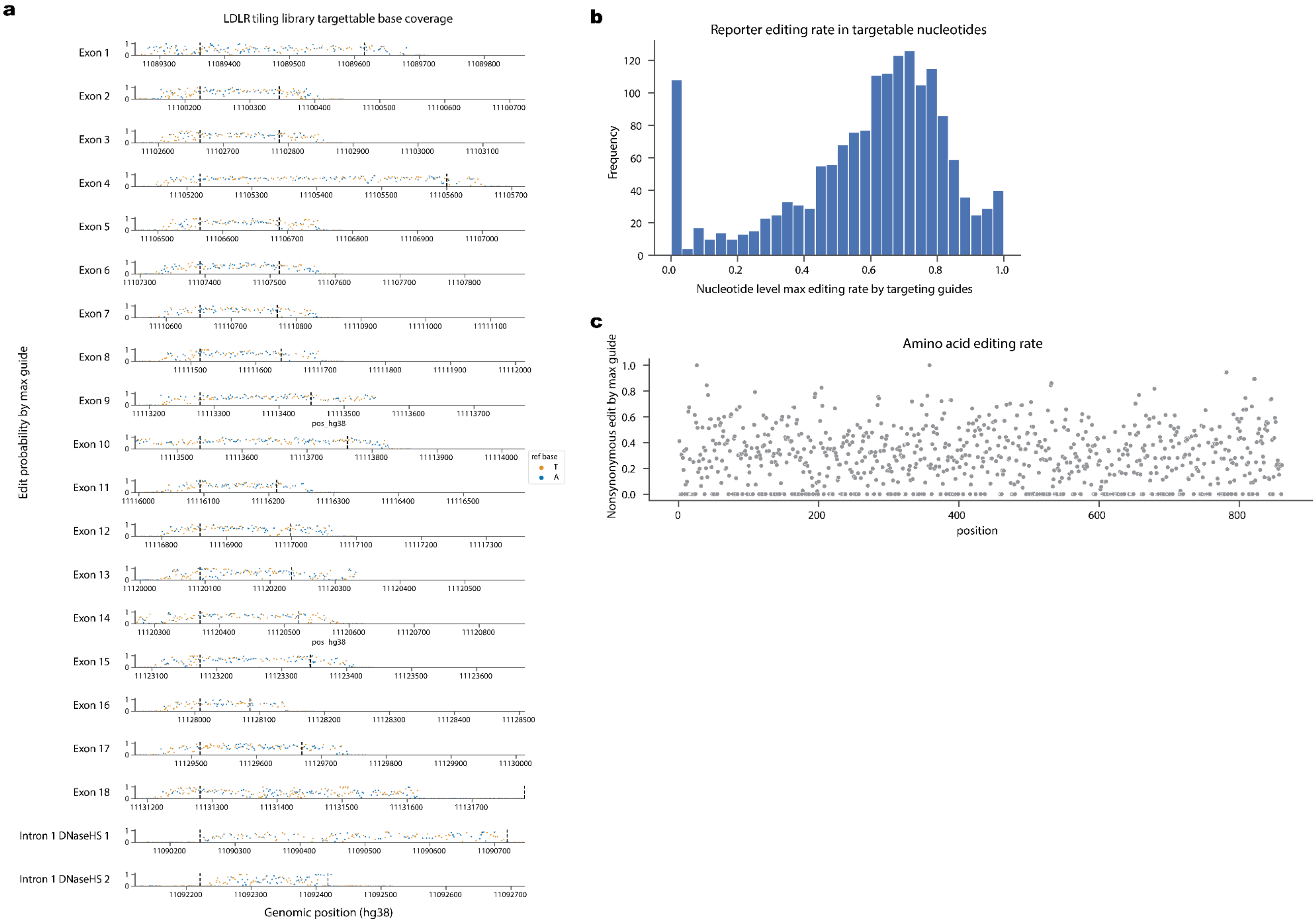
a
LDLR tiling library targettable base coverage
Exon 1
Exon 2
Exon 3
Exon 4
Exon 5
Exon 6
Exon 7
Exon 8
Exon 9
Exon 10
Exon 11
Exon 12
Exon 13
Exon 14
Exon 15
Exon 16
Exon 17
Exon 18
Intron 1 DNaseHS 1
Intron 1 DNaseHS 2
Edit probability by max guide
Genomic position (hg38)
pos_hg38
ref base
T
A
b
Reporter editing rate in targetable nucleotides
Frequency
Nucleotide level max editing rate by targeting guides
c
Amino acid editing rate
Nonsynonymous edit by max guide
position

Supplementary Figure 15. *LDLR* tiling library reporter editing coverage. For each nucleotide or amino acid, maximum editing by gRNA targeting the nucleotide or amino acid is plotted. a) Per-position nucleotide editing for targetable bases by ABE. b) Histogram of nucleotide level editing rate plotted in a). c) Nonsynonymous amino acid editing after filtering for alleles that appear in >5% frequency in any samples.

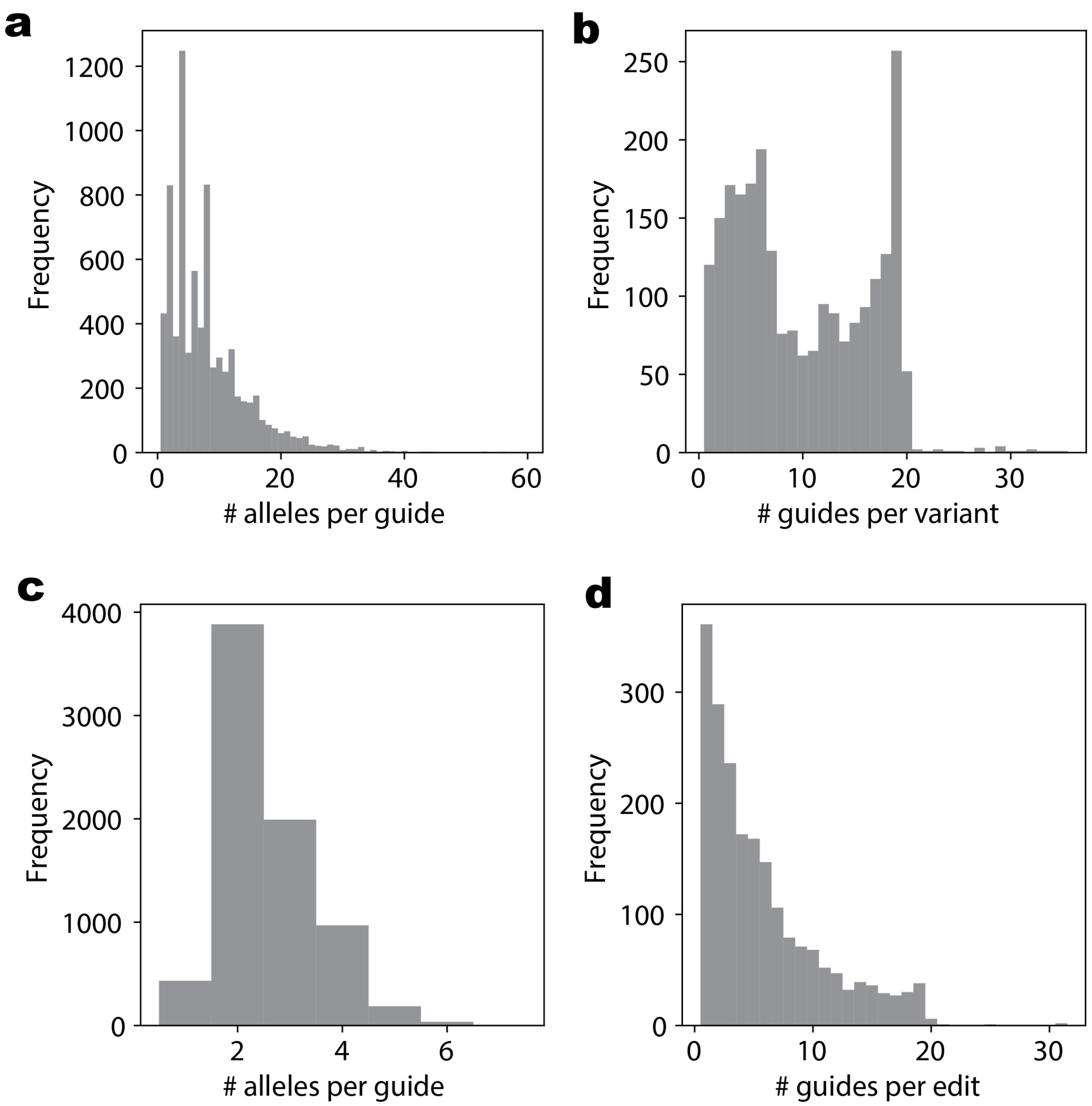


Supplementary Figure 16. *LDLR* tiling library allele representation statistics before and after allele translation and filtering. a) Histogram of number of translated alleles per gRNA, including wild type alleles, when filtered by significance compared to the plasmid library. Only the variants located in the coding sequence are translated into amino acid level variants and variants in the noncoding sequence remains as the nucleotide-level variants. b) Histogram of number of gRNAs that produces each variant that occurs in allele in a). c) Histogram of number of translated alleles per gRNA, including wild type alleles, after filtering alleles in a) by allele fraction of >10% in >30% of the replicates. d) Histogram of number of gRNAs that produces each variant that occurs in allele in c).

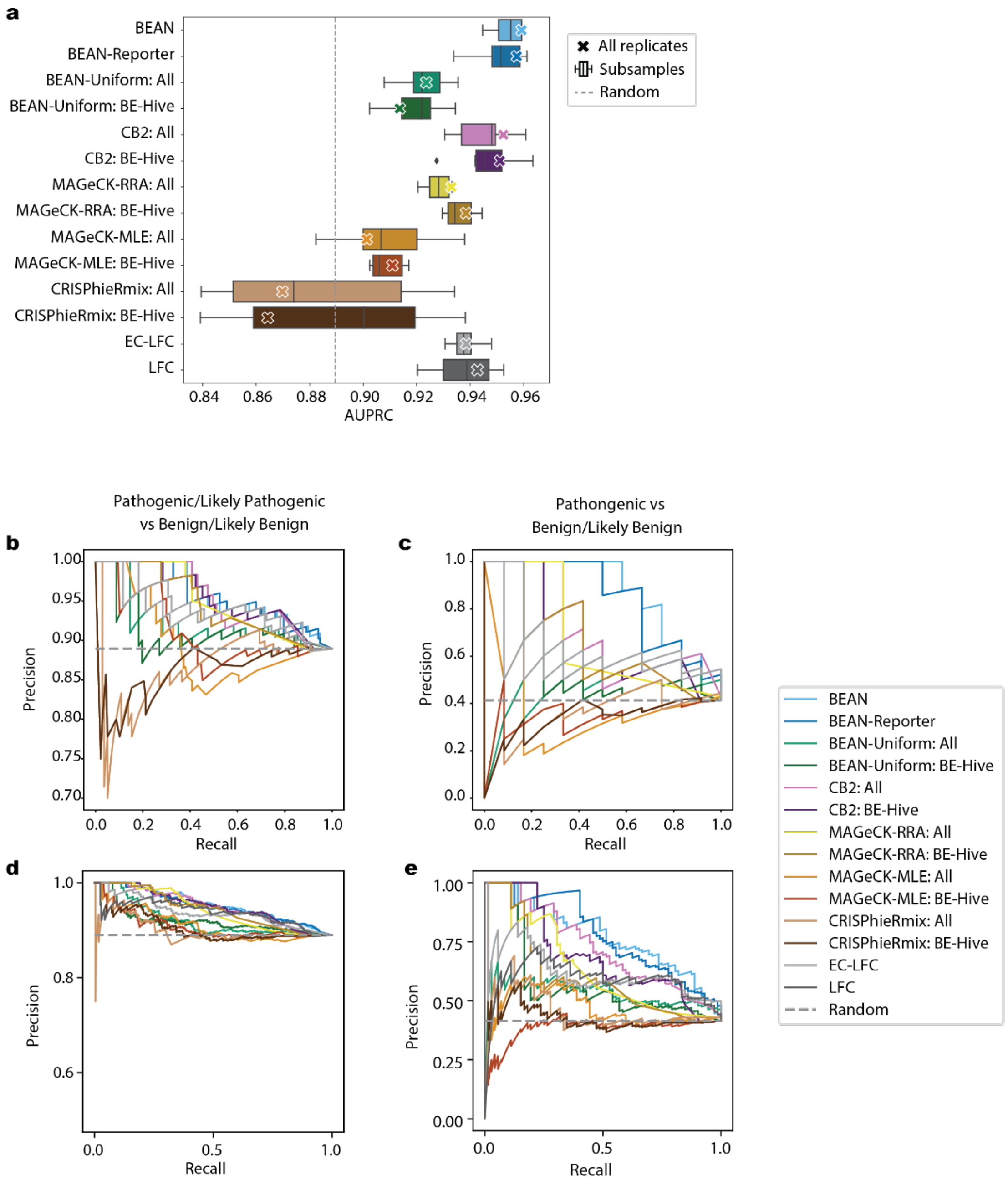


Supplementary Figure 17. *LDLR* tiling library quality control analysis. a) AUPRC of classifying Pathogenic/Likely Pathogenic from Benign/Likely Benign variants when using 4 replicates without failing samples and 6 2-replicates combinations among the replicates. b-c) Precision-recall curve of classifying b) Pathogenic/Likely Pathogenic c) Pathogenic from Benign/Likely Benign variants. Top panels show classification when used 4 replicates without failing samples. Bottom panels show when used 6 2-replicates combinations among 4 replicates without failing samples.

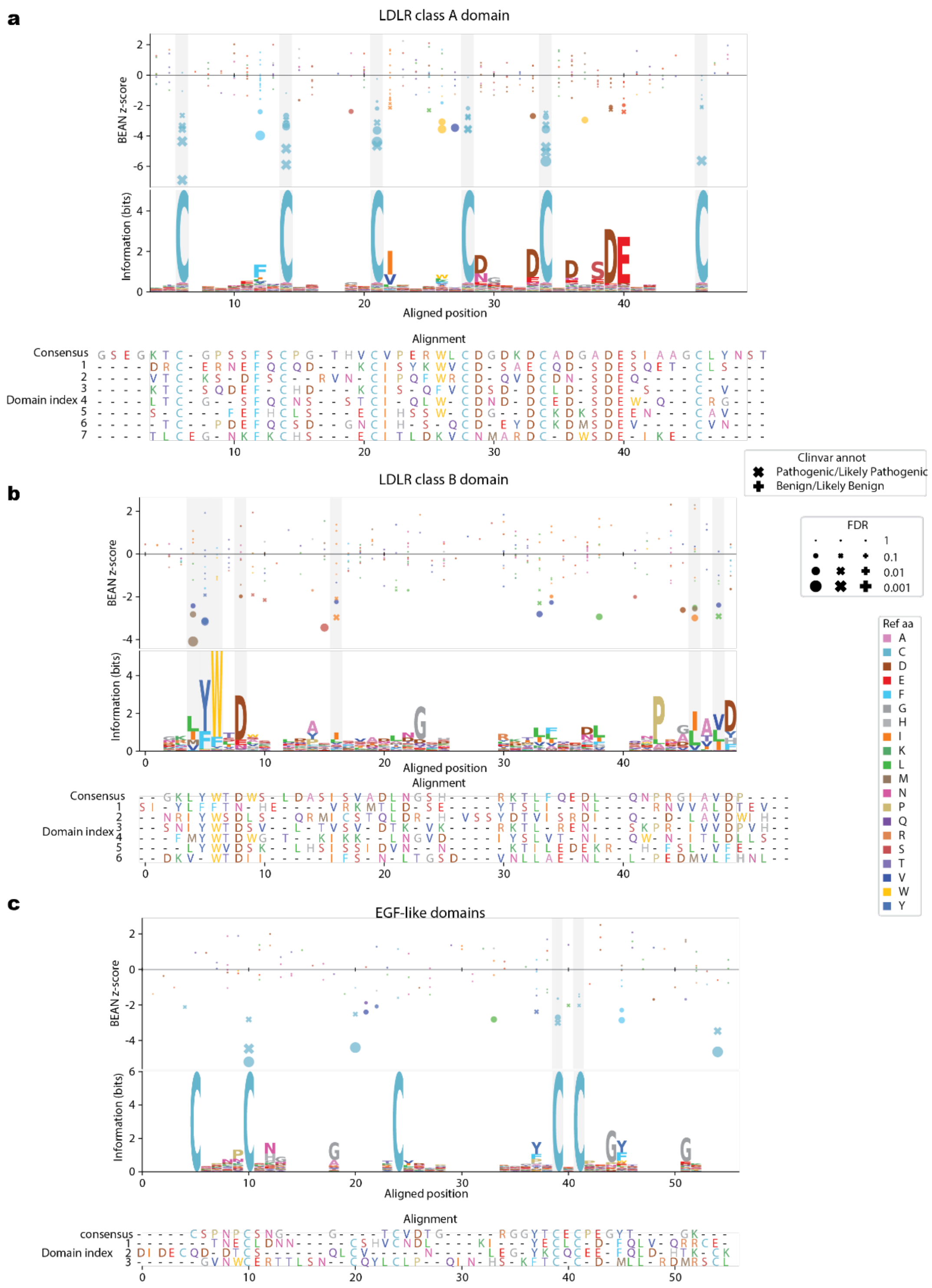


Supplementary Figure 18. Comparison of functional impact and conservation within conserved LDLR domains. Repeat domain alignments shown with BEAN z-score for a) LDLR class A repeat domain, b) LDLR class B repeat domain, c) EGF-like domains. For a), conserved cysteine residue position is highlighted and for b-c), consensus positions from Clustal Omega alignment output is highlighted in grey.

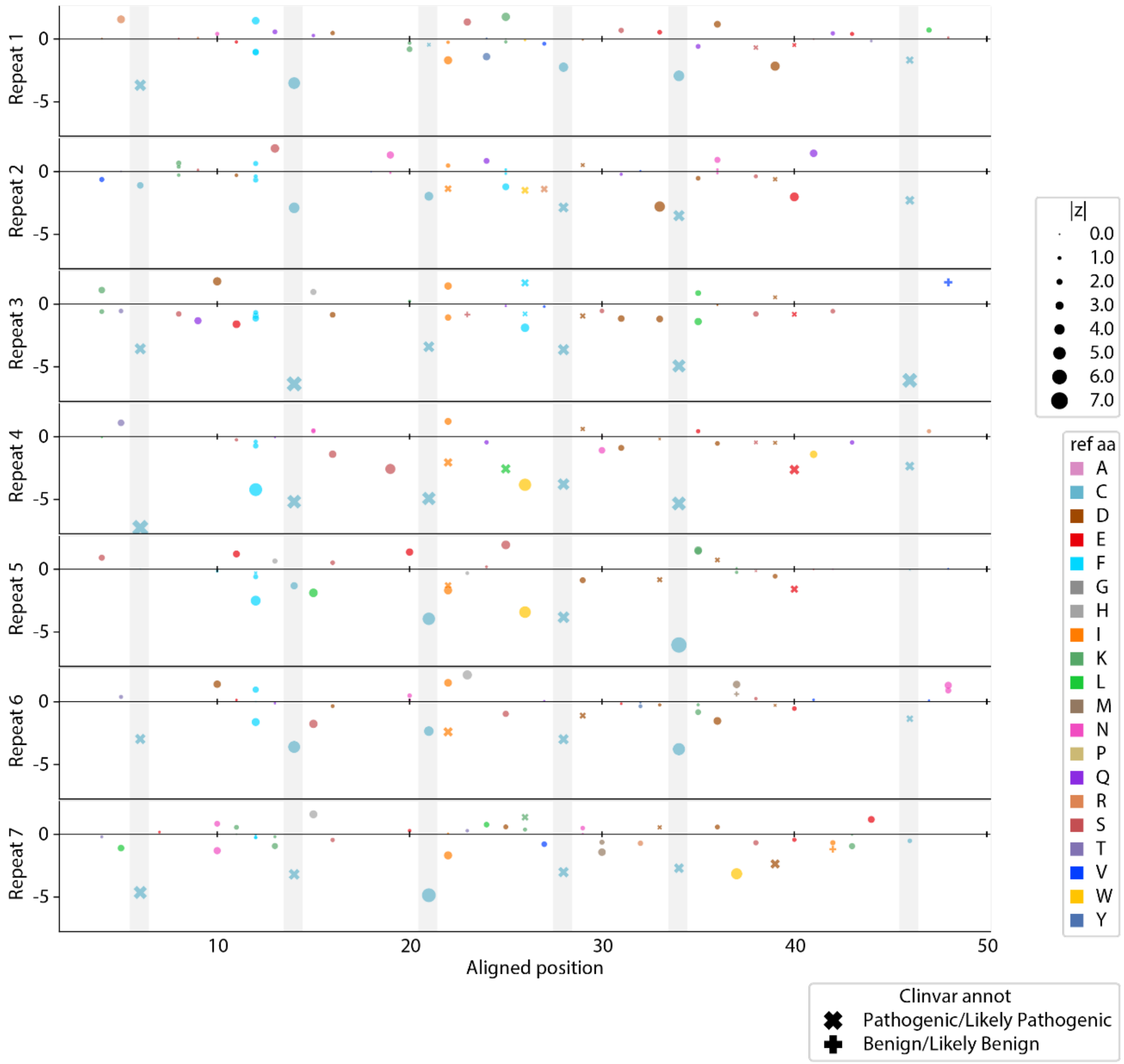


Supplementary Figure 19. BEAN z-scores across LDLR class A repeats. Conserved cysteine residue positions are highlighted in grey.

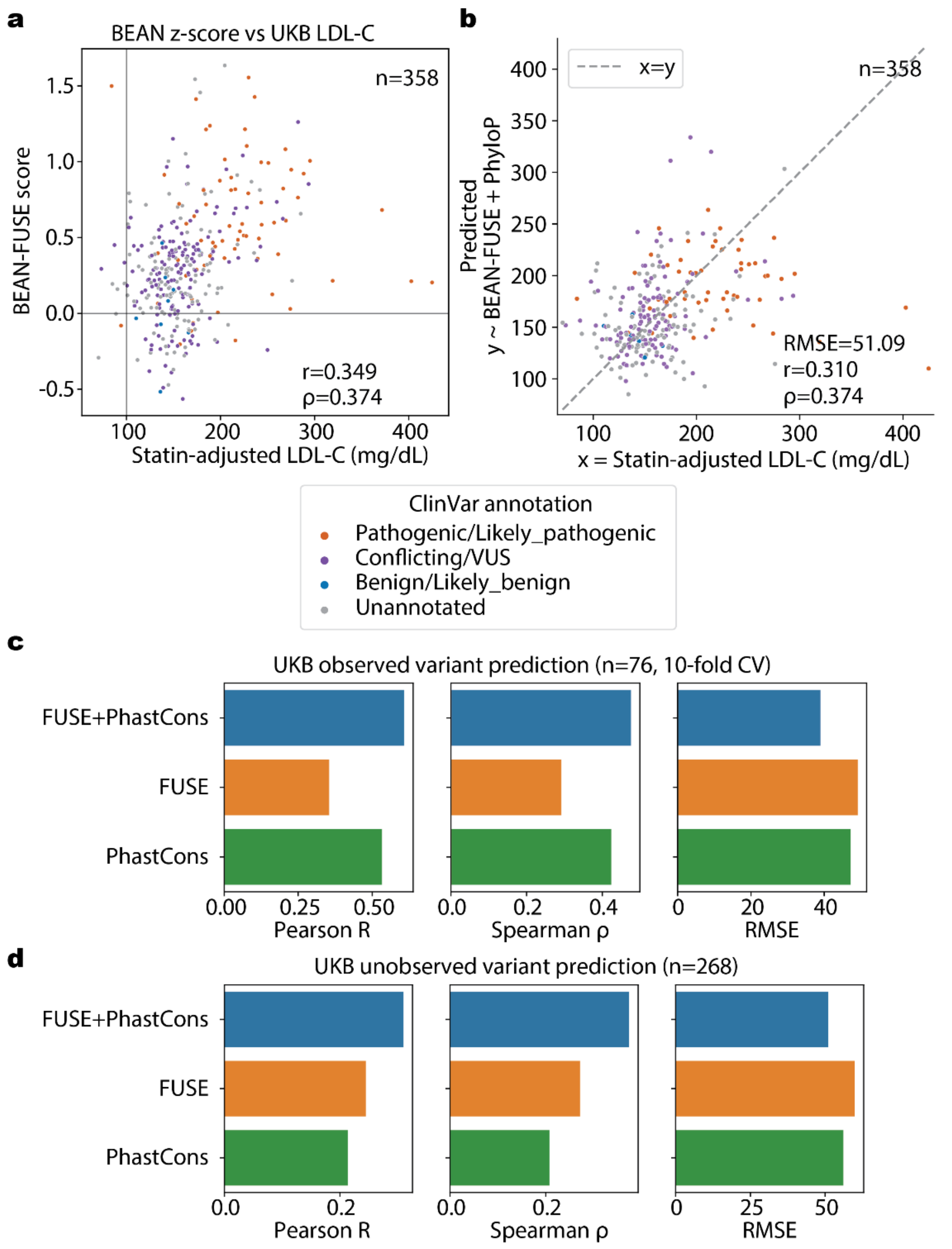


Supplementary Figure 20. Expanded LDLR missense variant pathogenicity estimates with FUSE. a) Scatterplot of all considered UKB variant mean statin-adjusted LDL-C level against imputed BEAN-FUSE score. b) Prediction outcome of unobserved variants with XGBoost model trained on observed UKB variants and mean statin-adjusted LDL levels. c-d) Correlation coefficients and root mean squared error (RMSE) for predicted and true UKB mean statin-adjusted LDL-C level for XGBoost model with FUSE score, PhastCons PhyloP conservation score, and both as the input in predicting LDL-C levels of c) observed variants with 10-fold cross validation and d) unobserved variants with model trained on observed variants.

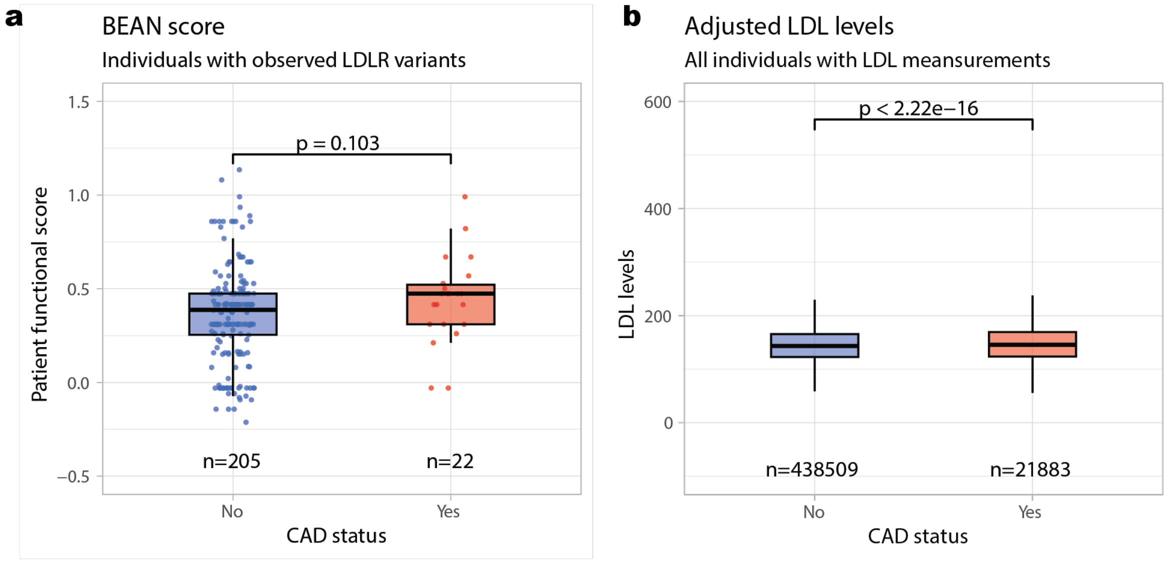


Supplementary Figure 21. Functional scores of individuals with UKB variants stratified by CAD status. a) BEAN z-score of variants observed in UKB stratified by CAD status. b) LDL-C levels of all UKB individuals with LDL-C and CAD status annotation by CAD status. P-values of two-sided Wilcoxon rank-sum test is denoted.

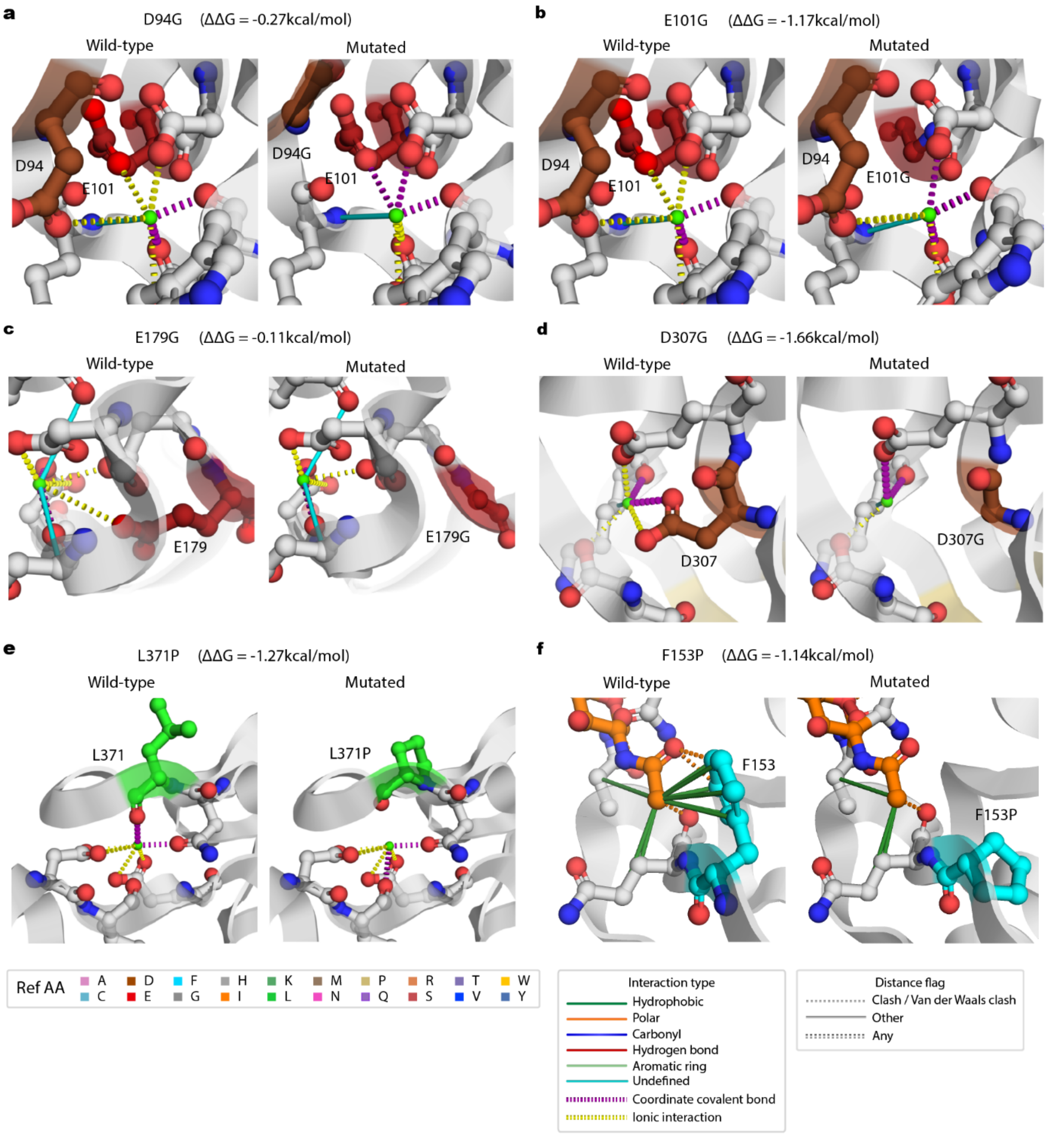


Supplementary Figure 22. Local atomic interaction in wild type and mutated structure for selected variants in LDLR class A repeat and EGF-like domains. Residues with interaction with the variant position are shown. Variant position and interacting residues are colored by the reference amino acid and atomic elements (O: red, N: blue, S: yellow). Ref AA; reference amino acid. The saccharide in panel f is colored as orange; Interaction types and distance flags from Arpeggio output is shown in the legend. Other; distance flag other than clash or Van der Waals clash, Any; coordinate covalent bond and ionic interaction.

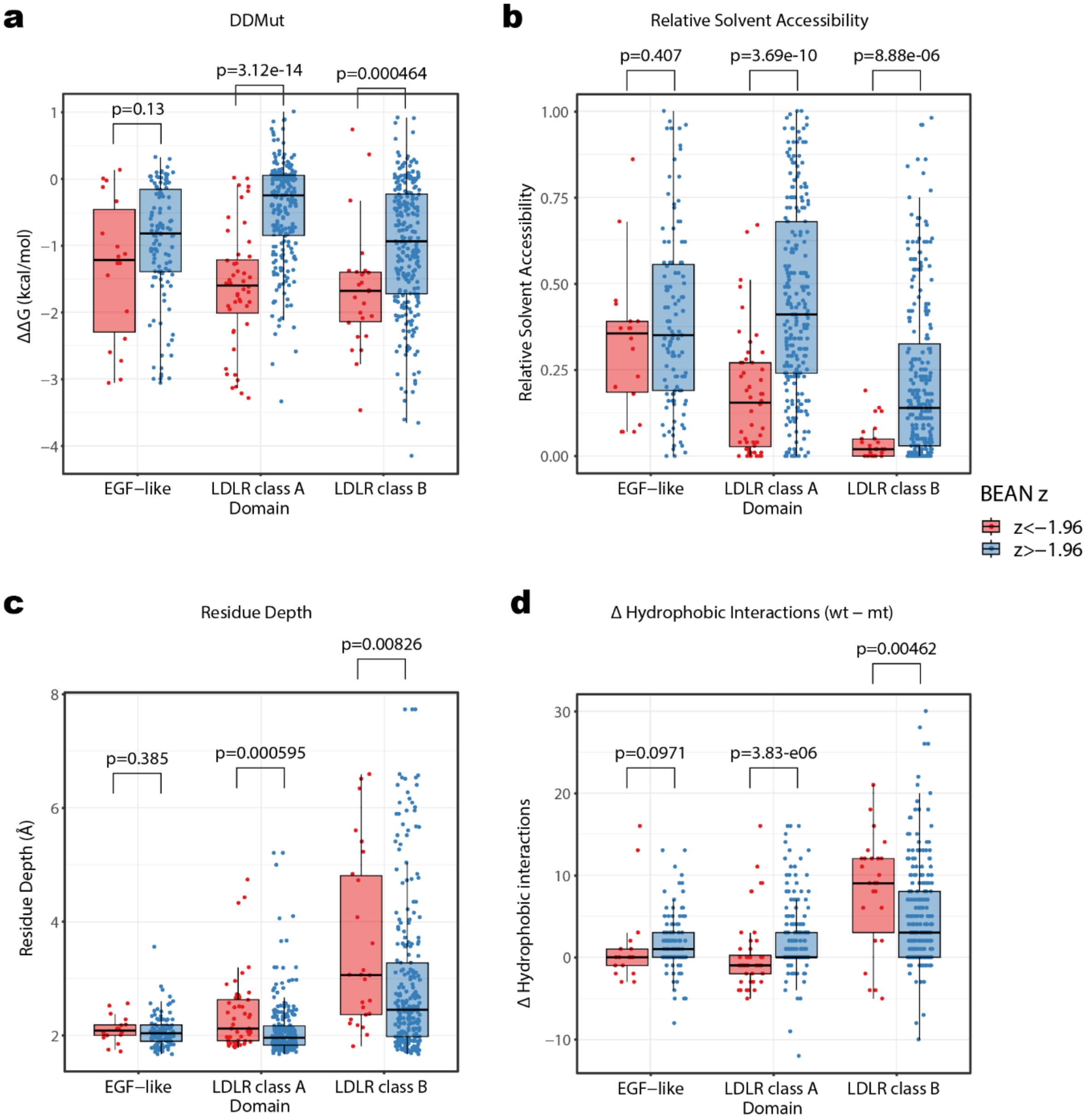


Supplementary Figure 23. Boxplot of structural characteristics of missense variants observed in LDLR tiling screens in LDLR class A repeat domain (48 variants with BEAN z<-1.96, 233 with z>-1.96), LDLR class B repeat domain (26 variants with z<-1.96, 259 with z>-1.96), and EGF-like domain (18 with z<-1.96, 115 with z>-1.96) plotted by BEAN z score threshold. a) Boxplot of DDMut-predicted $\Delta\Delta G$. b) Boxplot of relative solvent accessibility. c) Boxplot of residue depth of reference amino acid. d) Boxplot of lost hydrophobic interactions in mutated amino acid relative to the wild-type amino acid. P-values of two-sided Wilcoxon rank-sum test is denoted.

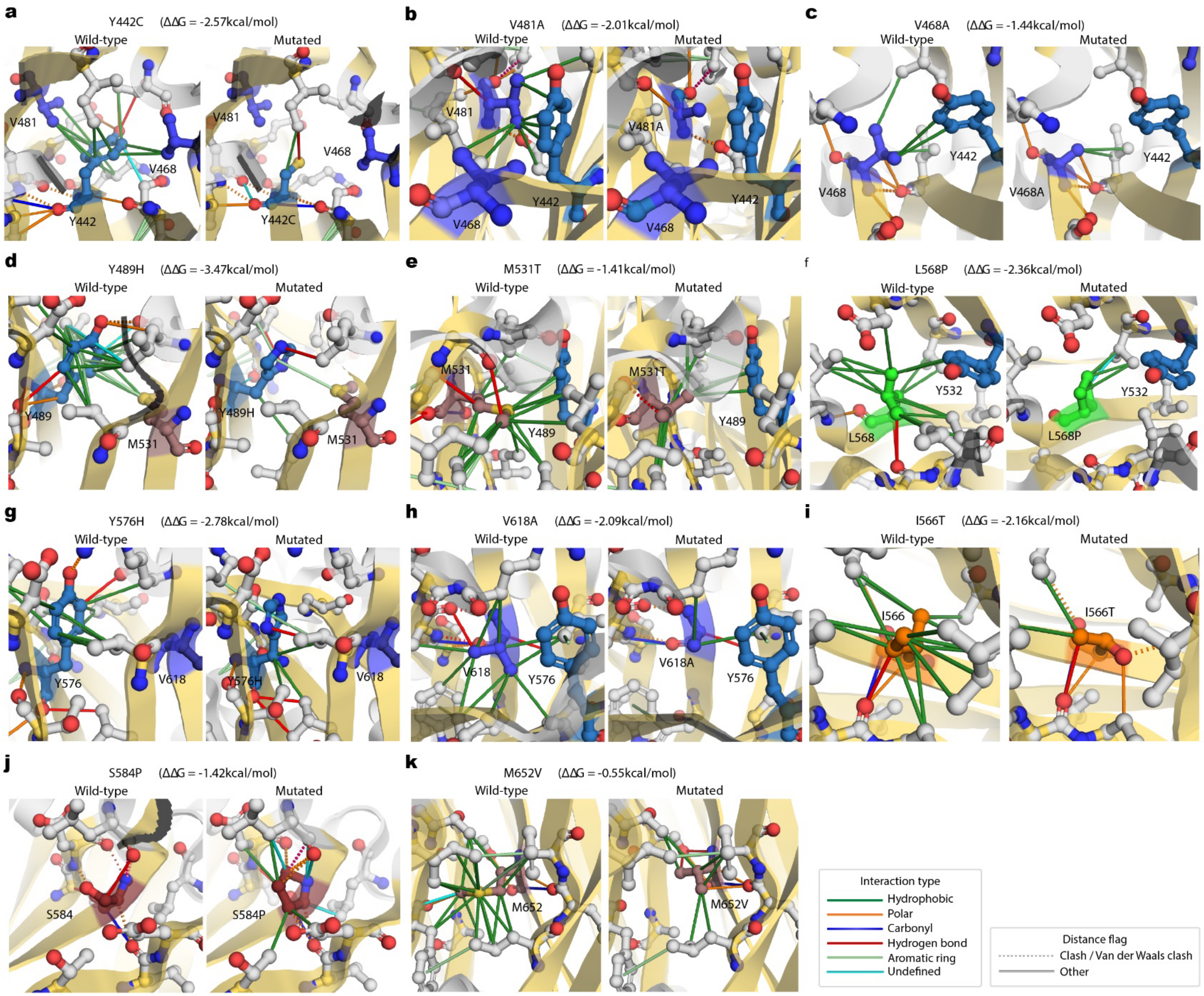
a
Y442C (ΔΔG = -2.57kcal/mol)
Wild-type
Mutated
V481
V468
Y442
Y442C
b
V481A (ΔΔG = -2.01kcal/mol)
Wild-type
Mutated
V481
V481A
V468
Y442
c
V468A (ΔΔG = -1.44kcal/mol)
Wild-type
Mutated
Y442
V468
V468A
d
Y489H (ΔΔG = -3.47kcal/mol)
Wild-type
Mutated
Y489
Y489H
M531
e
M531T (ΔΔG = -1.41kcal/mol)
Wild-type
Mutated
M531
M531T
Y489
f
L568P (ΔΔG = -2.36kcal/mol)
Wild-type
Mutated
Y532
L568
L568P
g
Y576H (ΔΔG = -2.78kcal/mol)
Wild-type
Mutated
Y576
Y576H
V618
h
V618A (ΔΔG = -2.09kcal/mol)
Wild-type
Mutated
V618
V618A
Y576
i
I566T (ΔΔG = -2.16kcal/mol)
Wild-type
Mutated
I566
I566T
j
S584P (ΔΔG = -1.42kcal/mol)
Wild-type
Mutated
S584
S584P
k
M652V (ΔΔG = -0.55kcal/mol)
Wild-type
Mutated
M652
M652V
Interaction type
Hydrophobic
Polar
Carbonyl
Hydrogen bond
Aromatic ring
Undefined
Distance flag
Clash / Van der Waals clash
Other

Supplementary Figure 24. Local atomic interaction in wild type and mutated structure for selected variants in LDLR class B repeat domain. Residues with interaction with the variant position are shown. Variant position and interacting residues are colored by the reference amino acid and atomic elements (O: red, N: blue, S: yellow). Ref AA; reference amino acid.

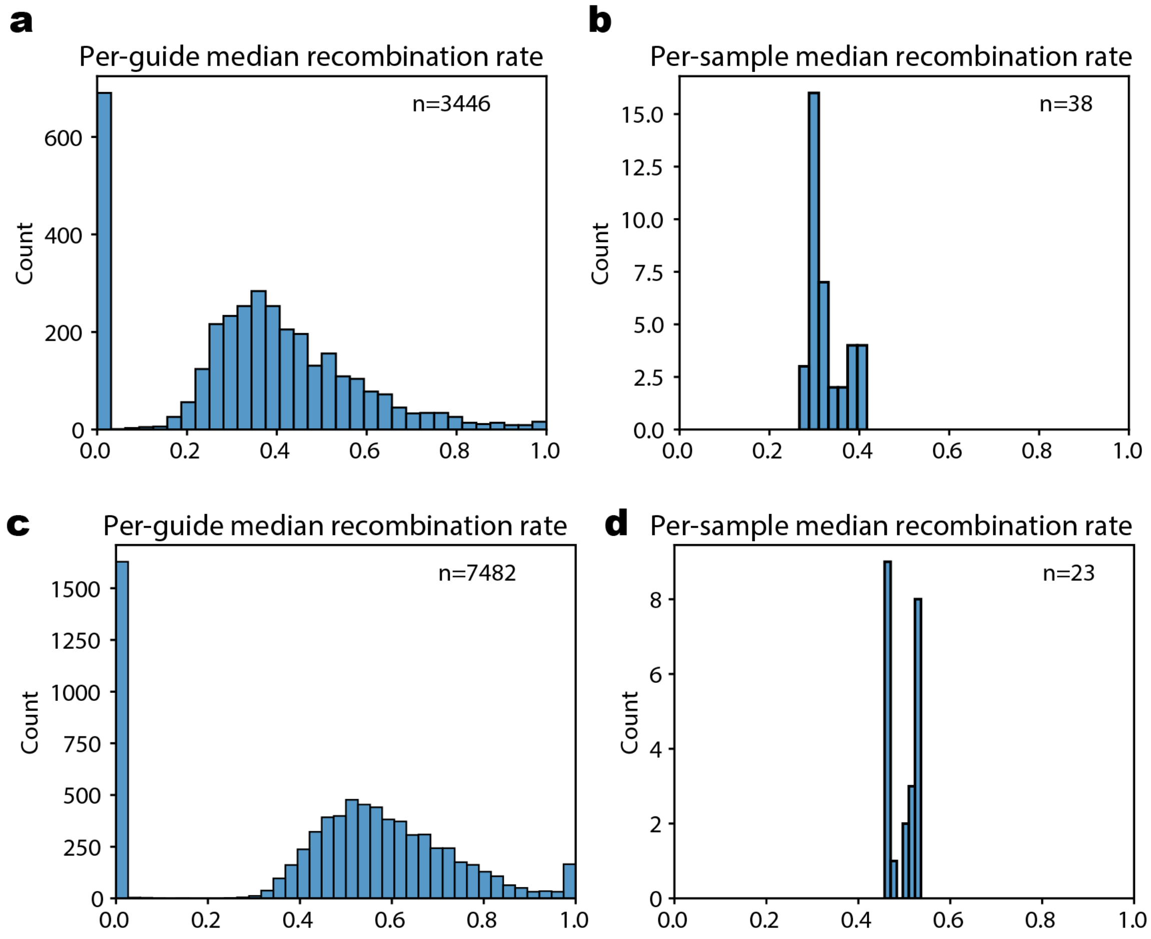


Supplementary Figure 25. GRNA-reporter recombination in lentiviral libraries. Median read 1 and read 2 recombination rates of a-b) LDL-C GWAS library and c-d) LDLR tiling library. Number of gRNAs with more than 10 samples are considered, and samples passed the quality controls are plotted in b) and d). Number of guides or samples plotted for the histogram is denoted as n.

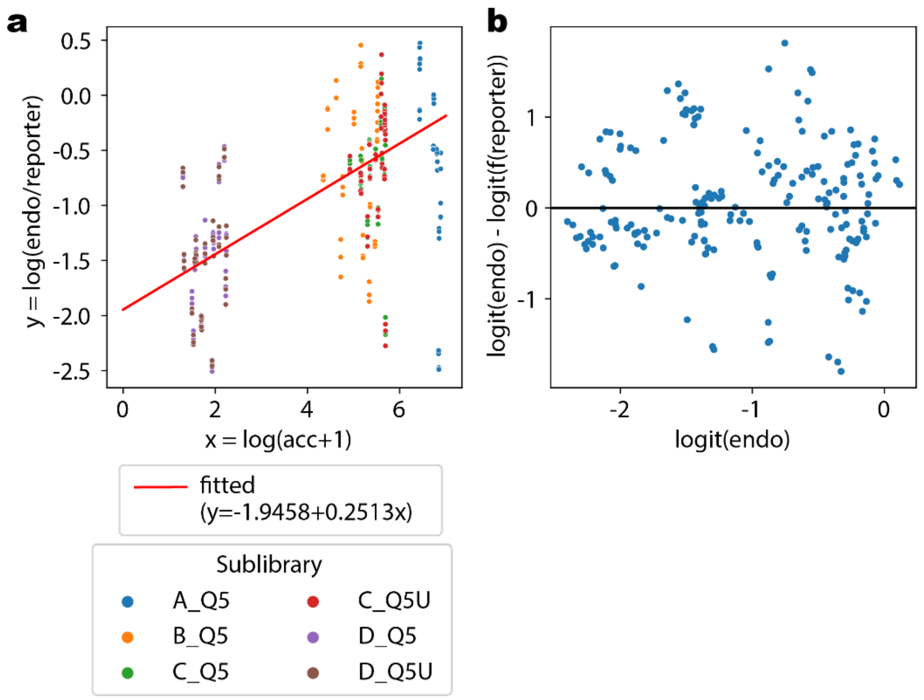


Supplementary Figure 26. Fitting the parameters for accessibility scaling in BEAN with endogenous comparison screen data. a) The relationship between log-transformed ATAC-seq signal (acc) and the log-ratio between endogenous (endo) and reporter editing rate per nucleotide. b) Residual plot of endogenous editing rate and its predicted values by the relationship fitted in a) in logit scale.

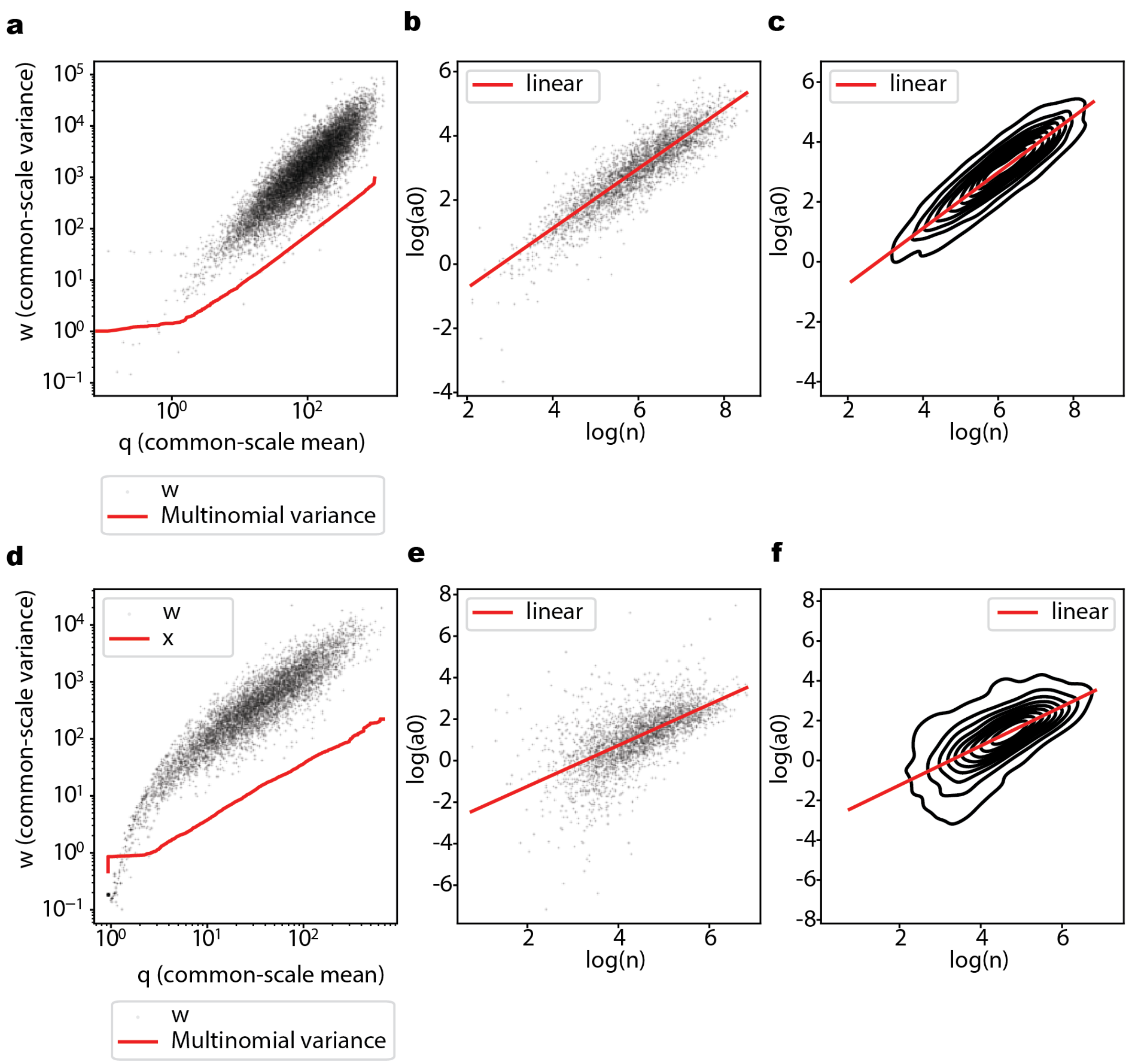


Supplementary Figure 27. Dirichlet-Multinomial parameter fitting from LDL-C GWAS library data. a-c) Estimated parameters for gRNA counts across sorting bins modeled as Dirichlet-Multinomial distribution. a) Mean-variance relationship from the normalized counts. b) Scatterplot of normalized total counts ($n$) plotted against estimated sum of concentration ($\alpha^{\circ}$) and linear fitted relationship in log-transformed scale. c) Data in b) plotted as contour plot. d-f) Estimated parameters for allele counts per gRNA modeled as Dirichlet-Multinomial distribution. d) Mean-variance relationship from the normalized counts. e) Scatterplot of normalized total counts ($n$) plotted against estimated sum of concentration ($\alpha^{\circ}$) and linear fitted relationship in log-transformed scale. f) Data in e) plotted as contour plot.
